# Genetic architecture of brain age and its casual relations with brain and mental disorders

**DOI:** 10.1101/2023.01.09.23284310

**Authors:** Esten H. Leonardsen, Didac Vidal-Piñeiro, James M. Roe, Oleksandr Frei, Alexey A. Shadrin, Olena Iakunchykova, Ann-Marie G. de Lange, Tobias Kaufmann, Bernd Taschler, Stephen M. Smith, Ole A. Andreassen, Thomas Wolfers, Lars T. Westlye, Yunpeng Wang

## Abstract

The difference between chronological age and the apparent age of the brain estimated from brain imaging data — the brain age gap (BAG) — is widely considered a general indicator of brain health. Converging evidence supports that BAG is sensitive to an array of genetic and non-genetic traits and diseases, yet few studies have examined the genetic architecture and its corresponding causal relationships with common brain disorders. Here, we estimate BAG using state-of-the-art neural networks trained on brain scans from 53,542 individuals (age range 3-95 years). A genome-wide association analysis across 28,104 individuals (40-84 years) from the UK Biobank revealed eight independent genomic regions significantly associated with BAG (p<5×10^−8^) implicating neurological, metabolic, and immunological pathways – among which seven are novel. No significant genetic correlations or causal relationships with BAG were found for Parkinson’s disease, major depressive disorder, or schizophrenia, but two-sample Mendelian randomization indicated a causal influence of AD (p=7.9×10^−4^) and bipolar disorder (p=1.35×10^−2^) on BAG. These results emphasize the polygenic architecture of brain age and provide insights into the causal relationship between selected neurological and neuropsychiatric disorders and BAG.

## Introduction

Over the last decade, brain age has emerged as a promising measure of overall brain health^1, 2^. To estimate brain age, machine learning models are applied to brain imaging data to learn visual patterns characteristic of different ages^3, 4^. The difference between predicted brain age and chronological age is termed the brain age gap (BAG) and indicates deviation from a normative aging trajectory, a potential health indicator. Earlier studies have found a large variation in the predicted brain age of individuals with the same chronological age, and that these interindividual variations correlate with neurological and mental disorders^5-7^, such as dementia^6, 8^, schizophrenia (SCZ)^9, 10^, major depressive disorder (MDD)^11^, and also mortality^7,12^. In addition, biological, environmental and lifestyle factors associated with these disorders have been reported to correlate with BAG, for example infections^13, 14^, smoking^5^, physical activity^15^, and education level^16^.

Genetic differences have been shown to explain a sizeable portion of interindividual variation in BAG. Twin-based heritability for BAG has been estimated to be as high as 0.66^17^, and single nucleotide polymorphism (SNP)-based heritability estimates are also relatively high — around 0.2 ^6, 18^. Earlier gene-discovery efforts investigating genetic associations with BAG have found and examined two genomic loci in detail: one on chromosome 1 containing the potassium channel gene, *KCNK2*, and one in the chromosome 17 inversion region (17q21.31)^18, 19^; Genetic variants in these two regions together explain a negligible fraction of estimated SNP-heritability^18^. These results suggest that existing GWAS were potentially underpowered to fully characterize the genetic architecture, supported by studies using a conditional false discovery rate-based models yielding a larger set of associations^6^. Furthermore, Smith *et al*.^20^ found a rich set of associations when investigating different facets of a multimodal brain age, suggesting that the interplay between genetic variants is complex.

Although BAG has been frequently associated with mental disorders, the underlying genetic basis for the observed association has seldom been investigated, possibly due to incomplete knowledge of the genetic architecture of the former. Furthermore, the causal relationships between BAG and brain disorders remain untapped. Mendelian randomization (MR) has become an attractive tool to interrogate cause-effect relationships between risk factors or disorders^21^. Two-sample MR models have been used to infer causal relations between hundreds of traits or diseases^22^. However, MR analyses targeting the causal relations between BAG and brain disorders and associated traits have been lacking^23^.

In the present work, we improve the yield of genetic associations for BAG using three strategies: First, we estimate brain age using a state-of-the-art deep neural network architecture (SFCN-reg) trained on one of the largest samples assembled to date^5^. Then we perform a GWAS for BAG on out-of-sample predictions for a portion of the UK Biobank v3 data containing 28,104 unrelated individuals, about eight thousand more than earlier studies. Finally, we use two-sample MR to assess the genetic and causal relations between BAG and SCZ, bipolar disorder (BIP), Alzheimer’s disease (AD), MDD, and Parkinson’s disease (PD).

## Methods

### Sample

All datasets used in the present study have been obtained from previously published studies which have been approved by their respective institutional review board, research ethic committee or other relevant ethic organizations.

We used UK Biobank imaging data (UKB, accession number 27412) released in 2019 in combination with a pre-compiled dataset from various sources (**Supplementary Table S1**) for brain age model training and estimation. For the downstream genetic analyses, we started with the initial 40,330 UKB participants that had undergone at least one brain scan (using baseline scans where more were available). We excluded those with recorded brain injury or neurological or psychiatric conditions, those failing standard image quality checks^5^, those whose genetic data failed quality check by the UKB team, in addition to participants who withdrew consent. Among related participants (UKB-reported kinship coefficient >0), only one was included in the study. In total, 28,104 unique participants remained.

### Brain age estimation

A minimal preprocessing protocol was applied to all raw T1-weighted brain MRI images before brain age estimation^5^: The auto-recon pipeline from FreeSurfer 5.3^24^ was used to remove non-brain tissue. The resulting volumes were reoriented to the standard FSL^25^ orientation using fslreorient2std, and linearly registered to the 1 mm FSL (version 6.0) MNI152 template using FLIRT^26^, with 6 degrees of freedom. For efficiency, during model fitting, we cropped a central cube spanning the voxels 6:173, 2:214, and 0:160 in the sagittal, coronal, and axial dimensions, respectively. Before modelling, all voxel intensities were normalized by a constant factor to produce values in the range [0, 1].

The data from all sources (**Supplementary Table S1** and UKB) were split into five equally-sized and disjoint folds with comparable age-ranges and sex distributions. Four of these folds were used for fitting the brain age model, and out-of-sample estimates were computed for the remaining fold. This procedure was repeated five times, resulting in out-of-sample brain age estimates for all participants. Next, BAG was calculated by subtracting chronological age from estimated brain age. The subsequent analyses were performed on the out-of-sample estimates of the UKB data (**Supplementary Table S2**).

### Genome-wide association study

Imputed genotypes for the 28,104 participants were obtained from UKB (Category 100314, for further details see^27^). We excluded SNPs based on missing rate (>0.02), the Hardy-Weinberg Equilibrium test (p<10^−6^) and minor allele frequency (MAF; < 0.01). In total, ≈ 8.6 million SNPs were analyzed. Since we have observed apparent differences in predicted brain age across folds (**Supplementary Fig. S1**), a GWAS was performed on each hold-out fold separately using PLINK 1.90 beta^28^. The additive genetic model was assumed, and chronological age, sex and the top ten principal components were included as covariates, accounting for population structure. Association results for each hold-out fold of UKB along with distributions of BAG are shown in **Supplementary Figure S1**. These association results were then meta-analyzed using the inverse variance weighted model implemented in PLINK to identify SNPs that are associated with BAG. **Supplementary Figure S2** shows the association QQ plot which indicated no noticeable genomic inflation.

### Associated regions and genes

Association results were ‘clumped’ by the FUMA^29^ web-service using the linkage disequilibrium (LD) structure from the 1000 Genomes projects phase 3 EUR dataset (1KGp3), with parameters *–clump-p 5e-8 –clump-2 1e-6 –clump-r2 0*.*1*. The standard 250 kilo-bases (kb) were used as the inter-region distance threshold. Genes whose genomic coordinates located within the boundaries of each region were assigned to the corresponding region. SNPs with the smallest association p-values were taken as the lead SNPs for the corresponding regions. In addition, the gene that is closest to each lead SNP by genomic position was annotated using the Ensembl tool VEP^30^ (**Table 1**).

**Table 1.** Genomic loci associated with BAG.

| Locus | Lead SNP | POS | Gene | A1 | A2 | Beta | I<sup>2</sup> | P |
| --- | --- | --- | --- | --- | --- | --- | --- | --- |
| **Chr3:183892867-183975709** | rs73185796 | 183975709 | CAMK2N2/ECE2 | T | G | -0.29 | 0.0 | 2.53x10<sup>-8</sup> |
| **Chr4:38591172-38779512** | rs13132853 | 38680015 | KLF3 | G | A | 0.23 | 0.0 | 2.34x10<sup>-18</sup> |
| **Chr5:78388694-78451813** | rs79107704 | 78388694 | BHMT2 | A | G | 0.63 | 0.0 | 1.65x10<sup>-8</sup> |
| **Chr6:45407654-45511945** | rs2790102 | 45432214 | RUNX2 | A | G | -0.15 | 0.0 | 8.92x10<sup>-9</sup> |
| **Chr8:124661974-124682971** | rs7461069 | 124669029 | KLHL38 | A | G | -0.17 | 0.0 | 1.57x10<sup>-8</sup> |
| **Chr10:134544247-134597265** | rs4880424 | 134584577 | INPP5A | T | C | 0.16 | 64.95 | 3.69x10<sup>-8</sup> |
| **Chr14: 88391116-88556525** | rs17203398 | 88449847 | GALC | C | G | -0.16 | 40.97 | 1.42x10<sup>-10</sup> |
| **Chr17: 43101281-44863413** | rs2106786 | 43919096 | MAPT | G | A | 0.29 | 0.0 | 1.87x10<sup>-23</sup> |

Eight independent genomic loci significantly associated with brain age gap (BAG). Lead SNP rs-number, genomic position (in hg19 coordinates), effective allele (A1), the other allele (A2), effect size (Beta), meta-analysis heterogeneity (I<sup>2</sup>), association strength (p value, P).

Associated regions were fine-mapped using the FINEMAP^31^ program. The LD structure from 1KGp3 was also used in this analysis. The default settings of FINEMAP were used, which compares causal models assuming one causal variant in each region to that assuming two, based on the estimated posterior probabilities (PP_1 versus PP_2). FINEMAP ranks all possible configurations in each model presented as 95% credible sets. The confidence of a variant belonging to a set was evaluated by posterior probabilities of inclusion (PPI). In the case of assuming one causal variant, each single variant was assigned a PPI. In the two causal variants cases, each pair of variants was assigned a PPI.

### Post-GWAS annotations

Both FUMA and Garfield^32^ were used for annotating associated SNPs. First, SNPs were assigned to genic elements (*e*.*g*., exon, intron, 3’ and 5’ untranslated regions, intergenic regions, etc.), and the enrichment of this assignment was tested by hypergeometric test (FUMA) or logistic regression models (Garfield). Expression levels of annotated genes to the associated SNPs were inspected in each of the 54 tissue types from the GTEx v8 dataset^33^. Detailed biological functions for proteins coded by these genes were manually searched in the NCBI Entrez Gene database^34^ and the UniProtKB database^35^.

### Genetic correlations between BAG and disorders

GWAS summary data for four disorders (SCZ^36^, BIP^37^, MDD^38^, and AD^39^) were obtained from the Psychiatric Genomics Consortium (PGC, https://med.unc.edu/pgc/download-results). For each GWAS, the association results for European ancestral samples excluding samples from 23andMe were used (SCZ: n case=67,390, n control=94,015; BIP: n case=41,917, n control=371,549; MDD: n case=59,851, n control=113,154; AD: n case=71,880, n control=383,378). The PD GWAS results were obtained from the fixed-effect meta-analysis performed by the International Parkinson Disease Genomics Consortium (IPDGC, n case=33674, n control=449056)^40^.

Before post-GWAS analysis we processed the results from all GWAS using a standard protocol. Specifically, SNPs having a MAF <0.05, or imputation INFO <0.5, or ambiguous allelic coding (A/T, or C/G) were removed from subsequent analyses. The LD score model (ldsc)^41^ was applied to estimate SNP-heritability and genetic correlations between BAG and disorders. Only high-quality SNPs published in the HapMap3 dataset were used for estimation. The LD score derived from the 1KGp3 was used as input to ldsc. The Benjamini-Hochberg False Discovery Rate (FDR) procedure was used to correct for multiple testing across disorders (FDR-corrected p<0.05 was considered statistically significant).

To visualize polygenic enrichment, conditional QQ plots^42, 43^ were made for BAG versus each disorder. In these plots, the QQ curves for the association statistics (-log10 p-values) for BAG were stratified by the corresponding association strength for the conditioned disorder. As the association strength to the conditioned disorder increases, a successive leftward deflation in these curves indicates polygenic enrichment. Similarly, conditional QQ for each disorder versus BAG shows polygenic enrichment in the reverse direction.

### Two-sample Mendelian randomization

To study the cause-effect relations between BAG and the five disorders, two sets of MR analyses were performed. The first set, using standard models, included the inverse-variance weighted model (IVW)^44^, weighted median (wMed)^45^, Egger regression (Egger)^46^ and MR-PRESSO (PRESSO)^47^. For these analyses, only genome wide significant SNPs (p<5×10^−8^) to the exposure traits or disorders were used as potential instruments. The PLINK program and the LD structure of 1KGp3 dataset were used to select instruments with the following parameters, --*clump-kb 500 kb, --clump-p1 5×10*^*-8*^, and *--clump-r2 0*.*01*. The TwoSampleMR package^19^ was used for data harmonization and causal inference for the IVW, wMed and Egger models. The same harmonized datasets were used as input to the MR-PRESSO software to assess outliers that may artificially affect MR estimates, *i*.*e*., SNPs that show horizontal pleiotropy to both BAG and disorders. Harmonized instrumental SNPs are shown in **Supplementary Tables S6**-**S15**.

The second set of models included the robust adjusted profile score (RAPS)^48^ and the CAUSE models^49^. These models can make use of SNPs that show a suggestive level of association (p<10^−3^) with exposure to increase statistical power without incurring weak instrument bias in estimation. While both models control for horizontal pleiotropy, CAUSE directly tests for a shared (correlated horizontal pleiotropy) versus a causal model for each relation^49^. Instead of estimating causal effects, CAUSE provides z scores for such tests; negative z scores suggest the existence of a causal relation and positive suggest otherwise. The same instrument selection procedures used in the first set of models were used here, except that 10^−3^ was taken as the cut-off for selecting instruments, *i*.*e*. –*clump-p1 10*^*-3*^.

As each of the six MR models has different assumptions that are difficult to verify in real data, a majority vote ensemble scheme was used to make conclusions for the existence of cause-effect relations: specifically, only when four or more models indicated a cause-effect relation (FDR adjusted p<0.05) was such a relation considered causal.

In addition to applying multiple MR models, GWAS results for height measured for European samples^50^ (n=253,288; https://portals.broadinstitute.org/collaboration/giant/), for AD diagnosed in a Japanese sample^51^ (n case=3,962 and n control=4,047) and an African sample^52^ (n case=2,784 and n control=5,222) and for BIP diagnosed in a Japanese sample^53^(n case=2,964 and n control=61,887) were used to corroborate MR findings. As commonly done in genetic studies, height was used as a negative exposure control to test if population stratification could generate spurious causal effects ^54^. The non-European GWAS data were used to test if any observed causal effects generalize across ancestry groups, although with significantly smaller sample sizes.

## Results

We obtained accurate brain age estimates; mean absolute errors (MAEs) in each of the five disjoint folds were consistently below 2.5 years (**Supplementary Table S2**). This was consistent when we split the dataset into different subsets based on covariates (MAE = 2.40 in females compared to 2.53 in males; 2.40 in the youngest half compared to 2.52 in the oldest), although we observed a slight age bias (**Supplementary Figure S9**). Based on the meta-analyzed GWAS results, we estimated a SNP heritability of 0.27 (standard error (SE)=0.036) for BAG (**Methods**). Our estimate is comparable to or higher than the two previously reported estimates (0.26, SE=0.044^6^; 0.19, SE=0.02^18^).

We identified eight independent genomic loci significantly associated with BAG (**Fig. 1 a**,**b** and **Table 1**). Associations of lead SNPs in these regions to BAG are highly consistent in directions and effect sizes across the five folds (**Supplementary Table S3**). Among these loci, the one in the inversion region on chromosome 17 (lead SNP rs2106786), including the *MAPT* gene, has been previously reported^18, 19^, although indexed by a different SNP (rs2435204). This SNP was also highly significant in our analysis (p=5.4×10^−21^, beta=0.27 years, effective-allele=G). The locus containing the *RUNX2* gene (lead by rs2790102: p=8.92×10^−9^, beta=-0.15 years, effective-allele=A), which showed suggestive significance in Jonsson *et al*.^18^, was genome-wide significant in the present study. The *RUNX2* gene codes for a master transcription factor which plays a critical role in skeletal development^55^. Among the remaining six novel loci, the rs79107704-A allele showed the largest association with BAG; one copy of this allele was associated with an average increase in brain age of 0.63 years (**Table 1**). This SNP is located 3,405bp downstream of the Betaine-homocystein S-methyltransferase 2 gene (*BHMT2*, **Fig. 1b**), a gene whose product is involved in choline metabolism during development^56^. Other protein-coding genes that are closest to the lead SNPs include those involved in calcium signaling (*CAMK2N2* and *INPP5A*) and metabolism and transcription regulation (*GALC, KLF3* and *KLHL38*), both processes are implicated in biological ageing^57^. In **Supplementary Table S5** we present detailed annotations of biological functions to each gene.

**Figure 1.**
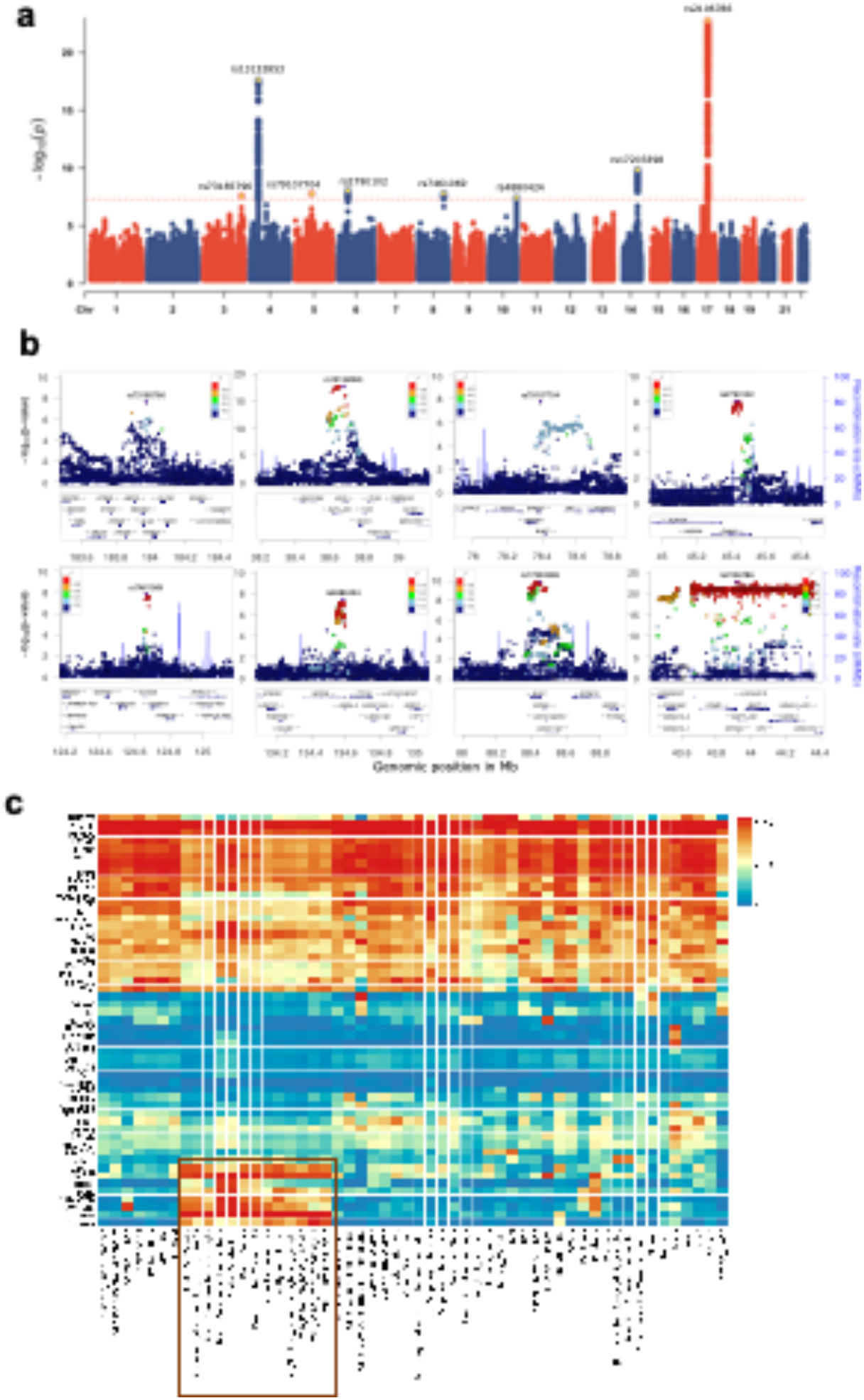
Genetic associations for brain age gap. **a**. The Manhattan plot of meta-analyzed association results for brain age gap (BAG). Chromosome numbers are shown on x axis, -log10 association p values on y axis and lead SNP rs-numbers in the plot. **b**. Region plots for each of the eight associated regions. Genes located in each region are shown below each figure. Linkage disequilibrium r-squared values are indicated by colors; and recombination frequences by curves. **c**. Expression levels of the annotated genes across tissues analyzed by the GTEx v8 study. Colors indicate average log2 transformed expression level in each tissue.

We further annotated these identified SNPs to nearby genes and regulatory elements (**Methods**). Most of the associated SNPs are in non-coding regions such as intergenic, intron or untranslated regions (**Supplementary Figs. S3** and **S4**). Using the default parameters in FUMA^29^, 54 unique genes were found to be implied by these significant associations by genomic position. The expression levels of these genes in the 54 tissue types from the GTEx v8 project^33^ showed three remarkable patterns (**Fig. 1c**). The first set of genes expressed highly across almost all 54 tissues; the second set of genes showed low expression levels in most tissue types; and the last set, including eight genes, was highly expressed only in brain tissue types (**Fig. 1c**), for example *MAPT, GFAP*, and the Homeobox protein gene *NKX6-2*. These results suggest that BAG encodes coordinated physiological processes implicating both the brain and the peripheral systems.

To nominate causal variants in each locus we performed statistical fine-mapping^31^ for regions around each lead SNP in **Table 1** (**Methods**). Except for the locus on chromosome 14 which was not resolvable, all loci clearly indicated that the 95% credible sets suggest a causal model with one causal SNP, instead of two, *i*.*e*., the posterior probability for the 1-SNP set were larger than those of the 2-SNP sets (**Supplementary Table S16**). Furthermore, four credible sets indicated that the lead SNPs were also the causal ones (posterior inclusion probability (PPI_1) > 0.05 and >PPI of the second most probable SNP(PPI_2)) (**Methods**; **Supplementary Table 16**) but identifying the causal SNP for the remaining were difficult. For example, the *MAPT* locus on chromosome 17 and the *RUNX2* locus on chromosome 6 showed two SNPs having almost equal and small PPIs (*i*.*e*., <=0.05), indicating that the true causal variants may be some untyped rare ones not investigated in this study. The clearest signal comes from the regions on chromosome 3 and 5, where the PPIs for the lead SNPs were much larger than that for the second most probable causal SNPs.

We observed nominally significant genetic correlation between BAG and AD that did not survive FDR-correction (r=0.23, SE=0.1, p=0.02, FDR adjusted-p=0.13) and no significant associations with any other of the four disorders (**Fig. 2a**). SNP heritability estimates for the five disorders were all significant but varied greatly; SCZ showed the largest (0.34, SE=0.01) and AD showed the lowest (0.01, SE=0.005) estimates. Bidirectional conditional QQ plots (**Fig. 2b-d**; **Supplementary Figs. S6** and **S7**) showed that there was noticeable genetic enrichment for BIP conditional on BAG but not in the reverse direction. For AD and PD, both directions showed clear enrichment, surprisingly for PD which did not show significant genetic correlation with BAG (r=-0.07, p=0.42).

**Figure 2.**
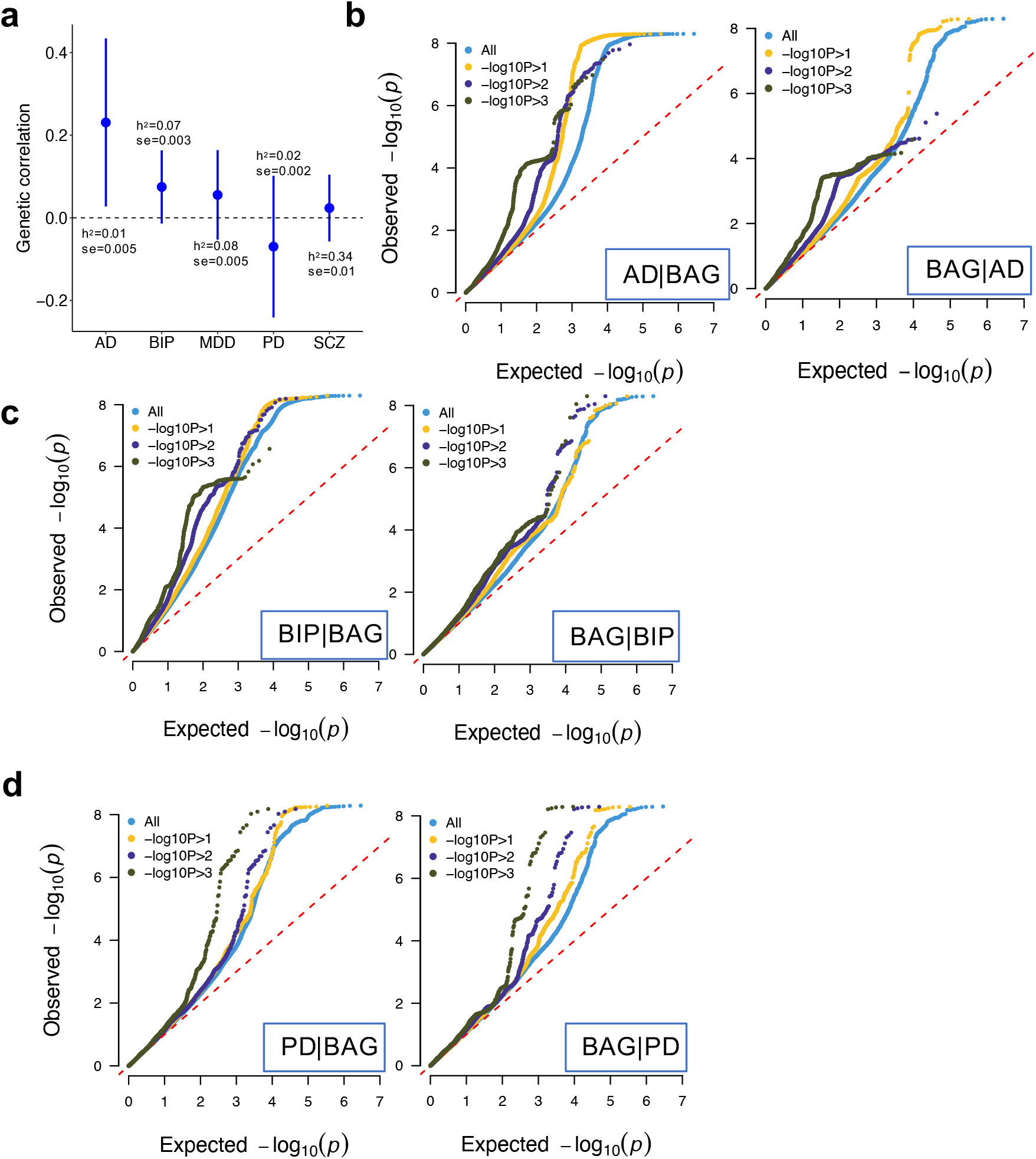
Polygenic genetic overlap between brain age gap and disorders. **a**. Genetic correlation between brain age gap and disorders computed by ldsc. SNP heritability and its standard error are indicated. **b-d**. Conditional QQ plot between brain age gap and disorders in both directions. Colors are used to indicate different association strength to the conditioned traits, i.e., the ones indicated after the vertical bar in each figure. Dashed diagonal lines indicate expected null distributions. Abbreviations used: AD, Alzheimer’s disease; BIP, bipolar disorder; MDD, major depression disorder; PD, Parkinson’s disease; SCZ, schizophrenia.

We then performed extensive MR analyses using six different models to examine the existence of cause-effect relations between BAG and the five disorders (**Methods**). **Figure 3a** shows that BAG was only causally associated with PD, *i*.*e*., four out of the six MR models showed a negative relation with varying effect sizes (all with adjusted p <0.05). One year increase in genetically predicted BAG was estimated to reduce the risk of PD by a log odds ratio from 1.4 (by Egger regression) to 0.02 (by MR-RAPS) (**Supplementary Table S15**). In the reverse direction (*i*.*e*., disorders as exposure), increased genetic risk for AD and BIP were causally associated with increased BAG (30 and 55 SNPs used as instruments, respectively); these estimated causal effects on BAG were relatively larger for AD than BIP (**Fig. 3b; Supplementary Table S16**).

**Figure 3.**
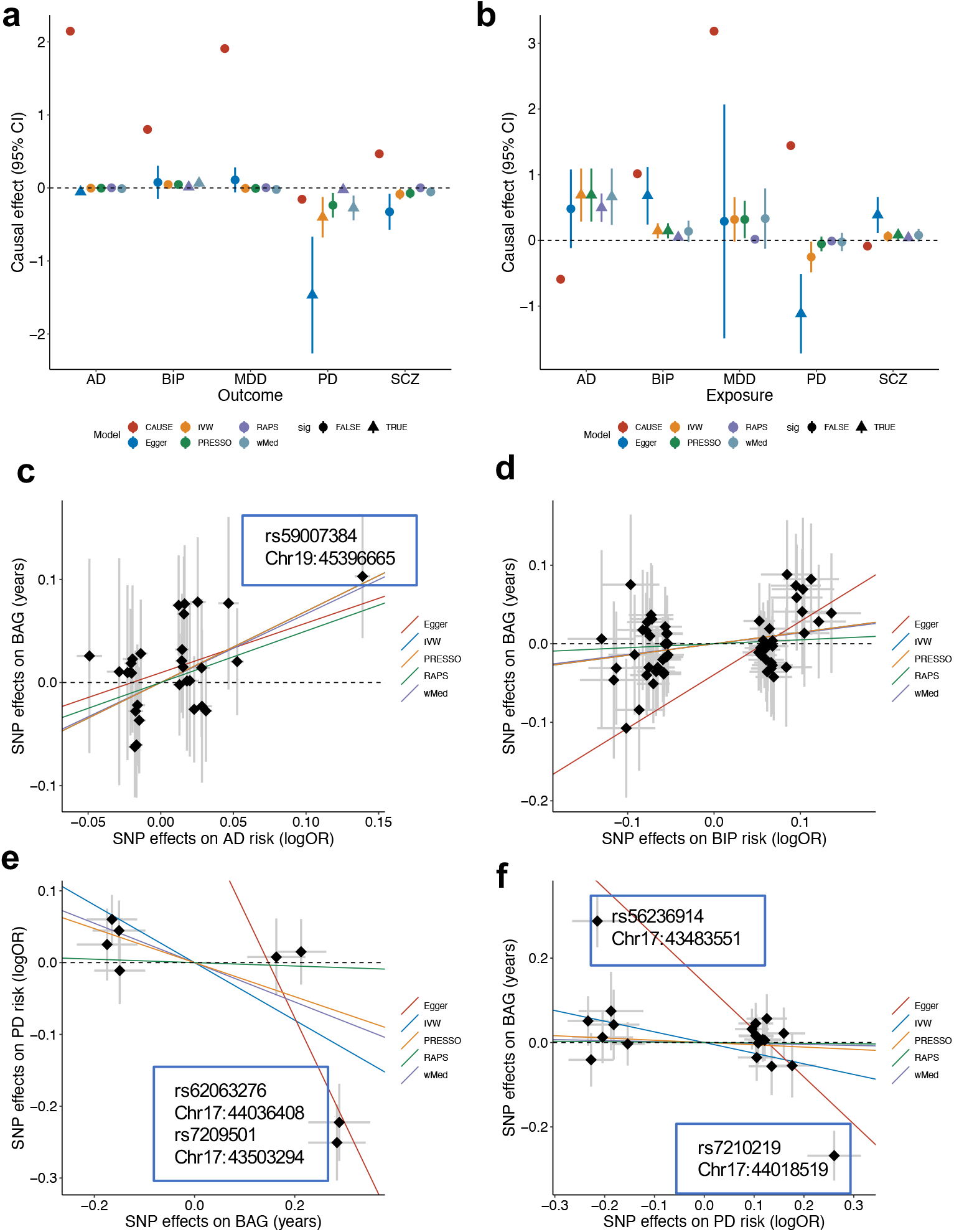
Causal inference between brain age gap and disorders. **a**. Causal effect of brain age gap (BAG) on risk of disorders; **b**. Causal effect of genetic risk of disorders on BAG. Colors indicate different models; triangle indicates significant effect after false discovery correction. Estimated standard errors for each effect are aslo shown. **c**. Scatter plots of SNP effects on AD (x axis) and BAG (y axis). **d**. Scatter plots of SNP effects on BIP (x axis) and BAG (y axis). **e**. Scatter plots of SNP effects BAG (x axis) and PD (y axis). **f**. Scatter plots of SNP effects on PD (x axis) and BAG (y axis). Causal effects estimated by the five models (except CAUSE) are shown by fitted lines; slopes of these lines indicate causal effect sizes. Exceptional SNPs are marked by boxes which include SNP rs-numbers and genome location in th hg19 coordinates. Abbreviation used: AD, Alzheimer’s disease; BIP, bipolar disorder; MDD, major depression disorder; PD, Parkinson’s disease; SCZ, schizophrenia.

A close investigation into the scatter plots of instrumental SNPs showed that the causal effect of AD on BAG was primarily driven by a SNP (rs59007384) in the APOE region, which was not identified as a horizontal pleiotropic instrument by MR-PRESSO (outlier test p>0.05) (**Fig. 3c**); there were no extreme instruments identified for the BIP to BAG relation by MR-PRESSO (**Fig. 3d**) but Egger-regression indicated existence of horizontal pleiotropy (Egger intercept test: p=0.017). The causal effects of BAG on PD were primarily driven by two SNPs in the inversion region on chromosome 17, effective alleles of these SNPs were associated with higher BAG and lower risk of PD (**Fig. 3e**). SNPs in the same region also drove the negative causal relation (not significant) from PD to BAG (**Fig. 3f**). However, both SNPs were flagged as horizontal pleiotropic instruments by MR-PRESSO (p<0.05) and Egger-regression (Egger intercept test: p=0.03 and 0.008, respectively). Therefore, the observed negative relations between BAG and PD are less likely to be causal.

We used the GWAS results for height of European samples and cross-ancestry MR analysis to corroborate the identified causal relations (**Methods**). We found no causal effect between BAG and height with any of the MR methods employed (all p > 0.05). Therefore, our observed AD and BIP to BAG relations are less likely to be driven by population stratification, *i*.*e*., both the exposure and outcome data originating from the same ancestry group. There was also no significant cross-ancestral causal effect detected using AD data from Japanese or African samples (IVW p=0.74, 0,85, respectively), and BIP data from the Japanese sample to BAG (beta=0.10, p=0.13).

## Discussion

Combining the advantages of large samples and advanced models for brain age prediction, we confirmed that BAG is a heritable and polygenic trait, and estimated the genetic pleiotropy and causal genetic relations with major brain and mental disorders. We identified seven novel loci associated with BAG, in addition to confirming the previously reported *MAPT* loci^18, 19^. While MR indicated that increased genetic risk for AD or BIP may be causally associated with higher BAG, our results demonstrate that individual variability and previously reported case-control differences in BAG only to a marginal degree should be attributed to the genetic risk of the respective diseases.

Functional annotation of the genes linked to the identified loci confirms that deviations in BAG are linked to complex processes encompassing multiple biological systems^20^. While earlier work observed this variety when investigating different multimodal aspects of imaging data linked to brain aging, our findings suggest it also exists when looking at a singular BAG computed from only T1-weighted structural imaging. Our coarse division of the implied 54 genes into three groups indicates that only eight genes are specifically expressed in brain tissues. The remaining genes were either expressed in abundance across all tissue types tested, including the brain, or expressed at very low levels across all tissues. Nonetheless, the proteins coded by these non-brain specific genes have been implicated in brain-related disorders or traits (**Supplementary Table S5**). For example, among the genes we found to be expressed across all tissue types (group 1), mutations in *AP2M1* have been linked to epilepsy, intellectual developmental disorder, and seizures^58^; among the genes expressed in low levels across tissues (group 2), *STH* has been associated with frontotemporal dementia and 17q21.31 duplication syndrome^59, 60^. In addition, although our analysis revealed no significant pathway enrichment, these 54 genes contribute to biological functions that include calcium signaling, protein metabolism, DNA damage repair and general innate immune defense. Thus, our analyses highlight the role of these diverse sets of processes affecting the brain throughout life.

Prior work has shown higher BAG in patients with a multitude of disorders compared to healthy controls^5, 6, 8, 9, 11^, and has documented partly overlapping genetic associations between BAG and clinical conditions^6^. However, the causal effects have remained unclear. Our MR approach revealed that genetically predicted risks for AD or BIP were causally associated with increased BAG. However, these relations were only weakly supported by genetic correlation analysis. One possible explanation for this weaker support from genome wide signals (genetic correlation) in contrast to MR (significant associations only) might be due to heterogeneous genetic correlations across the genome, *i*.*e*. some genomic regions show positive correlations while others show negative correlations^61, 62^. In such a scenario, the net genetic correlations between the two traits are expected to be lower than regional correlations.

The causal effect of genetically predicted risk to AD on BAG was small but consistent in directions across the six MR models, four of which were significant after multiple-testing correction. For BIP, although four models showed significant effects, the CAUSE model suggested an opposite direction of effect to the other five models. Thus, we advise careful interpretation of this result. Our attempts of testing across ancestral group causal relations led to largely null fundings for the AD to BAG relations. We believe these non-significant fundings are largely due to the lack of statistical power in the non-European GWAS^51-53^.

The observed causal relations between genetically predicted risk of brain disorders and BAG are intriguing. One possible interpretation is that overt changes in the brain incurred by the disorders contribute to accelerated aging. Another possibility may be that lifestyle and health-related behaviors of patients with clinical conditions such as AD and BIP, *e*.*g*., medication^63^, may increase brain age. Yet another is that genetic variation drives early life changes in brain structure which contribute to both phenotypes later in life. Given the comparable sample sizes to the GWAS of AD and BIP and the widely observed clinical correlations, surprisingly, no genetic nor causal relations of SCZ and MDD with BAG were found. On the one hand this may suggest that previously reported case-control differences do not reflect causal relations, but rather a combination of indirect and confounding factors. For example, smoking and physical exercise have been associated both with MDD and SCZ^64-67^ and brain age^5, 66^. Alternatively, it has been shown that both BAG^20^ and psychiatric disorders are highly heterogeneous phenotypes^68, 69^, and thus further identification and characterization of the causal relations may require even larger, and carefully screened, samples. It is also worth noting that while the sample sizes for the disorders are large, the one underlying our BAG GWAS is relatively small. Thus, our null findings in the direction from BAG to disorders may be due to too weak instruments^70^.

Our initial results showed weak evidence of a causal relation between BAG and PD, corroborating two recent studies which reported a weaker correlation between BAG and PD ^71, 72^. Striking patterns of enrichment between the two were shown in the conditional QQ plots and four out of six MR models indicated that genetically predicted BAG may have protective effect from PD. However, we found that these relations were completely caused by the *MAPT* gene region on chromosome 17: after removing chromosome 17 from our analyses, no enrichment was observed in either direction (**Supplementary Fig. S8**). In addition, instrumental SNPs in this region were detected by MR-PRESSO as horizontal pleiotropic SNPs, *i*.*e*., affecting BAG and PD through independent biological pathways. Reperforming MR analysis excluding these outlier SNPs confirmed null causal relations. Thus, we conclude that we found no evidence for causal relations between genetically predicted risk for PD and BAG. Our analytic procedures also highlight the importance of triangulation and converging evidence in causal inference analysis^73^.

While the present study advances current knowledge regarding the genetic architecture of and causal contributions to BAG, the results should be interpreted with caution. Although we confirm previously reported genetic associations with BAG, e.g., the *MAPT* gene locus^18, 19^, our sample overlaps with previous ones—which were also based on UK Biobank data. We attempted replicating our findings in three independent but small samples (n ranges from 321 to 702; **Supplementary Analysis** and **Table S4**) but no clear replications were achieved. Therefore, independent large-scale samples are needed for replication. We used a simple voting schema across six different MR models to infer causal relations between genetically predicted BAG and brain disorders. Furthermore, as we only identified eight independent loci showing significant associations to BAG, other models^74^ that require large number of genome-wide significant instruments were considered not applicable. However, it should be noted our simple voting approach may not be the most efficient strategy for identifying causal effects. Formal development of ensemble methods, such as bagging^75^, may provide better grounds for precise interpretation. Furthermore, our BAG GWAS is still smaller than GWAS performed for the disorders, which may partly explain the lack of causal effects of BAG on brain disorders. Therefore, to increase our confidence in the identified relations, large-scale data for BAG, and replications in independent datasets are needed. Relatedly, our estimation of brain age was based on cross-sectional samples, which makes its interpretation non-trivial^76^, and studies built on longitudinal data could help disentangle its complexities. Finally, although we refer to our brain age estimation in general terms, it is important to reaffirm that it is based on T1-weighted imaging data. The brain is a complex and heterogeneous organ, and different imaging modalities are known to capture different aspects of the naturally occurring variation. Thus, studies relying on other modalities, either independently or in combination, could reveal a broader set of associations^77^.

In conclusion, the present study increases the yield of genetic associations with brain age to eight genomic loci; implicated genes indicate involvement of calcium signaling, DNA damage repair, protein metabolism, and general innate immune defense. Our analysis did not provide evidence of a causal relationship between BAG and the included clinical conditions, and their interactions remain unclear.

## Data Availability

All data used in the study are from openly accessible datasets. Requests for the individual dataset have to be placed with the individual principal investigators

## Author contributions

EL, TW, and YW designed this study; EL, DVP, OF, AAS, OI, AMG and YW performed data analysis; EL, DVP, JMR, OI, TK, LTW, OOA, TW, BT, SMS, and YW interpreted the results; EL, DVP, JMR, OI, YW prepared the first draft of the manuscripts; All authors contributed and approved the final draft.

## Acknowledgement

This research has been conducted using the UK Biobank Resource under Application Number 27412. We thank the computational resources provided by UNINETT Sigma2-the National Infrastructure for High Performance Computing and Data Storage in Norway – with project no. (nn9769k/ns9769k), the PGC consortium and the International Parkinson Disease Genomics Consortium (IPDGC) for sharing of their GWAS results.

