## Supplemental Materials for "Genetic architecture of brain age and its casual relations with brain and mental disorders" (from another version of this preprint: version 1, 2023-01-11)

Leonardsen et al.

[Table S6. Harmonized instrumental SNPs (p<5x10-8) for the BAG to SCZ MR.	26](#_Toc1950128100)

[Table S7. Harmonized instrumental SNPs (p<5x10-8) for the BAG to BIP MR.	27](#_Toc1056618326)

[Table S8. Harmonized instrumental SNPs (p<5x10-8) for the BAG to MDD MR.	28](#_Toc364763797)

[Table S9. Harmonized instrumental SNPs (p<5x10-8) for the BAG to AD MR.	29](#_Toc1111985032)

[Table S10. Harmonized instrumental SNPs (p<5x10-8) for the BAG to PD MR.	30](#_Toc300439400)

[Table S11. Harmonized instrumental SNPs (p<5x10-8) for the SCZ to BAG MR.	31](#_Toc822513410)

[Table S12. Harmonized instrumental SNPs (p<5x10-8) for the BIP to BAG MR.	40](#_Toc1949864035)

[Table S13. Harmonized instrumental SNPs (p<5x10-8) for the MDD to BAG MR.	43](#_Toc874094663)

[Table S14. Harmonized instrumental SNPs (p<5x10-8) for the AD to BAG MR.	44](#_Toc14155446)

[Table S15. Harmonized instrumental SNPs (p<5x10-8) for the PD to BAG MR.	46](#_Toc1323970663)

**Replication analysis**

Genetic data

We attempted to replicate the eight associations to BAG using three independent datasets, the Alzheimer’s Disease Neuroimaging Initiative phase 1 (ADNI I) and 2 (ADNI II) and a healthy control dataset (Local sample) that have been previously published in Ripke et al^1^ . Out local sample has been imputed by psychiatry genomic consortium using their standardized pipeline Ricopili^2^. However, for the two ADNI datasets, only genotyped data are publicly available. We imputed these genotypes to the 1000 Genomes Projects reference data, using Genipe^3^. The default parameters of Genipe were used. After imputation, we excluded SNPs having MAF < 0.01, HWE p <10-6, and imputation R-sq <0.3 from subsequent analysis.

Image data

MRI data for ANDI sample were obtained from <http://adni.loni.usc.edu/> with permission. Brain scans for our local sample were performed at Oslo University Hospital and published in previous works^4-6^. The same protocols of image preprocessing and BAG estimation used in discovery sample were applied to these replication sample.

Replication association studies

We performed association test for each of the eight SNPs in each of the three replication datasets separately. For each analysis, chronological age, sex and the top ten principal components were included as covariates. Association results are shown in Supplementary Table S4.

**Supplementary Acknowledgement**

We thank the ADNI consortium for making their data publicly accessible. The ADNI was launched in 2003 as a public-private partnership, led by Principal Investigator Michael W. Weiner, MD. ADNI consists of 4 waves, the later is still ongoing (ADNI 3). A complete listing of ADNI investigators can be found at http://adni.loni.usc.edu/wp-content/uploads/how_to_apply/ADNI_Acknowledgement_List.pdf. Data collection and sharing for this project was funded by the ADNI (NIH Grant U01 AG024904) and DOD ADNI (Department of Defense award number W81XWH-12-2-0012). ADNI is funded by the National Institute on Aging, the National Institute of Biomedical Imaging and Bioengineering, and through generous contributions from the following: AbbVie, Alzheimer’s Association; Alzheimer’s Drug Discovery Foundation; Araclon Biotech; BioClinica, Inc.; Biogen; Bristol-Myers Squibb Company; CereSpir, Inc.; Cogstate Eisai Inc.; Elan Pharmaceuticals, Inc.; Eli Lilly and Company; EuroImmun; F. Hoffmann-La Roche Ltd and its affiliated company Genentech, Inc.; Fujirebio; GE Healthcare; IXICO Ltd.; Janssen Alzheimer Immunotherapy Research & Development, LLC.; Johnson & Johnson Pharmaceutical Research & Development LLC.; Lumosity; Lundbeck; Merck & Co., Inc.; Meso Scale Diagnostics, LLC.; NeuroRx Research; Neurotrack Technologies; Novartis Pharmaceuticals Corporation; Pfizer Inc.; Piramal Imaging; Servier; Takeda Pharmaceutical Company; and Transition Therapeutics. The Canadian Institutes of Health Research is providing funds to support ADNI clinical sites in Canada and the Saguenay Yout Study. Private sector contributions are facilitated by the Foundation for the National Institutes of Health (http://www.fnih.org). The grantee organization is the Northern California Institute for Research and Education, and the study is coordinated by the Alzheimer’s Therapeutic Research Institute at the University of Southern California. ADNI data are disseminated by the Laboratory for Neuro Imaging at the University of Southern California.

### **Figure S1. Genetic association to brain age gap in each fold.**

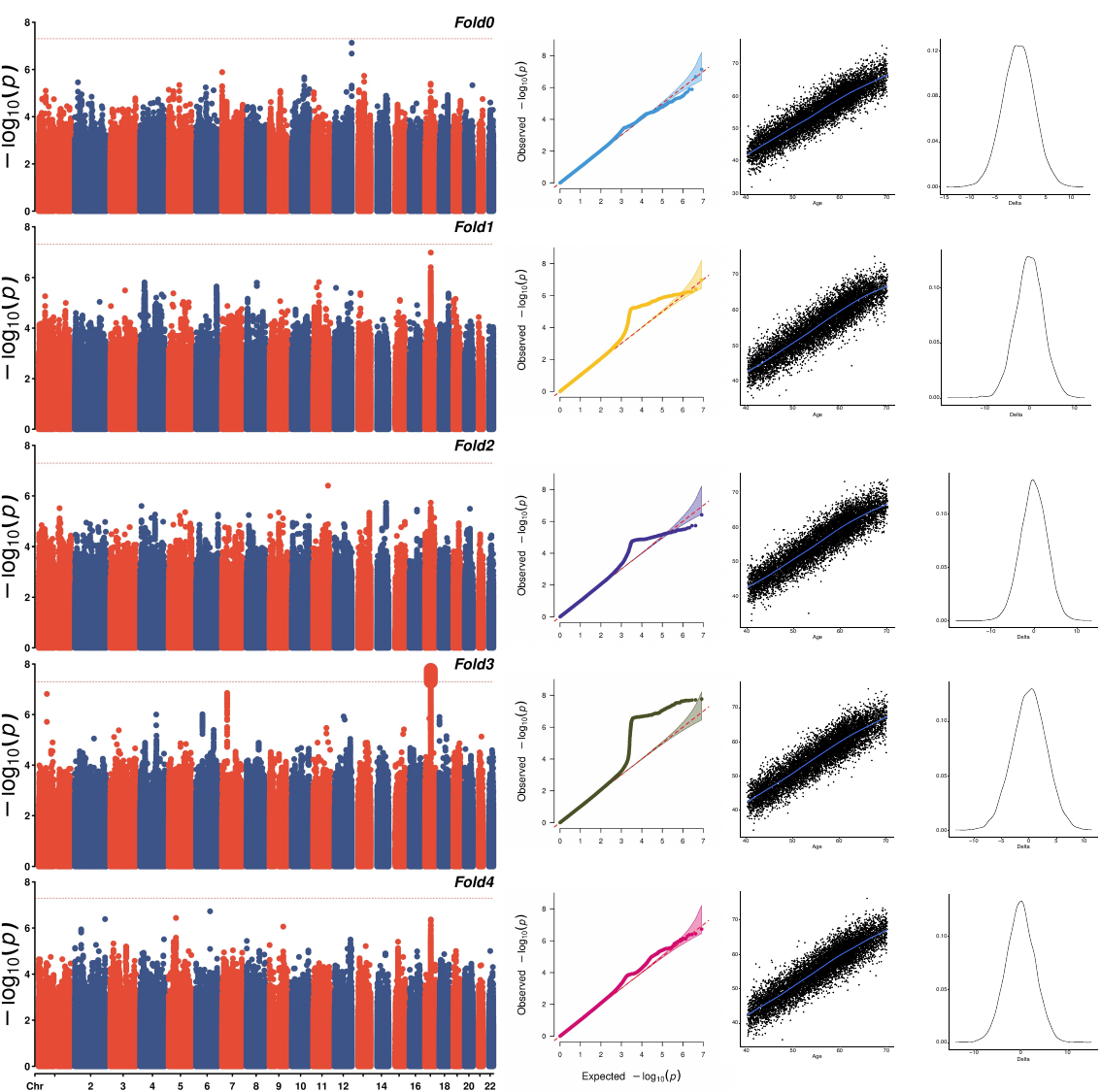

From left to right, GWAS association Manhattan plot and QQ plot, the scatter plot for estimated brain age (y axis) vs. chronological age (x axis), distribution for brain age gap (delta) for each fold of the UKB data (each row). In Manhattan plots, chromosome numbers are on x axes and -log10 association p value on the y axes. In QQ plots, the expected and observed p value on the -log10 scale are shown on the x and y axes. Blue lines in the scatter plots indicated a fitted loess model for each fold.

### **Figure S2. QQ plot for the genetic association to brain age gap.**

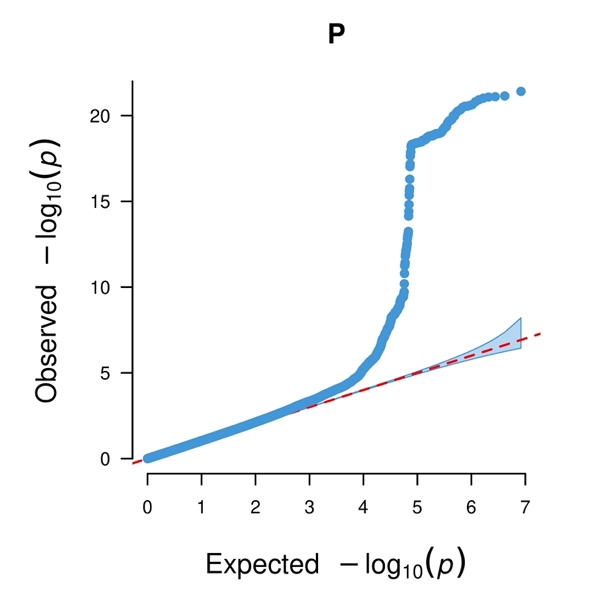

QQ plot for the results of meta-analysis over the GWAS for BAG in each of the five folds of UKB data.

### **Figure S3. Genic annotations for associated SNPs.**

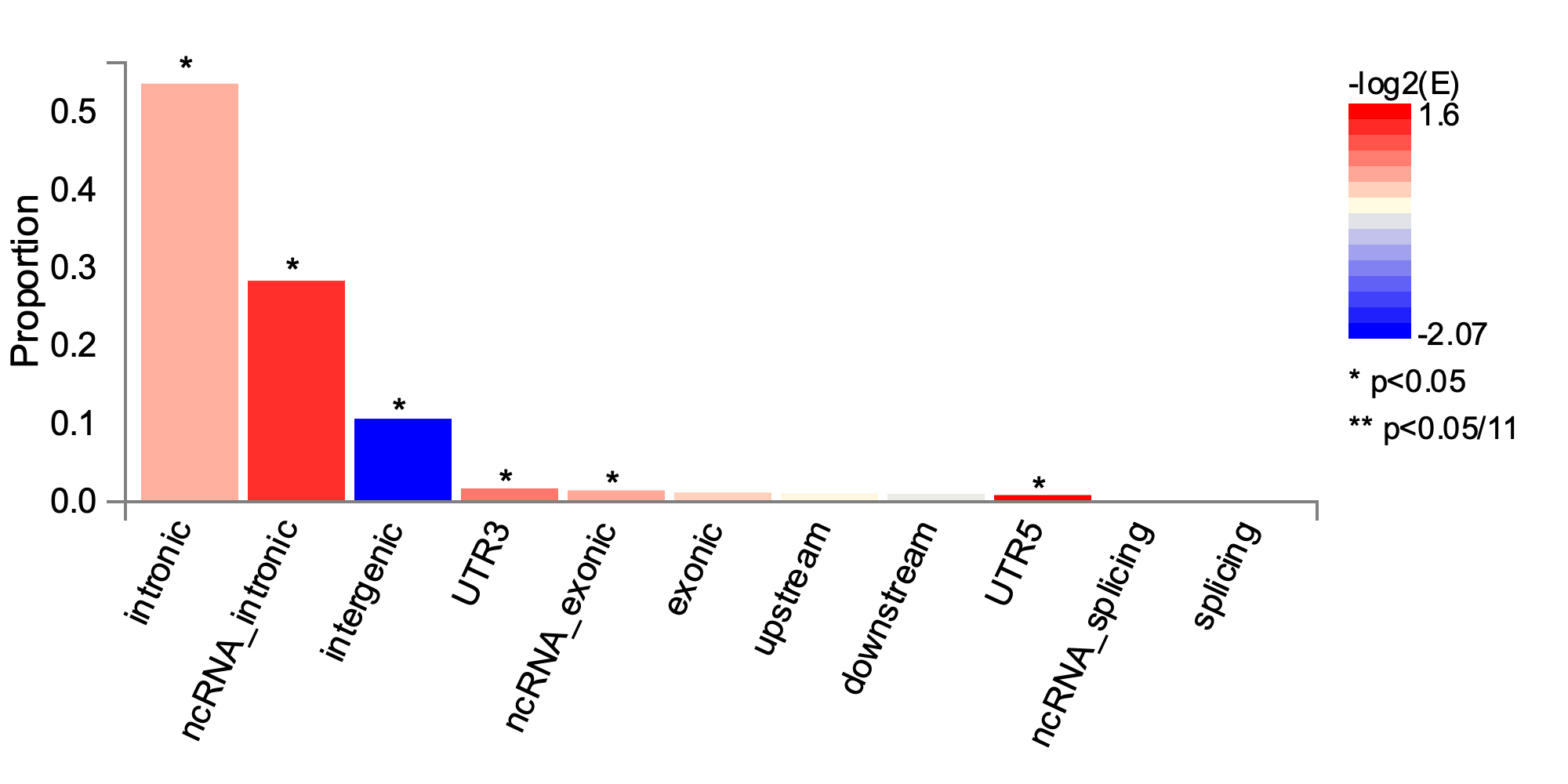

Annotations of associated SNPs to genic elements by FUMA^7^. UTR3, 3 prime untranslated region; ncRNA, non-coding RNA; UTR5, 5 prime untranslated region. Enrichment p values are based on hypergeometric test.

### **Figure S4. Annotations for associated SNPs.**

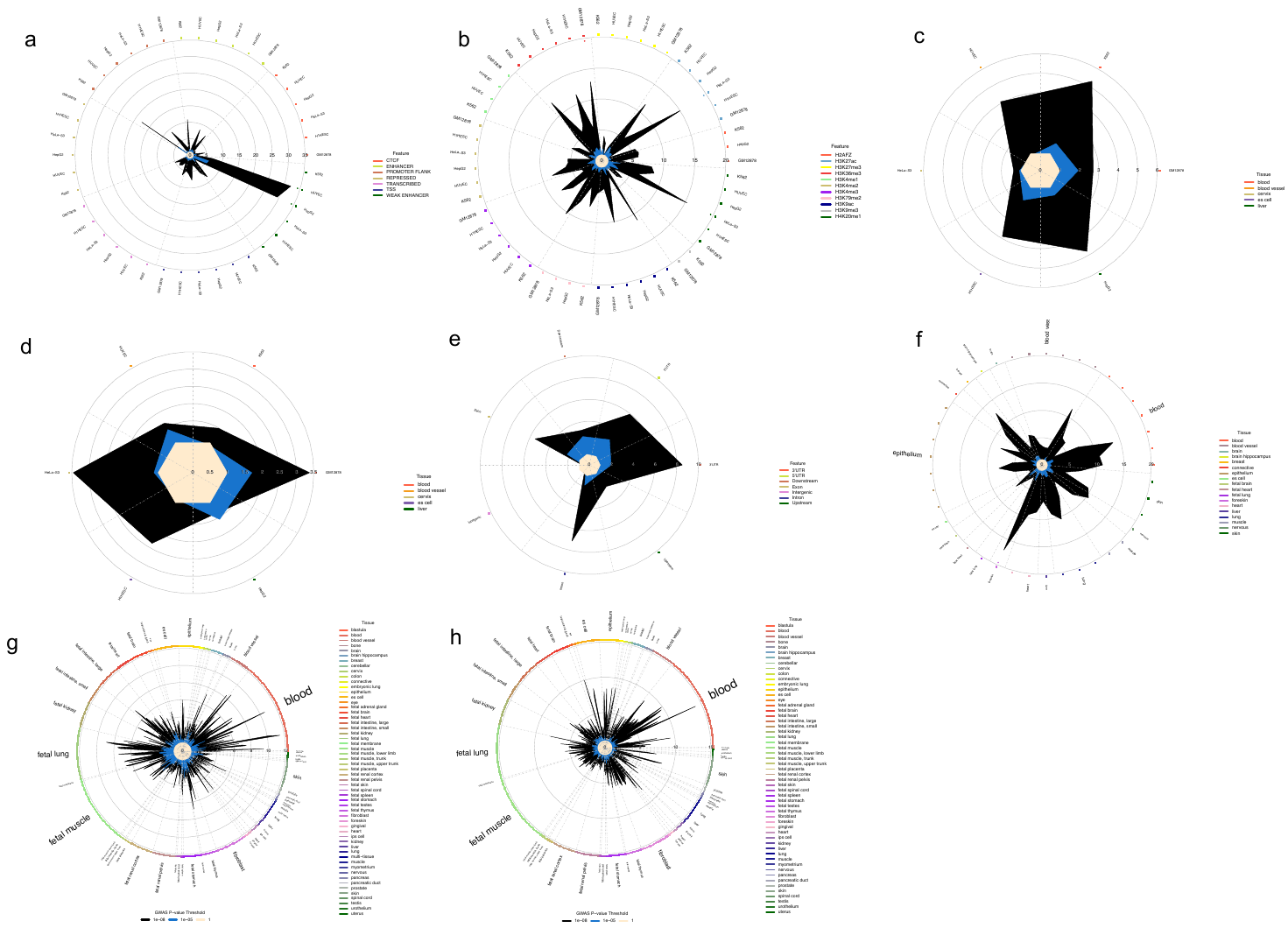

Annotations of associated SNPs to genic elements by Garfield^8^.

### **Figure S5. Gene based association results.**

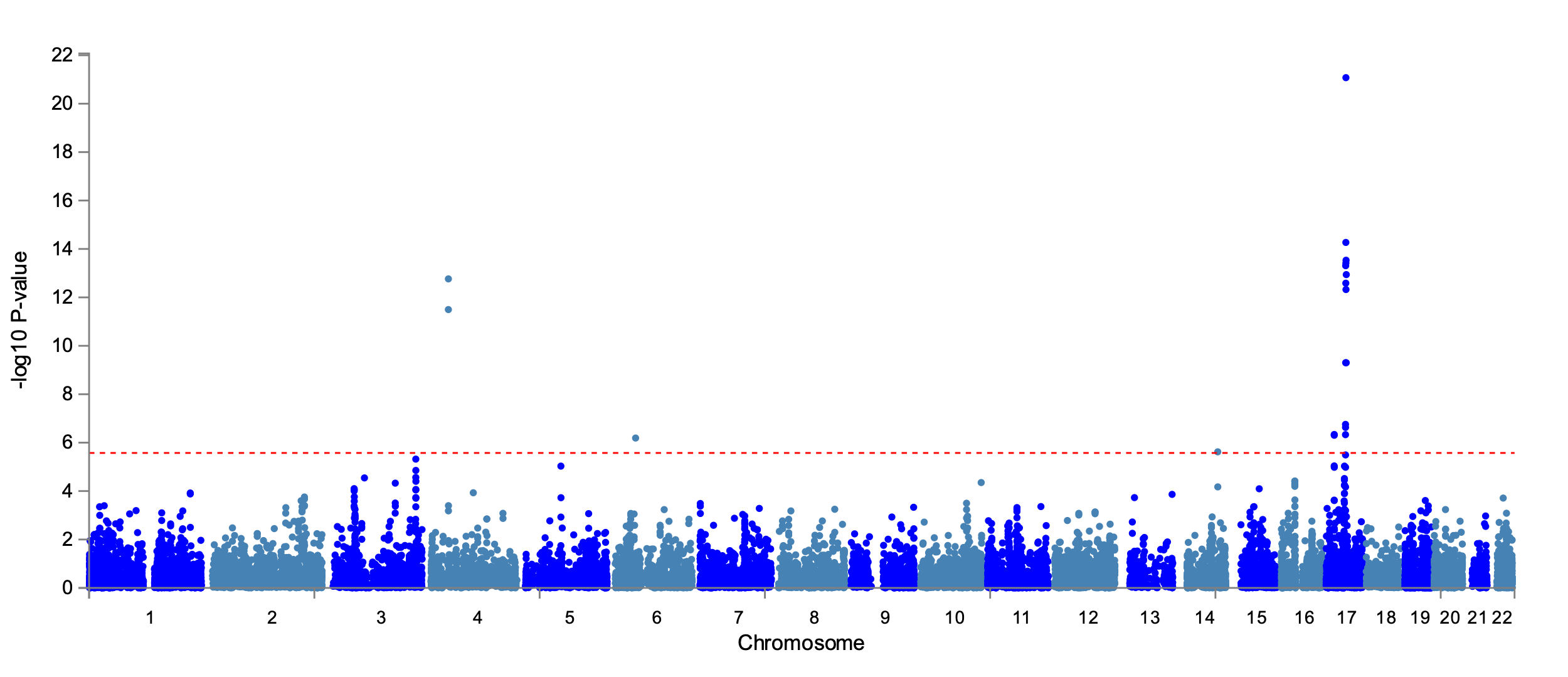

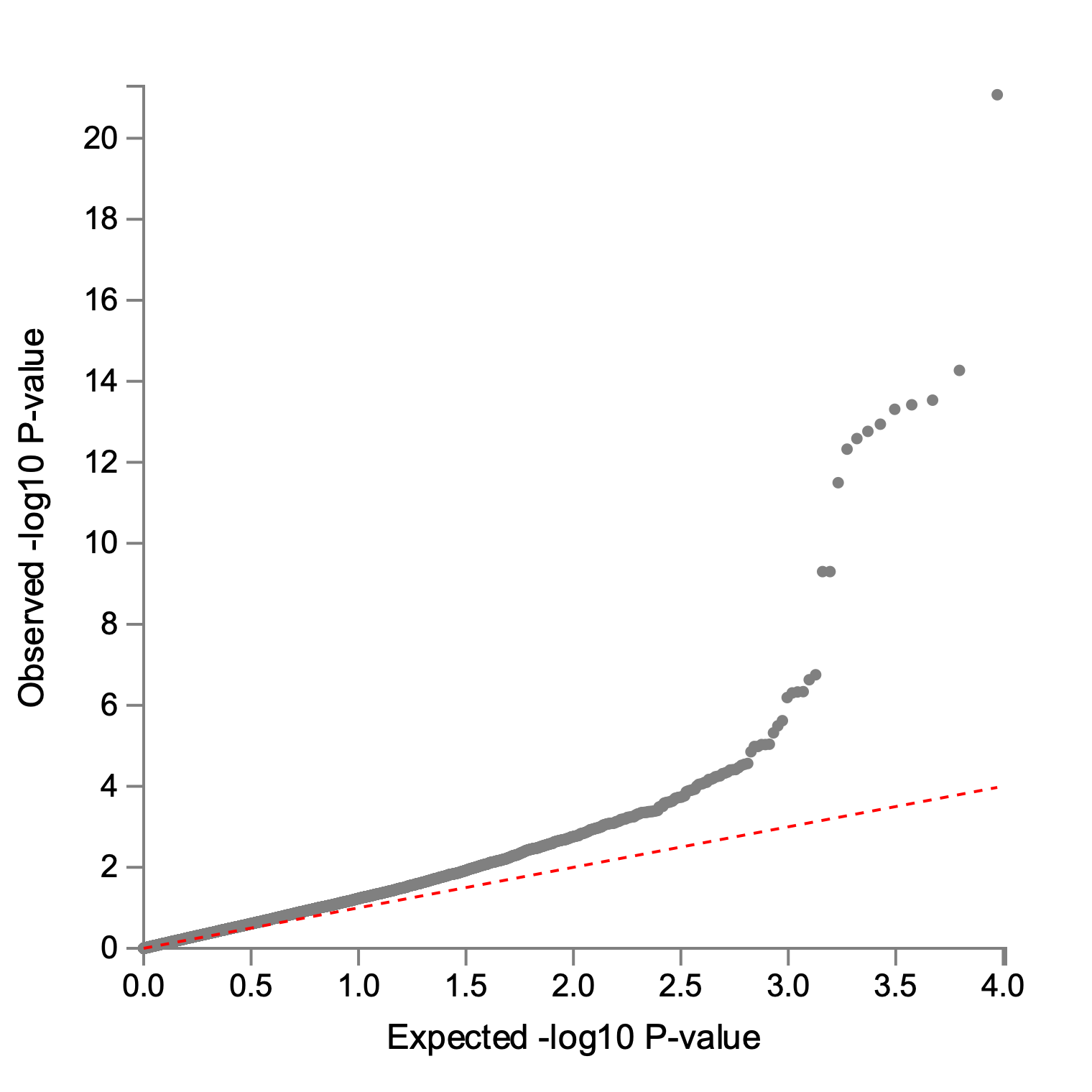

Manhattan plot and QQ plot for gene-based association statistics for BAG. Dashed line in the Manhattan plot indicates the genome wide significant threshold, and, in the QQ plot indicates the null association.

### **Figure S6. Conditional QQ plot for MDD vs BAG.**

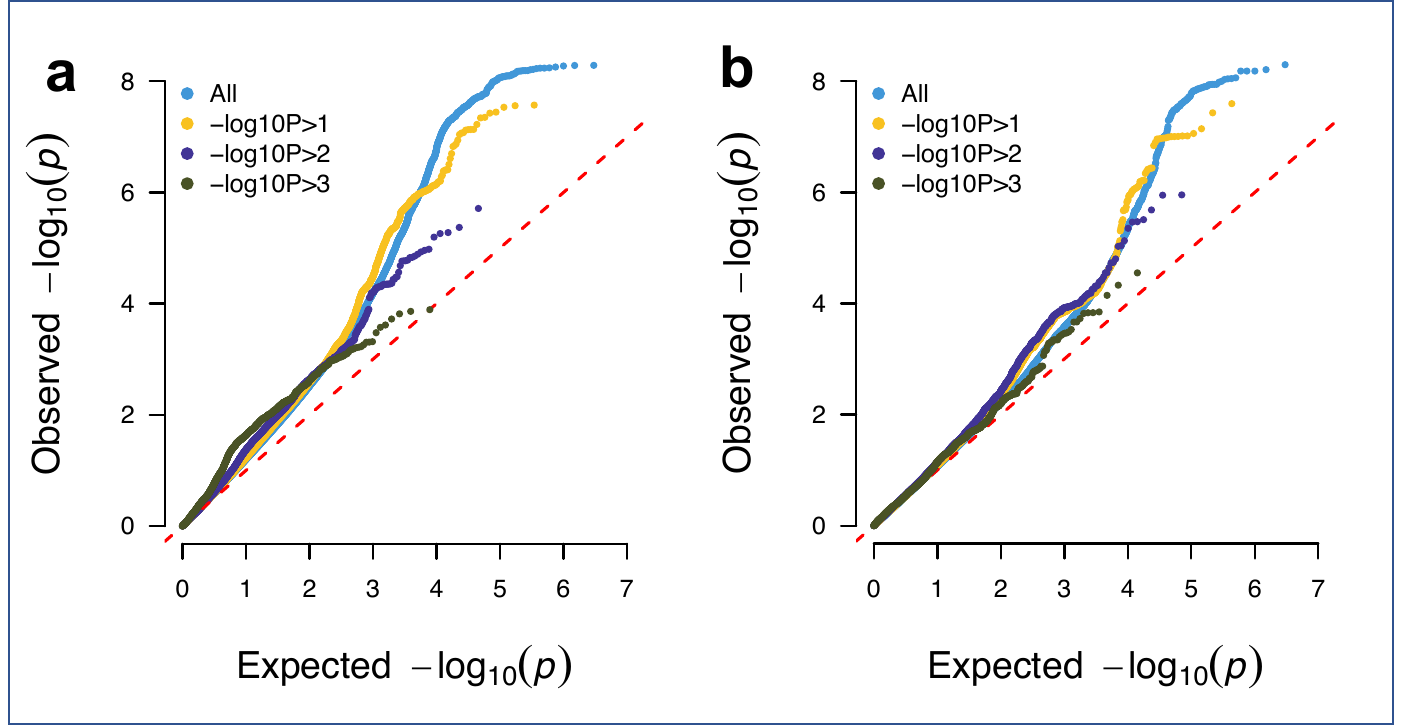

The conditional QQ plots for MDD condition on the levels of association for BAG (**a**), and for BAG condition on the level of association for MDD (**b**). As the association strength to the conditioned disorder increase, a successive leftward deflation in these curves indicates polygenic enrichment.

### **Figure S7. Conditional QQ plot for SCZ vs BAG.**

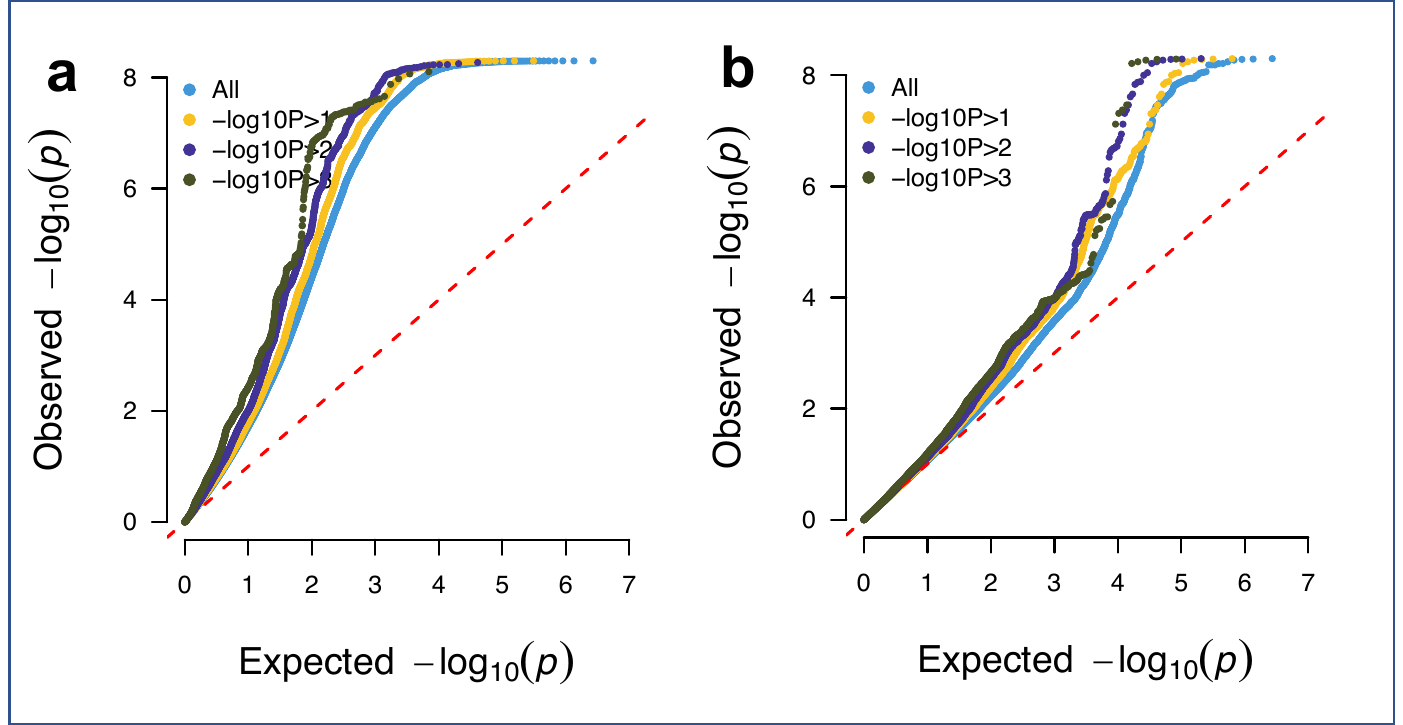

The conditional QQ plots for SCZ condition on the levels of association for BAG (**a**), and for BAG condition on the level of association for SCZ (**b**). As the association strength to the conditioned disorder increase, a successive leftward deflation in these curves indicates polygenic enrichment.

### **Figure S8. Conditional QQ plot for PD vs BAG excluding chr17.**

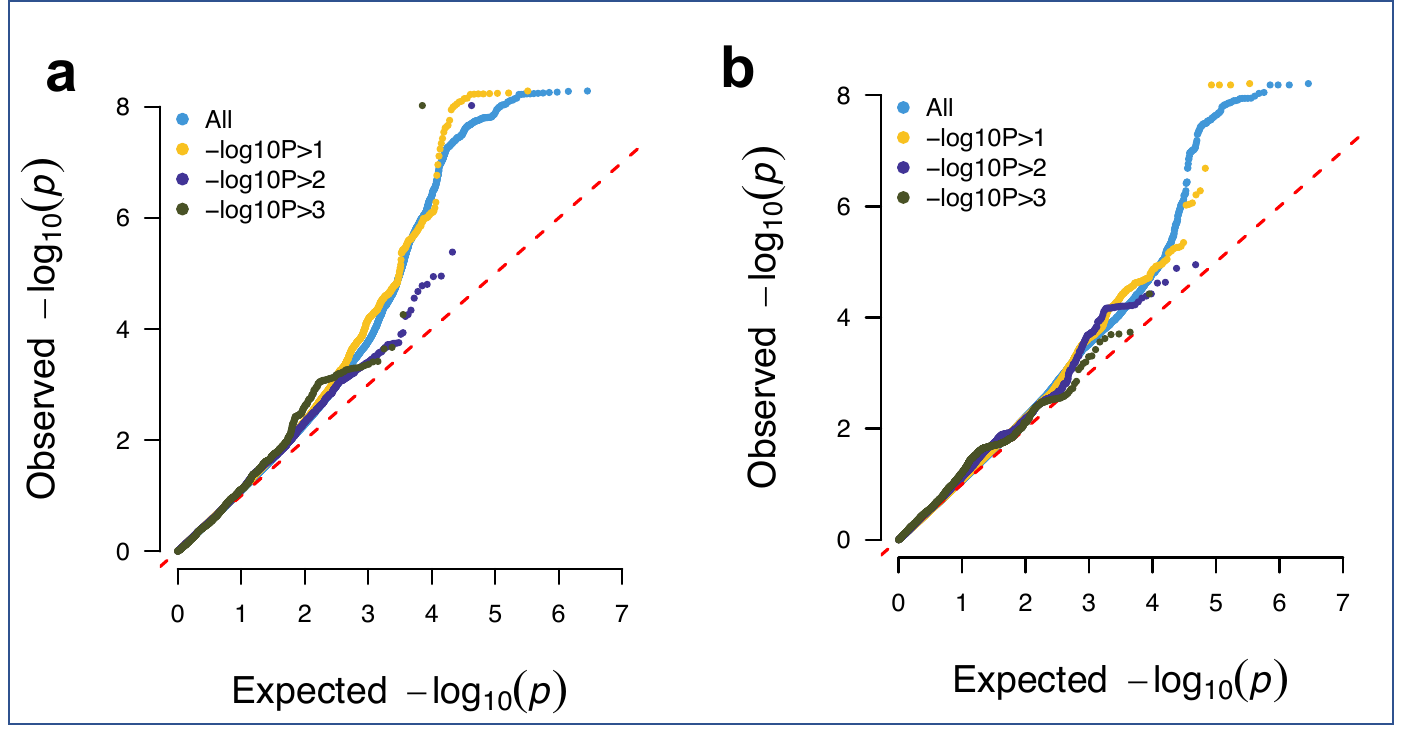

The conditional QQ plots for PD condition on the levels of association for BAG (**a**), and for BAG condition on the level of association for PD (**b**). As the association strength to the conditioned disorder increase, a successive leftward deflation in these curves indicates polygenic enrichment.

### **Figure S9. Predictive performance of the brain age model**

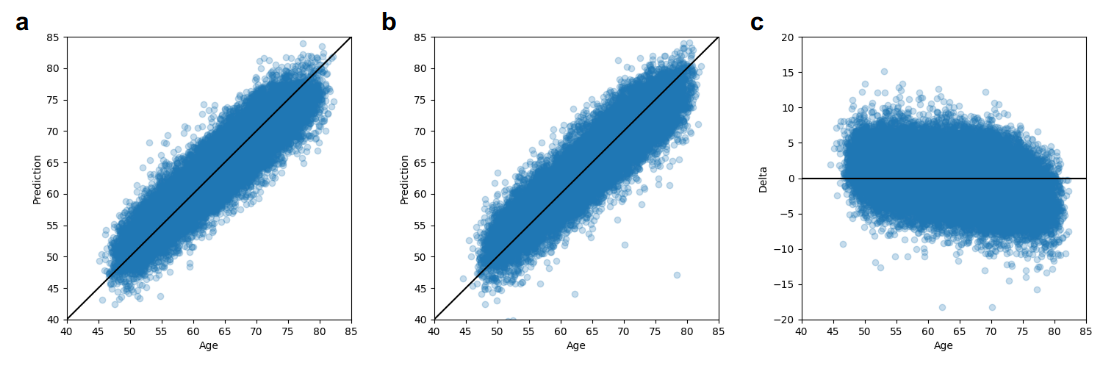

Predictive performance of the brain age models in the females (a) and males (b) of UKB separately. (c) shows the brain age gap as a function of age

### **Table S1. Non-UK Biobank model training datasets.**

| **Name** | **Size** | **Subset** | **Ages** | **Train** |
| --- | --- | --- | --- | --- |
| Autism Brain Imaging Data Exchange | 555 | Healthy Controls | 6-56 | 356 |
| Autism Brain Imaging Data Exchange II | 184 | Healthy Controls | 8-64 | 118 |
| ADHD 200 | 606 | Healthy Controls | 7-26 | 388 |
| Amsterdam Open MRI Collection | 819 | ID1000 | 19-26 | 524 |
| Beijing Normal University Enhanced Sample | 180 | Full | 17-28 | 116 |
| Cambridge Center for Aging and Neuroscience | 653 | Full | 18-89 | 418 |
| Consortium for Reliability and Reproducibility | 1368 | Full | 6-84 | 880 |
| Dallas Lifespan Brain Study | 314 | Full | 21-89 | 201 |
| ds000119 | 73 | Full | 8-27 | 47 |
| ds000202 | 95 | Full | 18-30 | 61 |
| ds000222 | 79 | Full | 21-73 | 51 |
| 1000 Functional Connectomes Project | 812 | Full | 8-78 | 520 |
| Healthy Brain Network | 1855 | Full | 5-22 | 1188 |
| Human Connectome Project | 1113 | Full | 22-37 | 712 |
| Max Planck Institute Leipzig Mind-Brain-Body | 73 | Full | 22-68 | 47 |
| Enhanced Nathan Kline Institute - Rockland Sample | 928 | Full | 6-85 | 594 |
| Open Access Series of Imaging Studies 3 | 1264 | Healthy Controls | 42-95 | 817 |
| Pediatric, Imaging and Neurocognition | 1174 | Full | 3-21 | 752 |
| Southwest University Adult Lifespan Dataset | 494 | Full | 19-80 | 316 |
| Southwest University Longitudinal Imaging Multimodal | 573 | Full | 17-27 | 367 |
| **Total** | **13212** | **-** | **3-95** | **8473** |

The dataset was compiled from 20 sources, spanning a wide age range and a multitude of scanners and scanning protocols. The number of participants from each source used for training is shown in the respective columns.

### Table S2. Characteristics for the five folds data from UK Biobank**.**

| Index | N | Age range (years) | Sex (F) | MAE(years) |
| --- | --- | --- | --- | --- |
| Fold 0 | 5622 | 40-70 | 2907 | 2.49 |
| Fold 1 | 5601 | 40-70 | 2885 | 2.44 |
| Fold 2 | 5601 | 40-70 | 2888 | 2.45 |
| Fold 3 | 5650 | 40-70 | 2914 | 2.44 |
| Fold 4 | 5630 | 40-70 | 2907 | 2.44 |

Four folds were used for model training and tuning, and the trained model was used for predicting brain age for the hold-off fold. This procedure was iterated five times until all subjects have a brain age estimated from independent training models. Indexes corresponding to those used in Supplementary Figure 1; N, sample size; MAE, mean absolute error for predicted brain age.

### Table S3. Statistics for lead SNPs of each associated region in each fold.

| Fold | SNP | BP | A1 | t-score | P |
| --- | --- | --- | --- | --- | --- |
| fold0 | rs2790102 | 45432214 | A | -2.54 | 1.12e-2 |
| fold1 | rs2790102 | 45432214 | A | -1.95 | 5.09e-2 |
| fold2 | rs2790102 | 45432214 | A | -3.56 | 3.77e-4 |
| fold3 | rs2790102 | 45432214 | A | -1.86 | 6.36e-2 |
| fold4 | rs2790102 | 45432214 | A | -2.97 | 2.98e-3 |
| fold0 | rs4880424 | 134584577 | T | 3.76 | 1.7e-4 |
| fold1 | rs4880424 | 134584577 | T | 3.16 | 1.61e-3 |
| fold2 | rs4880424 | 134584577 | T | 3.12 | 1.79e-3 |
| fold3 | rs4880424 | 134584577 | T | -0.54 | 0.59 |
| fold4 | rs4880424 | 134584577 | T | 2.75 | 5.97e-3 |
| fold0 | rs17203398 | 88449847 | C | -4.13 | 3.71e-5 |
| fold1 | rs17203398 | 88449847 | C | -1.37 | 0.17 |
| fold2 | rs17203398 | 88449847 | C | -4.35 | 1.41e-5 |
| fold3 | rs17203398 | 88449847 | C | -2.04 | 4.19e-2 |
| fold4 | rs17203398 | 88449847 | C | -2.44 | 1.47e-2 |
| fold0 | rs2106786 | 43919096 | G | 3.72 | 2.05e-4 |
| fold1 | rs2106786 | 43919096 | G | 4.46 | 8.42e-6 |
| fold2 | rs2106786 | 43919096 | G | 4.45 | 8.59e-6 |
| fold3 | rs2106786 | 43919096 | G | 5.50 | 4.01e-8 |
| fold4 | rs2106786 | 43919096 | G | 4.22 | 2.52e-5 |
| fold0 | rs73185796 | 183975709 | T | -1.21 | 0.23 |
| fold1 | rs73185796 | 183975709 | T | -2.84 | 4.52e-3 |
| fold2 | rs73185796 | 183975709 | T | -3.39 | 6.93e-4 |
| fold3 | rs73185796 | 183975709 | T | -2.32 | 2.02e-2 |
| fold4 | rs73185796 | 183975709 | T | -2.72 | 6.57e-3 |
| fold0 | rs13132853 | 38680015 | G | 3.59 | 3.32e-4 |
| fold1 | rs13132853 | 38680015 | G | 4.18 | 2.96e-5 |
| fold2 | rs13132853 | 38680015 | G | 4.29 | 1.82e-5 |
| fold3 | rs13132853 | 38680015 | G | 4.10 | 4.16e-5 |
| fold4 | rs13132853 | 38680015 | G | 3.378 | 7.35e-4 |
| fold0 | rs79107704 | 78388694 | A | 2.26 | 2.38e-2 |
| fold1 | rs79107704 | 78388694 | A | 1.64 | 0.10 |
| fold2 | rs79107704 | 78388694 | A | 3.27 | 1.1e-3 |
| fold3 | rs79107704 | 78388694 | A | 2.92 | 3.57e-3 |
| fold4 | rs79107704 | 78388694 | A | 2.49 | 1.29e-2 |
| fold0 | rs7461069 | 124669029 | A | -3.10 | 1.94e-3 |
| fold1 | rs7461069 | 124669029 | A | -2.64 | 8.37e-3 |
| fold2 | rs7461069 | 124669029 | A | -1.47 | 0.14 |
| fold3 | rs7461069 | 124669029 | A | -2.04 | 4.13e-2 |
| fold4 | rs7461069 | 124669029 | A | -3.39 | 7.12e-4 |

BP: base pair position on the hg19 build; A1: effective allele; t-score: effect size divided by standard error; P: association p values. Numeric values were rounded to two decimal places.

### Table S4. Statistics for lead SNPs of each associated region in each replication samples.

**a.**

| CHR | BP | SNP | A1 | N | t-score | P |
| --- | --- | --- | --- | --- | --- | --- |
| 4 | 38680015 | rs13132853 | G | 702 | 0.26 | 0.79 |
| 6 | 45432214 | rs2790102 | A | 702 | -0.13 | 0.90 |
| 8 | 124669029 | rs7461069 | A | 702 | -2.30 | 2.19e-2 |
| 10 | 134584577 | rs4880424 | T | 702 | -0.72 | 0.47 |
| 14 | 88449847 | rs17203398 | C | 702 | -2.70 | 7.17e-3 |
| 17 | 43919096 | rs2106786 | G | 702 | 0.86 | 0.39 |

**b.**

| CHR | BP | SNP | A1 | N | t-score | P |
| --- | --- | --- | --- | --- | --- | --- |
| 3 | 183975709 | rs73185796 | T | 321 | 0.88 | 0.38 |
| 4 | 38680015 | rs13132853 | G | 321 | -0.44 | 0.66 |
| 6 | 45432214 | rs2790102 | A | 321 | -1.06 | 0.29 |
| 8 | 124669029 | rs7461069 | A | 321 | 0.19 | 0.85 |
| 10 | 134584577 | rs4880424 | T | 321 | -0.19 | 0.85 |
| 14 | 88449847 | rs17203398 | C | 321 | 0.16 | 0.87 |
| 17 | 43919096 | rs2106786 | G | 321 | 0.52 | 0.60 |

**c.**

| CHR | BP | SNP | A1 | N | t-score | P |
| --- | --- | --- | --- | --- | --- | --- |
| 3 | 183975709 | rs73185796 | T | 608 | -1.73 | 8.50e-2 |
| 4 | 38680015 | rs13132853 | G | 600 | -0.44 | 0.66 |
| 5 | 78388694 | rs79107704 | A | 612 | 1.11 | 0.27 |
| 6 | 45432214 | rs2790102 | A | 501 | -1.22 | 0.22 |
| 8 | 124669029 | rs7461069 | A | 595 | 1.004 | 0.32 |
| 10 | 134584577 | rs4880424 | T | 602 | 0.35 | 0.72 |
| 14 | 88449847 | rs17203398 | C | 611 | 0.66 | 0.51 |

**a.** ADNI I sample; **b.** ADNI II sample; **c.** Local healthy control sample.

The ADNI samples I and II were impudently imputed on two the 1000 Genomes Project Phases3 reference data using the software Genipe^3^.

CHR: chromosome number; BP: base pair position on the hg19 build; A1: effective allele; N, number of subjects analyzed; t-score: effect size divided by standard error; P: association p values. Numeric values were rounded to two decimal places.

### Table S5 Function annotation for genes in Figure 1c.

1. Symbol: **BHMT2**; Name: **Betaine--Homocysteine S-Methyltransferase 2**

Homocysteine is a sulfur-containing amino acid that plays a crucial role in methylation reactions. Transfer of the methyl group from betaine to homocysteine creates methionine, which donates the methyl group to methylate DNA, proteins, lipids, and other intracellular metabolites. The protein encoded by this gene is one of two methyl transferases that can catalyze the transfer of the methyl group from betaine to homocysteine. Anomalies in homocysteine metabolism have been implicated in disorders ranging from vascular disease to neural tube birth defects such as spina bifida, Orofacial Cleft. Among its related pathways are Metabolism and Sulfur amino acid metabolism.

2. Symbol: **AP2M1**; Name: **Adaptor Related Protein Complex 2 Subunit Mu 1**

This gene encodes a subunit of the heterotetrameric coat assembly protein complex 2 (AP2), which belongs to the adaptor complexes medium subunits family. The encoded protein is required for the activity of a vacuolar ATPase, which is responsible for proton pumping occurring in the acidification of endosomes and lysosomes. The encoded protein may also play an important role in regulating the intracellular trafficking and function of CTLA-4 protein. Among its related pathways are Arf1 pathway and Metabolism. Adaptor protein complexes function in protein transport via transport vesicles in different membrane traffic pathways. AP-2 is involved in clathrin-dependent endocytosis in which cargo proteins are incorporated into vesicles surrounded by clathrin (clathrin-coated vesicles, CCVs) which are destined for fusion with the early endosome. Clathrin-associated adaptor protein (AP) complexes which can bind directly to both the clathrin lattice and to the lipid and protein components of membranes are considered to be the major clathrin adaptors contributing the CCV formation. AP-2 may also play a role in maintaining normal post-endocytic trafficking through the ARF6-regulated, non-clathrin pathway. During long-term potentiation in hippocampal neurons, AP-2 is responsible for the endocytosis of ADAM10. Diseases associated with AP2M1 include *Intellectual Developmental Disorder*, *Autosomal Dominant 60*, *Seizures* and *Epilepsy*, *Myoclonic-Atonic Seizures*.

3. Symbol: **PSMD2**; Name: **Proteasome 26S Subunit Ubiquitin Receptor, Non-ATPase 2**

The 26S proteasome is a multicatalytic proteinase complex with a highly ordered structure composed of 2 complexes, a 20S core and a 19S regulator. Proteasomes are distributed throughout eukaryotic cells at a high concentration and cleave peptides in an ATP/ubiquitin-dependent process in a non-lysosomal pathway. An essential function of a modified proteasome, the immunoproteasome, is the processing of class I MHC peptides. This gene encodes one of the non-ATPase subunits of the 19S regulator lid. In addition to participation in proteasome function, this subunit may also participate in the TNF signalling pathway since it interacts with the tumor necrosis factor type 1 receptor. This complex plays a key role in the maintenance of protein homeostasis by removing misfolded or damaged proteins, which could impair cellular functions, and by removing proteins whose functions are no longer required. Therefore, the proteasome participates in numerous cellular processes, including cell cycle progression, apoptosis, or DNA damage repair.

4. Symbol: **HEXIM1**; Name: **HEXIM P-TEFb Complex Subunit 1**

Expression of this gene is induced by hexamethylene-bis-acetamide in vascular smooth muscle cells. Among its related pathways are Initiation of transcription and translation elongation at the HIV-1 LTR and Chromatin Regulation/Acetylation. It is a transcriptional regulator which functions as a general RNA polymerase II transcription inhibitor. Core component of the 7SK RNP complex: in cooperation with 7SK snRNA sequesters P-TEFb in a large inactive 7SK snRNP complex preventing RNA polymerase II phosphorylation and subsequent transcriptional elongation. It may also regulate NF-kappa-B, ESR1, NR3C1 and CIITA-dependent transcriptional and plays a role in the regulation of DNA virus-mediated innate immune response by assembling into the HDP-RNP complex, a complex that serves as a platform for IRF3 phosphorylation and subsequent innate immune response activation through the cGAS-STING pathway. Diseases associated with HEXIM1 include *Nut Midline Carcinoma* and *Immune Deficiency Disease*.

5. Symbol: **EIF2B5**; Name: **Eukaryotic Translation Initiation Factor 2B Subunit Epsilon**

This gene encodes one of five subunits of eukaryotic translation initiation factor 2B (EIF2B), a GTP exchange factor for eukaryotic initiation factor 2 and an essential regulator for protein synthesis. Among its related pathways are Translational Control and Peptide chain elongation. It catalyzes the exchange of eukaryotic initiation factor 2-bound GDP for GTP. Mutations in this gene and the genes encoding other EIF2B subunits have been associated with *leukoencephalopathy* with vanishing white matter.

6. Symbol: **NMT1**; Name: **N-Myristoyltransferase 1**

Myristate, a rare 14-carbon saturated fatty acid, is cotranslationally attached by an amide linkage to the N-terminal glycine residue of cellular and viral proteins with diverse functions. N-myristoyltransferase catalyzes the transfer of myristate from CoA to proteins. N-myristoylation appears to be irreversible and is required for full expression of the biologic activities of several N-myristoylated proteins, including the alpha subunit of the signal-transducing guanine nucleotide-binding protein (G protein). Among its related pathways are Activation of BH3-only proteins and HIV Life Cycle. It adds a myristoyl group to the N-terminal glycine residue of certain cellular and viral proteins. Diseases associated with NMT1 include *Gallbladder Cancer* and *Noonan Syndrome 11*.

7. Symbol: **DVL3**; Name: **Dishevelled Segment Polarity Protein 3**

This gene is a member of a multi-gene family which shares strong similarity with the Drosophila dishevelled gene, dsh. The Drosophila dishevelled gene encodes a cytoplasmic phosphoprotein that regulates cell proliferation. It also Involved in the signal transduction pathway mediated by multiple Wnt genes.

8. Symbol: **PARL**; Name: **Presenilin Associated Rhomboid Like**

This gene encodes a member of the rhomboid family of intramembrane serine proteases that is localized to the inner mitochondrial membrane. The encoded protein regulates mitochondrial remodeling and apoptosis through regulated substrate proteolysis. Proteolytic processing of the encoded protein results in the release of a small peptide, P-beta, which may transit to the nucleus. This protein is required for the control of apoptosis during postnatal growth and promotes changes in mitochondria morphology regulated by phosphorylation of P-beta domain. Mutations in this gene may be associated with *Parkinson's disease*.

9. Symbol: **DCAKD**; Name: **Dephospho-CoA Kinase Domain Containing**

The protein coded by this gene is predicted to enable dephospho-CoA kinase activity and involved in coenzyme A biosynthetic process. Gene Ontology (GO) annotations related to this gene include dephospho-CoA kinase activity.

10. Symbol: **INPP5A**; Name: **Inositol Polyphosphate-5-Phosphatase A**

The protein encoded by this gene is a membrane-associated type I inositol 1,4,5-trisphosphate (InsP3) 5-phosphatase. InsP3 5-phosphatases hydrolyze Ins(1,4,5)P3, which mobilizes intracellular calcium and acts as a second messenger mediating cell responses to various stimulation. Among its related pathways are Metabolism and superpathway of D-myo-inositol (1,4,5)-trisphosphate metabolism. Diseases associated with INPP5A include *Alternating Esotropia* and *Lowe Oculocerebrorenal Syndrome*.

11. Symbol: **FAM114A1**; Name: **Family With Sequence Similarity 114 Member A1**

The protein encoded by this gene belongs to the FAM114 family and may play a role in *neuronal cell development*. It is a phosphatase that specifically hydrolyzes the 5-phosphate of inositol 1,4,5-trisphosphate to inositol 1,4-bisphosphate, and inositol 1,3,4,5-tetrasphosphate to inositol 1,3,4-trisphosphate. It also may play a crucial role in the survival of cerebellar Purkinje cells.

12. Symbol: **ALG3**; Name: **ALG3 Alpha-1,3- Mannosyltransferase**

This gene encodes a member of the ALG3 family. The encoded protein catalyses the addition of the first dol-P-Man derived mannose in an alpha 1,3 linkage to Man5GlcNAc2-PP-Dol. Defects in this gene have been associated with c*ongenital disorder of glycosylation type Id* (CDG-Id) characterized by abnormal N-glycosylation, and Congenital Disorders Of N-Linked Glycosylation. Among its related pathways are Synthesis of substrates in N-glycan biosythesis and Metabolism of proteins.

14. Symbol: **KLF3**; Name: **Kruppel Like Factor 3**

The protein coded by this gene enables sequence-specific double-stranded DNA binding activity. It is predicted to be involved in regulation of transcription by RNA polymerase II. The protein binds to the CACCC box of erythroid cell-expressed genes and may play a role in hematopoiesis.

15. Symbol: **ARHGAP27**; Name: **Rho GTPase Activating Protein 27**

This gene encodes a member of a large family of proteins that activate Rho-type guanosine triphosphate (GTP) metabolizing enzymes. The encoded protein may pay a role in clathrin-mediated endocytosis. GTPase activators for the Rho-type GTPases act by converting them to an inactive GDP-bound state.

16. Symbol: **ABCC5**; Name: **ATP Binding Cassette Subfamily C Member 5**

The protein encoded by this gene is a member of the superfamily of ATP-binding cassette (ABC) transporters. ABC proteins transport various molecules across extra- and intra-cellular membranes. This protein is a member of the MRP subfamily which is involved in multi-drug resistance. It functions in the cellular export of its substrate, cyclic nucleotides. This export contributes to the degradation of phosphodiesterases and possibly an elimination pathway for cyclic nucleotides. Studies show that this protein provides resistance to thiopurine anticancer drugs, 6-mercatopurine and thioguanine, and the *anti-HIV drug 9*-(2-phosphonylmethoxyethyl) adenine. Among its related pathways are Metabolism and Glycosaminoglycan metabolism. It acts as a heme transporter required for the translocation of cytosolic heme to the secretory pathway and may play a role in energy metabolism by regulating the glucagon-like peptide 1 (GLP-1) secretion from enteroendocrine cells. This protein may be involved in resistance to thiopurines in *acute lymphoblastic leukemia* and *antiretroviral* nucleoside analogs in HIV-infected patients. Diseases associated with ABCC5 include *Primary Angle-Closure Glaucoma* and *Episodic Kinesigenic Dyskinesia 1*.

17. Symbol: **NSF**; Name: **N-Ethylmaleimide Sensitive Factor, Vesicle Fusing ATPase**

The protein coded by this gene enables PDZ domain binding activity and ionotropic glutamate receptor binding activity. It is involved in intracellular protein transport, positive regulation of protein catabolic process, and positive regulation of receptor recycling. Among its related pathways are Neuroscience and Vesicle-mediated transport. Diseases associated with NSF include *Developmental And Epileptic Encephalopathy 96* and *Tetanus*.

18. Symbol: **FMNL1**; Name: **Formin Like 1**

This gene encodes a formin-related protein. Formin-related proteins have been implicated in morphogenesis, cytokinesis, and cell polarity. The protein may play a role in the control of cell motility, survival of macrophages, the regulation of cell morphology and cytoskeletal organization. It is required in the cortical actin filament dynamics and cell shape.

19. Symbol: **ACBD4**; Name: **Acyl-CoA Binding Domain Containing 4**

This gene encodes a member of the acyl-coenzyme A binding domain containing protein family. All family members contain the conserved acyl-Coenzyme A binding domain, which binds acyl-CoA thiol esters. They are thought to play roles in acyl-CoA dependent lipid metabolism by binding to medium- and long-chain acyl-CoA esters and may function as an intracellular carrier of acyl-CoA esters.

20. Symbol: **GALC**; Name: **Galactosylceramidase**

This gene encodes a lysosomal protein which hydrolyzes the galactose ester bonds of galactosylceramide, galactosylsphingosine, lactosylceramide, and monogalactosyldiglyceride. Enzyme with very low activity responsible for the lysosomal catabolism of galactosylceramide, a major lipid in myelin, kidney and epithelial cells of small intestine and colon. Mutations in this gene have been associated with *Krabbe disease*, also known as globoid cell leukodystrophy.

21. Symbol: **KANSL1**; Name: **KAT8 Regulatory NSL Complex Subunit 1**

This gene encodes a nuclear protein that is a subunit of two protein complexes involved with histone acetylation, the MLL1 complex and the NSL1 complex. As part of the NSL complex it is involved in acetylation of nucleosomal histone H4 on several lysine residues and therefore may be involved in the regulation of transcription.

22. Symbol: **PLEKHM1**; Name: **Pleckstrin Homology And RUN Domain Containing M1**

The protein encoded by this gene is essential for bone resorption, and may play a critical role in vesicular transport in the osteoclast. It acts as a multivalent adapter protein that regulates Rab7-dependent and HOPS complex-dependent fusion events in the endolysosomal system and couples autophagic and the endocytic trafficking pathways. PLEKGN1 is a dual effector of RAB7A and ARL8B that simultaneously binds these GTPases, bringing about clustering and fusion of late endosomes and lysosomes. It is required for late stages of endolysosomal maturation, facilitating both endocytosis-mediated degradation of growth factor receptors and autophagosome clearance. For example, in case of infection, it contributes to Salmonella typhimurium pathogenesis by supporting the integrity of the Salmonella-containing vacuole (SCV) probably in concert with the HOPS complex and Rab7. Mutations in this gene are associated with *autosomal recessive osteopetrosis type 6* (OPTB6).

23. Symbol: **FBXO32**; Name: **F-Box Protein 32**

This gene encodes a member of the F-box protein family which is characterized by an approximately 40 amino acid motif, the F-box. The F-box proteins constitute one of the four subunits of the ubiquitin protein ligase complex called SCFs (SKP1-cullin-F-box), which function in phosphorylation-dependent ubiquitination. The protein probably recognizes and binds to phosphorylated target proteins during skeletal muscle atrophy. It is highly expressed during *muscle atrophy*, whereas mice deficient in this gene were found to be resistant to atrophy. This protein is thus a potential drug target for the treatment of *muscle atrophy.*

24. Symbol: **PLCD3**; Name: **1-Phosphatidylinositol 4,5-Bisphosphate Phosphodiesterase Delta-3**

This gene encodes a member of the phospholipase C family, which catalyze the hydrolysis of phosphatidylinositol 4,5-bisphosphate to generate the second messengers diacylglycerol and inositol 1,4,5-trisphosphate (IP3). Diacylglycerol and IP3 mediate a variety of cellular responses to extracellular stimuli by inducing protein kinase C and increasing cytosolic Ca(2+) concentrations. Its activity is inhibited by spermine, sphingosine, and several phospholipids. It is essential for trophoblast and placental development and may participate in cytokinesis by hydrolyzing PIP2 at the cleavage furrow; It also regulates *neurite outgrowth* through the inhibition of RhoA/Rho kinase signaling.

**GROUP 2**

25. Symbol: **TLR10**; Name: **Toll Like Receptor 10**

The protein encoded by this gene is a member of the Toll-like receptor (TLR) family which plays a fundamental role in pathogen recognition and activation of innate immunity. TLRs are highly conserved from Drosophila to humans and share structural and functional similarities. They recognize pathogen-associated molecular patterns (PAMPs) that are expressed on infectious agents, and mediate the production of cytokines necessary for the development of *effective immunity*. It acts via MYD88 and TRAF6, leading to NF-kappa-B activation, cytokine secretion and the inflammatory response.

26. Symbol: **GPR65**; Name: **G Protein-Coupled Receptor 65**

The protein encoded by this gene enables G protein-coupled receptor activity and is involved in several processes, including actin cytoskeleton reorganization, activation of GTPase activity, and positive regulation of stress fiber assembly. It is a receptor for the glycosphingolipid psychosine (PSY) and several related glycosphingolipids. It also plays a role in *immune response* by maintaining lysosome function and supporting phagocytosis-mediated intracellular *bacteria clearance* and may have a role in activation-induced cell death or differentiation of T-cells.

27. Symbol: **TLR1**; Name: **Toll Like Receptor 1**

The protein encoded by this gene is a member of the Toll-like receptor (TLR) family which plays a fundamental role in pathogen recognition and activation of innate immunity. This gene is ubiquitously expressed, and at higher levels than other TLR genes. This protein participates in the innate immune response to microbial agents, specifically recognizes diacylated and triacylated lipopeptides. It cooperates with TLR2 to mediate the innate immune response to bacterial lipoproteins or lipopeptides by forming the activation cluster TLR2:TLR1:CD14 in response to triacylated lipopeptides. This cluster triggers signaling from the cell surface and subsequently is targeted to the Golgi in a lipid-raft dependent pathway, acting via MYD88 and TRAF6, leading to NF-kappa-B activation, cytokine secretion and the inflammatory response.

28. Symbol: **KLHL38**; Name: **Kelch Like Family Member 38**

KLHL38 is a Protein Coding gene. Diseases associated with KLHL38 include *Posterior Myocardial Infarction.*

29. Symbol: **SPATA32**; Name: **Spermatogenesis Associated 32**

The protein encoded by this gene is predicted to enable actin binding activity, to be involved in spermatogenesis and active in perinuclear region of cytoplasm.

30. Symbol: **C17orf104**

This is an RNA Gene and is affiliated with the lncRNA class; Its function is unclear.

31. Symbol: **SPPL2C**; Name: **Signal Peptide Peptidase Like 2C**

The protein encoded by this gene enables protein homodimerization activity. And, it is predicted to be involved in membrane protein proteolysis. SPPL2C is integral component of cytoplasmic side of endoplasmic reticulum membrane and integral component of lumenal side of endoplasmic reticulum membrane. Diseases associated with SPPL2C include *chromosome 17Q21.31 Duplication Syndrome* and *Caplan's Syndrome*.

32. Symbol: **LRRC37A**; Name: **Leucine Rich Repeat Containing 37A**

The protein encoded by this gene is predicted to be integral component of membrane. Diseases associated with LRRC37A include *Koolen-De Vries Syndrome* and *Supranuclear Palsy, Progressive, 1*.

33. Symbol: **ARL17A**; Name: **ADP Ribosylation Factor Like GTPase 17A**

The protein encoded by this gene is predicted to enable GTP binding activity, involved in intracellular protein transport and vesicle-mediated transport. It functions as an allosteric activator of the cholera toxin catalytic subunit, an ADP-ribosyltransferase. Diseases associated with ARL17A include *Bardet-Biedl Syndrome* and *Retinitis Pigmentosa*.

34. Symbol: **ARL17B**; Name: **ADP Ribosylation Factor Like GTPase 17B**

The protein encoded by this gene is predicted to enable GTP binding activity, involved in intracellular protein transport and vesicle-mediated transport. It functions as an allosteric activator of the cholera toxin catalytic subunit, an ADP-ribosyltransferase.

35. Symbol: **STH**; Name: **Microtubule-Associated Protein Tau (MAPT) Intronic Transcript**

The protein encoded by this gene is involved in positive regulation of mRNA splicing via spliceosome. It is located in nucleus and perinuclear region of cytoplasm. Diseases associated with STH include *Frontotemporal Dementia* and *Chromosome 17Q21.31 Duplication Syndrome*.

36. Symbol: **C10orf91** (**LINC02870**); Name: **Long Intergenic Non-Protein Coding RNA 2870**

No functions have been annotated to this non-coding RNA yet.

37. Symbol: **WNT9B**; Name: **Wnt Family Member 9B**

This gene is a member of the WNT gene family. The WNT gene family consists of structurally related genes that encode secreted signaling proteins. These proteins have been implicated in oncogenesis and in several developmental processes, including regulation of cell fate and patterning during embryogenesis. Study of its expression in the teratocarcinoma cell line NT2 suggests that it may be implicated in the early process of *neuronal* differentiation of NT2 cells induced by retinoic acid. This gene is clustered with WNT3, another family member, in the chromosome 17q21 region. It belongs to the canonical Wnt/beta-catenin signaling pathway. It activates a signaling cascade in the metanephric mesenchyme that induces tubulogenesis and plays a role in cranofacial development and is required for normal fusion of the palate during embryonic.

38. Symbol: **RUNX2**; Name: **RUNX Family Transcription Factor 2**

This gene is a member of the RUNX family of transcription factors and encodes a nuclear protein with an Runt DNA-binding domain. This protein is essential for osteoblastic differentiation and skeletal morphogenesis and acts as a scaffold for nucleic acids and regulatory factors involved in skeletal gene expression. The protein can bind DNA both as a monomer or, with more affinity, as a subunit of a heterodimeric complex. Two regions of potential trinucleotide repeat expansions are present in the N-terminal region of the encoded protein, and these and other mutations in this gene have been associated with the bone development disorder cleidocranial dysplasia (CCD). Transcript variants that encode different protein isoforms result from the use of alternate promoters as well as alternate splicing. It is essential for the maturation of osteoblasts and both intramembranous and endochondral ossification. In osteoblasts, it supports transcription activation: synergizes with SPEN/MINT to enhance FGFR2-mediated activation of the osteocalcin FGF-responsive element (OCFRE).

39. Symbol: **C17orf53** (HROB); Name: **Homologous Recombination Factor With OB-Fold**

The protein encoded by this gene is predicted to enable single-stranded DNA binding activity and involved in DNA synthesis, DNA repair and interstrand cross-link repair. It functions by recruiting the MCM8-MCM9 helicase complex to sites of DNA damage to promote DNA repair synthesis.

40. Symbol: **EFCAB13**; Name: **EF-Hand Calcium Binding Domain 13**

EFCAB13 is a Protein Coding gene, and its annotations include calcium ion binding.

41. Symbol: **GJC1**; Name: **Gap Junction Protein Gamma 1**

This gene is a member of the connexin gene family. The encoded protein is a component of gap junctions, which are composed of arrays of intercellular channels that provide a route for the diffusion of low molecular weight materials from cell to cell. One gap junction consists of a cluster of closely packed pairs of transmembrane channels, the connexons, through which materials of low MW diffuse from one cell to a neighboring cell. Gene Ontology annotations related to this gene include ion channel activity and gap junction channel activity. They allow passive diffusion of molecules up to 1 kDa, including nutrients, metabolites (glucose), ions (K+, Ca2+) and second messengers (IP3, cAMP).

42. Symbol: **DBF4B**; Name: **DBF4 Zinc Finger B**

This gene encodes a regulator of the cell division cycle 7 homolog (S. cerevisiae) protein, a serine-threonine kinase which links cell cycle regulation to genome duplication. It is a regulatory subunit for CDC7 which activates its kinase activity thereby playing a central role in DNA replication and cell proliferation and is required for progression of S and M phases. The complex CDC7-DBF4B selectively phosphorylates MCM2 subunit at 'Ser-40' and then is involved in regulating the initiation of DNA replication during cell cycle.

43. Symbol: **HEXIM2**; Name: **HEXIM P-TEFb Complex Subunit 2**

This gene encodes a member of the HEXIM family of proteins. This protein is a component of the 7SK small nuclear ribonucleoprotein. This protein has been found to negatively regulate the kinase activity of the cyclin-dependent kinase P-TEFb, which phosphorylates multiple target proteins to promote transcriptional elongation. This gene is located approximately 7 kb downstream from related family member HEXIM1 on chromosome 17. It is a transcriptional regulator which functions as a general RNA polymerase II transcription inhibitor and is a core component of the 7SK RNP complex: in cooperation with 7SK snRNA sequesters P-TEFb in a large inactive 7SK snRNP complex preventing RNA polymerase II phosphorylation and subsequent transcriptional.

44. Symbol: **LRRC37A2**; Name: **leucine rich repeat containing 37 member A2**

The protein encoded by this gene is predicted to be an integral component of membrane.

Diseases associated with LRRC37A2 include *Epilepsy*, *Progressive Myoclonic 6* and *Developmental And Epileptic Encephalopathy 96*.

45. Symbol: **DMGDH**; Name: **dimethylglycine dehydrogenase**

This gene encodes an enzyme involved in the catabolism of choline, catalyzing the

oxidative demethylation of dimethylglycine to form sarcosine. The enzyme is found

as a monomer in the mitochondrial matrix, and uses flavin adenine dinucleotide

and folate as cofactors. Mutation in this gene causes *dimethylglycine dehydrogenase*

*deficiency*, characterized by a fishlike body odor, chronic muscle fatigue, and elevated

levels of the muscle form of creatine kinase in serum.

**GROUP 3**

46. Symbol: **WNT3**; Name: **Wnt family member 3**

The WNT gene family consists of structurally related genes which encode secreted signaling proteins. These proteins have been implicated in oncogenesis and in several developmental processes, including regulation of cell fate and patterning during embryogenesis. Studies of the gene expression suggest that this gene may play a key role in some cases of human breast, rectal, lung, and gastric cancer through activation of the WNT-beta-catenin-TCF signaling pathway. The WNT3 protein functions in the canonical Wnt signaling pathway that results in activation of transcription factors of the TCF/LEF family. And it is required for normal gastrulation, formation of the primitive streak, and for the formation of the mesoderm during early embryogenesis.

47. Symbol: **C1QL1**; Name: **complement C1q like 1**

The protein encoded by this gene is predicted to enable signaling receptor binding activity, act upstream of or within maintenance of synapse structure, motor learning, and *neuron remodeling*. It is predicted to be located in several cellular components, including climbing fiber; presynapse; and synaptic cleft. It may also regulate the number of excitatory synapses that are formed on hippocampus neurons but has no effect on inhibitory synapses.

48. Symbol: **MAPT**; Name: **microtubule associated protein tau**

This gene encodes the microtubule-associated protein tau (MAPT) whose transcript undergoes complex, regulated alternative splicing, giving rise to several mRNA species. MAPT transcripts are differentially expressed in the nervous system, depending on stage of neuronal maturation and neuron type. This protein promotes microtubule assembly and stability, and might be involved in the establishment and maintenance of neuronal polarity. The C-terminus binds axonal microtubules while the N-terminus binds neural plasma membrane components, suggesting that tau functions as a linker protein between both. Axonal polarity is predetermined by TAU/MAPT localization (in the neuronal cell) in the domain of the cell body defined by the centrosome. The short isoforms allow plasticity of the cytoskeleton whereas the longer isoforms may preferentially play a role in its stabilization. MAPT gene mutations have been associated with several neurodegenerative disorders such as *Alzheimer's disease*, *Pick's disease*, *frontotemporal dementia*, *cortico-basal degeneration* and *progressive supranuclear palsy*.

49. Symbol: **CRHR1**; Name: **corticotropin releasing hormone receptor 1**

This gene encodes a G-protein coupled receptor that binds neuropeptides of the corticotropin releasing hormone family that are major regulators of the *hypothalamic-pituitary-adrenal pathway*. The encoded protein is essential for the activation of signal transduction pathways that regulate diverse physiological processes including stress, reproduction, immune response and obesity. This G-protein coupled receptor for CRH (corticotropin-releasing factor) and UCN (urocortin) has high affinity for CRH and UCN: Ligand binding causes a conformation change that triggers signaling via guanine nucleotide-binding proteins (G proteins) and down-stream effectors, such as adenylate cyclase. It promotes the activation of adenylate cyclase, leading to increased intracellular cAMP levels, inhibits the activity of the calcium channel CACNA1H. CRHR1 is required for normal embryonic development of the adrenal gland and for normal hormonal responses to stress. 50. Symbol: **ECE2**; Name: **Endothelin Converting Enzyme 2**

The enzym coded by this gene enables metalloendopeptidase activity. It is involved in peptide hormone processing, converts big endothelin-1 to endothelin-1. And it is also involved in the processing of various neuroendocrine peptides, including neurotensin, angiotensin I, substance P, proenkephalin-derived peptides, and prodynorphin-derived peptides.

51. Symbol: **CAMK2N2**; Name: **Calcium/Calmodulin Dependent Protein Kinase II Inhibitor 2**

This gene encodes a protein that is highly similar to the rat CaM-KII inhibitory protein, an inhibitor of calcium/calmodulin-dependent protein kinase II (CAMKII). CAMKII regulates numerous physiological functions, including neuronal synaptic plasticity through the phosphorylation of alpha-amino-3-hydroxy-5-methyl-4-isoxazolepropionic acid-type glutamate (AMPA) receptors. Studies of the similar protein in rat suggest that this protein may function as a negative regulator of CaM-KII and may act to inhibit the phosphorylation of AMPA receptors. Gene Ontology (GO) annotations related to this gene include protein kinase binding and calcium-dependent protein kinase inhibitor activity. Diseases associated with CAMK2N2 include *Amyotrophic Lateral Sclerosis 21* and *Amyotrophic Lateral Sclerosis Type 6*.

52. Symbol: **VWA5B2**; Name: **Von Willebrand Factor A Domain Containing 5B2**

Variations in this gene have been associated with Mathemtaics ability. Somatic mutations in it have been reported in various cancers.

53. Symbol: **GFAP**; Name: **Glial fibrillary acidic protein**

This gene encodes one of the major intermediate filament proteins of mature astrocytes. It is used as a marker to distinguish astrocytes from other glial cells during development. Mutations in this gene cause Alexander disease, a rare disorder of astrocytes in the central nervous system. GFAP, a class-III intermediate filament, is a cell-specific marker that distinguishes astrocytes from other glial cells during the development of the central nervous system. Defects in GFAP are a cause of *Alexander disease* (ALEXD), which is a rare disorder of the central nervous system.

54. Symbol: **NKX6-2**; Name: **NK6 homeobox 2**

The protein encoded by this gene enables sequence-specific double-stranded DNA binding activity and is Predicted to be involved in cell differentiation, regulation of myelination and transcription. It acts upstream of or within several processes, including negative regulation of transcription by RNA polymerase II, neurogenesis; and neuromuscular process controlling balance. Diseases associated with mutations in this gene include *Spastic Ataxia 8*.

### **Table S6. Harmonized instrumental SNPs (p<5x10^-8^) for the BAG to SCZ MR.**

| SNP | CHR | POS | A1 | A2 | A1_exp_frq | Beta_exp | SE_exp | N_exp | P_exp | A1_out_frq | Beta_out | SE_out | N_out | P_out |
| --- | --- | --- | --- | --- | --- | --- | --- | --- | --- | --- | --- | --- | --- | --- |
| rs10137195 | 14 | 88444882 | C | A | 0.36 | -0.16 | 0.0257 | 28104 | 1.42e-10 | 0.44 | 0.0056 | 0.0082 | 73173 | 0.49 |
| rs2239923 | 17 | 43176804 | T | C | 0.29 | -0.15 | 0.0269 | 28104 | 2.18e-8 | 0.28 | 0.0251 | 0.0086 | 73173 | 3.33e-3 |
| rs2242189 | 4 | 38691024 | C | T | 0.38 | 0.21 | 0.0257 | 28104 | 1.24e-16 | 0.33 | -0.0032 | 0.0084 | 73173 | 0.70 |
| rs2790102 | 6 | 45432214 | A | G | 0.35 | -0.15 | 0.0260 | 28104 | 8.92e-9 | 0.32 | -0.0063 | 0.0084 | 73173 | 0.45 |
| rs4880424 | 10 | 134584577 | T | C | 0.22 | 0.16 | 0.0296 | 28104 | 3.69e-8 | 0.20 | -0.0121 | 0.0098 | 73173 | 0.22 |
| rs62063276 | 17 | 44036408 | G | T | 0.21 | 0.28 | 0.0293 | 28104 | 2.72e-22 | 0.20 | -0.0588 | 0.0108 | 60448 | 5.92e-8 |
| rs7209501 | 17 | 43503294 | C | A | 0.18 | 0.29 | 0.0315 | 28104 | 5.33e-20 | 0.16 | -0.0482 | 0.0114 | 64002 | 2.50e-5 |
| rs7461069 | 8 | 124669029 | A | G | 0.21 | -0.17 | 0.0309 | 28104 | 1.57e-8 | 0.18 | -0.0149 | 0.01 | 73173 | 0.14 |

Instrumental SNPs select for the BAG to SCZ classical Mendelian randomization analysis. CHR: chromosome; POS: hg19 genomic position; A1: effective allele; A2: the other allele; A1_exp_frq: A1 frequency in the exposure dataset; Beta_exp, effect of A1 for the exposure; SE_exp; standard error for Beta_exp; N_exp: sample size for exposure; P_exp, association p value for exposure; A1_out_frq: A1 frequency in the outcome dataset; Beta_out: effect size of A1 on outcome; SE_out: standard error of Beta_out; N_out: sample size for outcome; P_out: association p value for outcome.

### **Table S7. Harmonized instrumental SNPs (p<5x10^-8^) for the BAG to BIP MR.**

| SNP | CHR | POS | A1 | A2 | A1_exp_frq | Beta_exp | SE_exp | N_exp | P_exp | A1_out_frq | Beta_out | SE_out | N_out | P_out |
| --- | --- | --- | --- | --- | --- | --- | --- | --- | --- | --- | --- | --- | --- | --- |
| rs10137195 | 14 | 88444882 | C | A | 0.36 | -0.16 | 0.0257 | 28104 | 1.42e-10 | 0.39 | -0.0151 | 0.0098 | 50981 | 0.13 |
| rs2239923 | 17 | 43176804 | T | C | 0.29 | -0.15 | 0.0269 | 28104 | 2.18e-8 | 0.28 | -0.0333 | 0.0104 | 50981 | 1.3e-3 |
| rs2242189 | 4 | 38691024 | C | T | 0.38 | 0.21 | 0.0257 | 28104 | 1.24e-16 | 0.34 | -0.0021 | 0.0098 | 50981 | 0.83 |
| rs2790102 | 6 | 45432214 | A | G | 0.35 | -0.15 | 0.0260 | 28104 | 8.92e-9 | 0.35 | 0.0017 | 0.0098 | 50981 | 0.87 |
| rs4880424 | 10 | 134584577 | T | C | 0.22 | 0.16 | 0.0296 | 28104 | 3.69e-8 | 0.21 | -0.0045 | 0.0115 | 50981 | 0.69 |
| rs62063276 | 17 | 44036408 | G | T | 0.21 | 0.28 | 0.0293 | 28104 | 2.72e-22 | 0.19 | 0.0215 | 0.0116 | 50981 | 0.06 |
| rs7209501 | 17 | 43503294 | C | A | 0.18 | 0.29 | 0.0315 | 28104 | 5.34e-20 | 0.16 | 0.0218 | 0.0125 | 50981 | 0.08 |
| rs7461069 | 8 | 124669029 | A | G | 0.21 | -0.17 | 0.0309 | 28104 | 1.57e-8 | 0.20 | 0.0134 | 0.0118 | 50981 | 0.26 |

Instrumental SNPs select for the BAG to BIP classical Mendelian randomization analysis. CHR: chromosome; POS: hg19 genomic position; A1: effective allele; A2: the other allele; A1_exp_frq: A1 frequency in the exposure dataset; Beta_exp, effect of A1 for the exposure; SE_exp; standard error for Beta_exp; N_exp: sample size for exposure; P_exp, association p value for exposure; A1_out_frq: A1 frequency in the outcome dataset; Beta_out: effect size of A1 on outcome; SE_out: standard error of Beta_out; N_out: sample size for outcome; P_out: association p value for outcome.

### **Table S8. Harmonized instrumental SNPs (p<5x10^-8^) for the BAG to MDD MR.**

| SNP | CHR | POS | A1 | A2 | A1_exp_frq | Beta_exp | SE_exp | N_exp | P_exp | A1_out_frq | Beta_out | SE_out | N_out | P_out |
| --- | --- | --- | --- | --- | --- | --- | --- | --- | --- | --- | --- | --- | --- | --- |
| rs10137195 | 14 | 88444882 | C | A | 0.36 | -0.16 | 0.0257 | 28104 | 1.421e-10 | 0.3596 | 0.0051 | 0.0082 | 69115 | 0.53 |
| rs11079732 | 17 | 44161581 | T | C | 0.45 | -0.14 | 0.0247 | 28104 | 4.67e-9 | 0.4537 | -0.0076 | 0.0092 | 52547 | 0.41 |
| rs1879581 | 17 | 43545893 | C | T | 0.18 | 0.29 | 0.0315 | 28104 | 3.23e-20 | 0.1766 | 0.0214 | 0.0105 | 69115 | 0.04 |
| rs2239923 | 17 | 43176804 | T | C | 0.29 | -0.15 | 0.0269 | 28104 | 2.18e-08 | 0.284 | 0.002 | 0.0088 | 69115 | 0.82 |
| rs2242189 | 4 | 38691024 | C | T | 0.38 | 0.21 | 0.0257 | 28104 | 1.24e-16 | 0.3681 | -0.0115 | 0.0082 | 69115 | 0.16 |
| rs2790102 | 6 | 45432214 | A | G | 0.35 | -0.15 | 0.0260 | 28104 | 8.92e-9 | 0.3487 | 0.0041 | 0.0084 | 69115 | 0.62 |
| rs4880424 | 10 | 134584577 | T | C | 0.22 | 0.16 | 0.0296 | 28104 | 3.69e-8 | 0.2144 | -0.0175 | 0.0103 | 69115 | 0.09 |
| rs7461069 | 8 | 124669029 | A | G | 0.21 | -0.17 | 0.0309 | 28104 | 1.57e-8 | 0.1953 | 0.0008 | 0.0099 | 69115 | 0.93 |

Instrumental SNPs select for the BAG to MDD classical Mendelian randomization analysis. CHR: chromosome; POS: hg19 genomic position; A1: effective allele; A2: the other allele; A1_exp_frq: A1 frequency in the exposure dataset; Beta_exp, effect of A1 for the exposure; SE_exp; standard error for Beta_exp; N_exp: sample size for exposure; P_exp, association p value for exposure; A1_out_frq: A1 frequency in the outcome dataset; Beta_out: effect size of A1 on outcome; SE_out: standard error of Beta_out; N_out: sample size for outcome; P_out: association p value for outcome.

### **Table S9. Harmonized instrumental SNPs (p<5x10^-8^) for the BAG to AD MR.**

| SNP | CHR | POS | A1 | A2 | A1_exp_frq | Beta_exp | SE_exp | N_exp | P_exp | A1_out_frq | Beta_out | SE_out | N_out | P_out |
| --- | --- | --- | --- | --- | --- | --- | --- | --- | --- | --- | --- | --- | --- | --- |
| rs10137195 | 14 | 88444882 | C | A | 0.36 | -0.1648 | 0.0257 | 28104 | 1.421e-10 | 0.37 | 0.0008 | 0.0022 | 429826.3 | 0.71 |
| rs2239923 | 17 | 43176804 | T | C | 0.29 | -0.1507 | 0.0269 | 28104 | 4.67e-9 | 0.28 | -0.0032 | 0.0024 | 428474.56 | 0.19 |
| rs2242189 | 4 | 38691024 | C | T | 0.38 | 0.2126 | 0.0257 | 28104 | 3.23e-20 | 0.34 | -0.0025 | 0.0023 | 417305.82 | 0.27 |
| rs2790102 | 6 | 45432214 | A | G | 0.35 | -0.1493 | 0.0260 | 28104 | 2.18e-08 | 0.35 | -0.0056 | 0.0023 | 428230.78 | 0.01 |
| rs4880424 | 10 | 134584577 | T | C | 0.22 | 0.1632 | 0.0296 | 28104 | 1.24e-16 | 0.22 | 0.0003 | 0.0026 | 426375.66 | 0.92 |
| rs62063276 | 17 | 44036408 | G | T | 0.21 | 0.2842 | 0.0293 | 28104 | 8.92e-9 | 0.21 | -0.0054 | 0.0027 | 427814.95 | 0.04 |
| rs7209501 | 17 | 43503294 | C | A | 0.18 | 0.2888 | 0.0315 | 28104 | 3.69e-8 | 0.17 | -0.0029 | 0.0028 | 429246.56 | 0.30 |
| rs7461069 | 8 | 124669029 | A | G | 0.21 | -0.1745 | 0.0309 | 28104 | 1.57e-8 | 0.18 | -0.0011 | 0.0028 | 429105.83 | 0.69 |

Instrumental SNPs select for the BAG to AD classical Mendelian randomization analysis. CHR: chromosome; POS: hg19 genomic position; A1: effective allele; A2: the other allele; A1_exp_frq: A1 frequency in the exposure dataset; Beta_exp, effect of A1 for the exposure; SE_exp; standard error for Beta_exp; N_exp: sample size for exposure; P_exp, association p value for exposure; A1_out_frq: A1 frequency in the outcome dataset; Beta_out: effect size of A1 on outcome; SE_out: standard error of Beta_out; N_out: sample size for outcome; P_out: association p value for outcome.

### **Table S10. Harmonized instrumental SNPs (p<5x10^-8^) for the BAG to PD MR.**

| SNP | CHR | POS | A1 | A2 | A1_exp_frq | Beta_exp | SE_exp | N_exp | P_exp | A1_out_frq | Beta_out | SE_out | N_out | P_out |
| --- | --- | --- | --- | --- | --- | --- | --- | --- | --- | --- | --- | --- | --- | --- |
| rs10137195 | 14 | 88444882 | C | A | 0.36 | -0.1648 | 0.0257 | 28104 | 1.421e-10 | 0.37 | 0.0602 | 0.0174 | 62649 | 5e-4 |
| rs2239923 | 17 | 43176804 | T | C | 0.29 | -0.1507 | 0.0269 | 28104 | 4.67e-9 | 0.31 | 0.0446 | 0.0214 | 62649 | 0.04 |
| rs2242189 | 4 | 38691024 | C | T | 0.38 | 0.2126 | 0.0257 | 28104 | 3.23e-20 | 0.36 | 0.0151 | 0.0233 | 49863 | 0.52 |
| rs2790102 | 6 | 45432214 | A | G | 0.35 | -0.1493 | 0.0260 | 28104 | 2.18e-08 | 0.34 | -0.0111 | 0.0238 | 49863 | 0.64 |
| rs4880424 | 10 | 134584577 | T | C | 0.22 | 0.1632 | 0.0296 | 28104 | 1.24e-16 | 0.22 | 0.0078 | 0.0274 | 49863 | 0.77 |
| rs62063276 | 17 | 44036408 | G | T | 0.21 | 0.2842 | 0.0293 | 28104 | 8.92e-9 | 0.22 | -0.2507 | 0.027 | 60613 | 1.46e-20 |
| rs7209501 | 17 | 43503294 | C | A | 0.18 | 0.2888 | 0.0315 | 28104 | 3.69e-8 | 0.19 | -0.2225 | 0.0274 | 60613 | 4.66e-16 |
| rs7461069 | 8 | 124669029 | A | G | 0.21 | -0.1745 | 0.0309 | 28104 | 1.57e-8 | 0.19 | 0.0252 | 0.0256 | 62649 | 0.33 |

Instrumental SNPs select for the BAG to PD classical Mendelian randomization analysis. CHR: chromosome; POS: hg19 genomic position; A1: effective allele; A2: the other allele; A1_exp_frq: A1 frequency in the exposure dataset; Beta_exp, effect of A1 for the exposure; SE_exp; standard error for Beta_exp; N_exp: sample size for exposure; P_exp, association p value for exposure; A1_out_frq: A1 frequency in the outcome dataset; Beta_out: effect size of A1 on outcome; SE_out: standard error of Beta_out; N_out: sample size for outcome; P_out: association p value for outcome.

### **Table S11. Harmonized instrumental SNPs (p<5x10^-8^) for the SCZ to BAG MR.**

| SNP | CHR | POS | A1 | A2 | A1_exp_frq | Beta_exp | SE_exp | N_exp | P_exp | A1_out_frq | Beta_out | SE_out | N_out | P_out |
| --- | --- | --- | --- | --- | --- | --- | --- | --- | --- | --- | --- | --- | --- | --- |
| rs10098869 | 8 | 4208761 | T | C | 0.85 | -0.07 | 0.0108 | 73173 | 2.85e-10 | 0.86 | -0.0265 | 0.0358 | 28104 | 0.4 |
| 7rs10127983 | 1 | 153923276 | C | T | 0.70 | -0.05 | 0.0084 | 73173 | 3.11e-8 | 0.70 | -0.0233 | 0.0271 | 28104 | 0.39 |
| rs10167385 | 2 | 37238824 | C | T | 0.68 | -0.05 | 0.0086 | 73173 | 7.94e-9 | 0.75 | 0.0046 | 0.0287 | 28104 | 0.87 |
| rs10238960 | 7 | 70773271 | C | T | 0.32 | -0.05 | 0.0084 | 73173 | 7.65e-9 | 0.30 | -0.021 | 0.0270 | 28104 | 0.44 |
| rs10243922 | 7 | 138599980 | G | A | 0.73 | -0.05 | 0.0089 | 73173 | 1.02e-8 | 0.69 | 0.0496 | 0.0267 | 28104 | 0.06 |
| rs1034326 | 6 | 30084446 | A | G | 0.88 | 0.12 | 0.0126 | 66018 | 1.92e-22 | 0.84 | 0.0664 | 0.0326 | 28104 | 0.04 |
| rs10447226 | 5 | 139028810 | G | T | 0.75 | -0.05 | 0.0092 | 73173 | 1.99e-8 | 0.72 | -0.0208 | 0.0272 | 28104 | 0.45 |
| rs10520313 | 4 | 176744539 | G | A | 0.92 | -0.09 | 0.014 | 73173 | 8.73e-11 | 0.93 | -0.0424 | 0.0477 | 28104 | 0.37 |
| rs10736470 | 11 | 113418371 | G | A | 0.29 | -0.07 | 0.009 | 73173 | 1.11e-14 | 0.36 | 0.0169 | 0.0256 | 28104 | 0.51 |
| rs10774034 | 12 | 2330458 | C | T | 0.66 | -0.08 | 0.0085 | 73173 | 7.10e-23 | 0.61 | -0.054 | 0.0251 | 28104 | 0.03 |
| rs10774545 | 12 | 110564659 | A | C | 0.28 | 0.06 | 0.0086 | 73173 | 6.98e-13 | 0.28 | -0.0109 | 0.0274 | 28104 | 0.69 |
| rs10861176 | 12 | 104631552 | G | A | 0.27 | -0.05 | 0.0086 | 73173 | 5.23e-9 | 0.27 | 5,003-4 | 0.0280 | 28104 | 0.99 |
| rs10861879 | 12 | 108609634 | G | A | 0.64 | -0.05 | 0.0083 | 73173 | 1.23e-8 | 0.69 | 0.0397 | 0.0264 | 28104 | 0.13 |
| rs10873538 | 14 | 104255569 | T | G | 0.66 | -0.06 | 0.0082 | 73173 | 9.59e-15 | 0.66 | 0.0409 | 0.0258 | 28104 | 0.11 |
| rs10894308 | 11 | 130891895 | G | A | 0.54 | 0.05 | 0.0079 | 73173 | 1.36e-9 | 0.55 | -0.0263 | 0.0247 | 28104 | 0.29 |
| rs10906984 | 15 | 83366296 | A | C | 0.39 | 0.05 | 0.0079 | 73173 | 4.84e-10 | 0.38 | -0.0228 | 0.0255 | 28104 | 0.37 |
| rs10985811 | 9 | 101070487 | T | C | 0.80 | -0.05 | 0.0098 | 73173 | 2.53e-8 | 0.83 | 0.0114 | 0.0333 | 28104 | 0.73 |
| rs11038906 | 11 | 46536442 | G | A | 0.85 | -0.09 | 0.0108 | 73173 | 6.43e-16 | 0.83 | -0.0047 | 0.0327 | 28104 | 0.89 |
| rs11122119 | 1 | 6768856 | C | A | 0.65 | -0.05 | 0.0081 | 73173 | 2.31e-8 | 0.68 | -0.0419 | 0.0262 | 28104 | 0.11 |
| rs11159633 | 14 | 84670453 | T | G | 0.37 | -0.05 | 0.008 | 73173 | 1.89e-8 | 0.38 | 0.0589 | 0.0251 | 28104 | 0.02 |
| rs11167584 | 5 | 152004727 | A | C | 0.78 | 0.05 | 0.0095 | 73173 | 5.47e-9 | 0.73 | 0.0045 | 0.0269 | 28104 | 0.87 |
| rs11210203 | 1 | 73775513 | A | G | 0.36 | 0.06 | 0.0082 | 73173 | 2.31e-15 | 0.37 | 0.0146 | 0.0255 | 28104 | 0.57 |
| rs1121296 | 1 | 72174197 | T | C | 0.59 | 0.05 | 0.008 | 73173 | 3.74e-9 | 0.63 | -0.0245 | 0.0255 | 28104 | 0.34 |
| rs112205523 | 19 | 50114350 | C | T | 0.71 | 0.06 | 0.0088 | 73173 | 1.82e-10 | 0.69 | 0.0613 | 0.0266 | 28104 | 0.02 |
| rs1143702 | 1 | 44086831 | C | T | 0.33 | 0.07 | 0.0082 | 73173 | 2.76e-16 | 0.3296 | -0.0012 | 0.0259 | 28104 | 0.9631 |
| rs1157635 | 11 | 24404596 | A | G | 0.52 | -0.04 | 0.0078 | 73173 | 2.00e-8 | 0.4917 | 0.05 | 0.0246 | 28104 | 0.04242 |
| rs11696755 | 20 | 48105317 | T | C | 0.83 | -0.07 | 0.0104 | 73173 | 2.99e-10 | 0.824 | -0.0148 | 0.0318 | 28104 | 0.6422 |
| rs11715134 | 3 | 50552866 | G | A | 0.89 | -0.07 | 0.0123 | 73173 | 2.52e-28 | 0.9105 | -0.0361 | 0.0418 | 28104 | 0.3878 |
| rs11807834 | 1 | 230272624 | G | A | 0.76 | -0.05 | 0.0093 | 73173 | 1.12e-8 | 0.7399 | -0.0294 | 0.0281 | 28104 | 0.2962 |
| rs11890137 | 2 | 198816662 | G | A | 0.36 | 0.06 | 0.0081 | 73173 | 7.34e-12 | 0.3253 | -0.0883 | 0.0262 | 28104 | 7.61e-4 |
| rs11899740 | 2 | 73631212 | G | A | 0.79 | -0.06 | 0.0103 | 63155 | 9.94e-9 | 0.7783 | -0.0209 | 0.0295 | 28104 | 0.479 |
| rs1191551 | 14 | 30000405 | T | G | 0.18 | 0.06 | 0.0103 | 73173 | 4.72e-9 | 0.2229 | -0.0089 | 0.0291 | 28104 | 0.7598 |
| rs11956576 | 5 | 91001001 | A | C | 0.78 | -0.05 | 0.0093 | 73173 | 1.00e-8 | 0.7968 | -0.0334 | 0.0310 | 28104 | 0.2821 |
| rs12126806 | 1 | 200963825 | C | T | 0.75 | 0.05 | 0.009 | 73173 | 2.89e-8 | 0.7062 | 0.0392 | 0.0267 | 28104 | 0.1434 |
| rs12131135 | 1 | 200418613 | G | T | 0.55 | 0.04 | 0.0081 | 73173 | 3.68e-8 | 0.4924 | -0.0274 | 0.0244 | 28104 | 0.2632 |
| rs12244388 | 10 | 104640052 | G | A | 0.61 | 0.07 | 0.008 | 73173 | 1.59e-18 | 0.6684 | 4,00E-04 | 0.0279 | 28104 | 0.9886 |
| rs12277680 | 11 | 134586708 | A | G | 0.51 | -0.05 | 0.0081 | 73173.9 | 3.84e-11 | 0.5707 | 0.0132 | 0.0248 | 28104 | 0.5947 |
| rs12301769 | 12 | 72231313 | A | C | 0.91 | -0.08 | 0.014 | 72630.42 | 1.92e-9 | 0.90081 | -0.0052 | 0.0418 | 28104 | 0.9011 |
| rs12311848 | 12 | 124486851 | A | G | 0.70 | -0.05 | 0.0087 | 73173.9 | 1.65e-8 | 0.6713 | 0.0486 | 0.0259 | 28104 | 0.06069 |
| rs12614183 | 2 | 105005181 | C | T | 0.78 | -0.05 | 0.0095 | 73173.9 | 4.17e-8 | 0.8232 | -0.0128 | 0.0320 | 28104 | 0.6898 |
| rs12652777 | 5 | 155775075 | T | C | 0.43 | 0. | 0.0079 | 73173.9 | 1.06e-8 | 0.4916 | 0.0137 | 0.0246 | 28104 | 0.5781 |
| rs12668848 | 7 | 2020995 | G | A | 0.56 | 0.08 | 0.0078 | 73173.9 | 3.21e-26 | 0.5709 | 0.0043 | 0.0247 | 28104 | 0.8618 |
| rs12713008 | 2 | 48503561 | G | A | 0.55 | 0.04 | 0.0078 | 73173.9 | 3.05e-8 | 0.5428 | 0.001 | 0.0239 | 28104 | 0.9667 |
| rs12883788 | 14 | 33303540 | C | T | 0.57 | -0.05 | 0.0079 | 73173.9 | 8.43e-12 | 0.547 | -0.0085 | 0.0244 | 28104 | 0.7281 |
| rs12905223 | 15 | 85114268 | T | C | 0.75 | 0.067 | 0.009 | 73173.9 | 1.65e-10 | 0.7288 | -0.0252 | 0.0273 | 28104 | 0.3569 |
| rs12950148 | 17 | 12872802 | A | G | 0.83 | -0.06 | 0.0105 | 73173.9 | 1.48e-09 | 0.8189 | -0.0467 | 0.0316 | 28104 | 0.1406 |
| rs12969453 | 18 | 52751708 | A | G | 0.56 | 0.05 | 0.0078 | 73173.9 | 3.53e-12 | 0.5989 | 0.0059 | 0.02509 | 28104 | 0.8141 |
| rs13090130 | 3 | 161777035 | G | A | 0.62 | 0.05 | 0.0079 | 73173.9 | 9.92e-11 | 0.6343 | -0.0143 | 0.02538 | 28104 | 0.5732 |
| rs13107325 | 4 | 103188709 | C | T | 0.93 | -0.16 | 0.0168 | 58439.19 | 2.9e-21 | 0.9263 | 0.0877 | 0.0482 | 28104 | 0.06889 |
| rs13115045 | 4 | 143866615 | G | A | 0.78 | 0.05 | 0.0094 | 73173.9 | 4.36e-8 | 0.7626 | 0.0301 | 0.0288 | 28104 | 0.2976 |
| rs13211901 | 6 | 27187612 | G | A | 0.84 | 0.11 | 0.011 | 71487.15 | 9.02e-26 | 0.798 | -0.018 | 0.0299 | 28104 | 0.5476 |
| rs13219424 | 6 | 128332084 | C | T | 0.67 | 0.05 | 0.0084 | 73173 | 4.52e-8 | 0.6986 | -0.0287 | 0.0268 | 28104 | 0.285 |
| rs13230189 | 7 | 137073259 | C | T | 0.668 | 0.07 | 0.0081 | 73173 | 1.04e-18 | 0.6427 | 0.0358 | 0.0258 | 28104 | 0.166 |
| rs133293 | 22 | 42382797 | G | A | 0.45 | -0.05 | 0.0081 | 73173 | 1.85e-11 | 0.5284 | -0.0394 | 0.0246 | 28104 | 0.1106 |
| rs1401988 | 12 | 103561799 | C | T | 0.67 | 0.058 | 0.0085 | 73173 | 4.63e-8 | 0.631 | 0.0145 | 0.0251 | 28104 | 0.564 |
| rs1480380 | 6 | 32913246 | C | T | 0.92 | 0.15 | 0.0159 | 59322 | 4.75e-22 | 0.899 | 0.1107 | 0.0394 | 28104 | 5.07e-3 |
| rs1553933 | 2 | 60509080 | C | T | 0.38 | 0.04 | 0.008 | 73173 | 3.89e-8 | 0.4104 | -0.0162 | 0.0249 | 28104 | 0.5157 |
| rs1600295 | 3 | 181234906 | C | T | 0.17 | -0.07 | 0.0109 | 73173 | 1.29e-11 | 0.2058 | -0.0061 | 0.0304 | 28104 | 0.8413 |
| rs16825349 | 2 | 146425531 | A | G | 0.83 | -0.07 | 0.0104 | 73173 | 1.32e-11 | 0.8161 | 0.0219 | 0.0314 | 28104 | 0.4856 |
| rs169738 | 6 | 33537546 | A | G | 0.42 | -0.05 | 0.0079 | 73173 | 4.39e-11 | 0.4003 | -0.0234 | 0.0250 | 28104 | 0.3508 |
| rs17194490 | 3 | 2547786 | G | T | 0.84 | -0.08 | 0.0116 | 60303 | 1.85e-11 | 0.8454 | -0.0316 | 0.0330 | 28104 | 0.3394 |
| rs17693963 | 6 | 27710165 | A | C | 0.92 | 0.19 | 0.0151 | 64563 | 1.32e-37 | 0.8858 | 0.0283 | 0.0368 | 28104 | 0.4431 |
| rs1784223 | 11 | 65380248 | T | C | 0.70 | -0.05 | 0.0086 | 73173 | 2.43e-8 | 0.6536 | 0.0329 | 0.0257 | 28104 | 0.2017 |
| rs1924378 | 13 | 38361744 | T | G | 0.48 | 0.05 | 0.0081 | 73173 | 9.31e-9 | 0.4139 | -0.0011 | 0.0249 | 28104 | 0.9648 |
| rs1939514 | 11 | 132572894 | T | C | 0.52 | 0.06 | 0.0077 | 73173 | 1.06e-12 | 0.4942 | -0.0443 | 0.0246 | 28104 | 0.0721 |
| rs209180 | 6 | 28793605 | C | T | 0.61 | 0.05 | 0.0083 | 73173 | 6.93e-10 | 0.6087 | 0.0618 | 0.0251 | 28104 | 0.01366 |
| rs2095401 | 6 | 43124720 | C | T | 0.83 | 0.06 | 0.0106 | 73173 | 4.61e-8 | 0.8643 | -0.0097 | 0.0361 | 28104 | 0.7884 |
| rs2119242 | 10 | 21344773 | G | A | 0.83 | -0.06 | 0.0106 | 73173 | 1.34e-8 | 0.8686 | -0.012 | 0.0354 | 28104 | 0.7351 |
| rs215483 | 4 | 23377121 | G | A | 0.69 | -0.05 | 0.0084 | 73173 | 1.59e-9 | 0.6638 | 0.02 | 0.02620 | 28104 | 0.4453 |
| rs2161203 | 5 | 88696876 | T | C | 0.89 | 0.08 | 0.0135 | 62066 | 2.49e-9 | 0.8772 | 0.0063 | 0.03664 | 28104 | 0.8635 |
| rs216221 | 17 | 2155430 | G | A | 0.32 | -0.05 | 0.0082 | 73173 | 8.65e-10 | 0.3494 | -0.0409 | 0.0256 | 28104 | 0.1115 |
| rs217291 | 6 | 84393942 | T | C | 0.57 | 0.05 | 0.0078 | 73173 | 5.47e-10 | 0.5632 | -0.0072 | 0.02479 | 28104 | 0.7715 |
| rs2190864 | 14 | 72416219 | T | C | 0.39 | 0.07 | 0.008 | 73173 | 1.12e-16 | 0.4004 | -0.0316 | 0.0248 | 28104 | 0.2044 |
| rs2224086 | 1 | 115309590 | C | A | 0.20 | -0.06 | 0.0103 | 73173 | 2.09e-8 | 0.2276 | -0.0215 | 0.02936 | 28104 | 0.464 |
| rs2252074 | 7 | 104594253 | T | G | 0.59 | -0.07 | 0.0078 | 73173 | 1.27e-14 | 0.6017 | 0.0252 | 0.0250 | 28104 | 0.314 |
| rs2319280 | 1 | 150014017 | A | C | 0.15 | -0.07 | 0.0118 | 61788 | 1.37e-8 | 0.1617 | -0.0177 | 0.03266 | 28104 | 0.5879 |
| rs2381411 | 9 | 36319928 | T | C | 0.57 | -0.05 | 0.0079 | 73173.9 | 1.28e-8 | 0.6055 | 0.0055 | 0.0253 | 28104 | 0.8283 |
| rs2455415 | 13 | 38860697 | C | T | 0.6 | -0.05 | 0.0081 | 73173.9 | 3.431e-09 | 0.5697 | 0.0366 | 0.02490 | 28104 | 0.1417 |
| rs246024 | 5 | 140333952 | C | T | 0.47 | 0.05 | 0.0077 | 73173.9 | 1.71e-09 | 0.4789 | 0.0513 | 0.02454 | 28104 | 0.03659 |
| rs2470578 | 3 | 17330092 | G | A | 0.60 | 0.05 | 0.0085 | 73173.9 | 3.49e-08 | 0.5221 | 0.0114 | 0.0246 | 28104 | 0.644 |
| rs2686386 | 12 | 121639657 | C | T | 0.21 | 0.05 | 0.0096 | 73173.9 | 1.26e-08 | 0.1837 | 0.0028 | 0.0320 | 28104 | 0.9303 |
| rs2710323 | 3 | 52815905 | T | C | 0.53 | 0.07 | 0.0077 | 73173.9 | 5.92e-22 | 0.533 | -0.0034 | 0.02483 | 28104 | 0.8911 |
| rs2802535 | 1 | 98508258 | C | T | 0.17 | -0.10 | 0.0105 | 73173.9 | 3.54e-20 | 0.1852 | 0.0628 | 0.0313 | 28104 | 0.04486 |
| rs2815731 | 6 | 73155285 | C | A | 0.64 | 0.05 | 0.0081 | 73173.9 | 3.41e-10 | 0.6654 | -0.0408 | 0.0259 | 28104 | 0.1153 |
| rs281774 | 2 | 200814572 | C | T | 0.20 | 0.08 | 0.0096 | 73173.9 | 3.68e-17 | 0.1881 | 0.0772 | 0.0308 | 28104 | 0.01223 |
| rs28669908 | 15 | 78910267 | C | A | 0.80 | 0.08 | 0.0098 | 73173.9 | 9.25e-15 | 0.7851 | 0.0296 | 0.02996 | 28104 | 0.3233 |
| rs28758902 | 18 | 53408187 | C | T | 0.54 | -0.05 | 0.0077 | 73173.9 | 7.69e-12 | 0.569 | 0.0239 | 0.02499 | 28104 | 0.339 |
| rs28768122 | 12 | 123885974 | T | C | 0.26 | 0.08 | 0.009 | 73173.9 | 3.71e-18 | 0.2508 | 0.0511 | 0.02892 | 28104 | 0.07733 |
| rs28865701 | 18 | 77620911 | A | G | 0.62 | -0.06 | 0.0079 | 73173.9 | 4.7e-16 | 0.6344 | -0.0274 | 0.02548 | 28104 | 0.2824 |
| rs2909457 | 2 | 162845855 | G | A | 0.38 | 0.05 | 0.0083 | 73173.9 | 3.09e-8 | 0.4546 | 0.0599 | 0.02459 | 28104 | 0.01488 |
| rs2910032 | 5 | 152540354 | C | T | 0.42 | 0.06 | 0.008 | 73173.9 | 4.52e-12 | 0.4826 | 0.0368 | 0.02456 | 28104 | 0.1341 |
| rs2914983 | 2 | 198314568 | A | G | 0.35 | 0.06 | 0.0081 | 73173.9 | 1.1e-14 | 0.3244 | -0.0706 | 0.0260 | 28104 | 6.78e-3 |
| rs2968533 | 7 | 71787660 | G | A | 0.60 | 0.05 | 0.008 | 73173.9 | 4.593e-9 | 0.5627 | -0.0411 | 0.02453 | 28104 | 0.09387 |
| rs308697 | 3 | 161487491 | C | A | 0.60 | 0.056 | 0.0079 | 73173.9 | 3.35e-9 | 0.5715 | -0.0306 | 0.02453 | 28104 | 0.2123 |
| rs312477 | 3 | 53515136 | G | A | 0.746 | 0.06 | 0.0089 | 73173.9 | 1.37e-10 | 0.7758 | -0.0624 | 0.02916 | 28104 | 0.03237 |
| rs3129963 | 6 | 32380208 | A | G | 0.84 | 0.09 | 0.0113 | 70162.94 | 8.34e-15 | 0.8193 | 0.1026 | 0.0308 | 28104 | 8.97e-4 |
| rs3131063 | 6 | 30763756 | G | A | 0.55 | 0.05 | 0.0082 | 69694.12 | 6.50e-9 | 0.5554 | 0.0617 | 0.0246 | 28104 | 0.01238 |
| rs3134797 | 6 | 32186050 | G | A | 0.59 | 0.07 | 0.0083 | 70162.94 | 2.42e-11 | 0.5667 | 0.0499 | 0.02464 | 28104 | 0.0429 |
| rs322128 | 19 | 11402416 | C | T | 0.78 | -0.06 | 0.0095 | 73173.9 | 2.08e-09 | 0.8021 | 0.008 | 0.0309 | 28104 | 0.7962 |
| rs337718 | 18 | 69774278 | T | C | 0.30 | 0.05 | 0.0084 | 73173.9 | 4.39e-09 | 0.2937 | -0.035 | 0.02695 | 28104 | 0.1942 |
| rs34829140 | 2 | 155904379 | C | A | 0.80 | -0.05 | 0.0098 | 73173.9 | 4.04e-08 | 0.7834 | 0.0216 | 0.02947 | 28104 | 0.4637 |
| rs35045093 | 7 | 127738471 | A | C | 0.826 | 0.06 | 0.0103 | 73173.9 | 3.36e-08 | 0.8248 | -0.0379 | 0.03322 | 28104 | 0.254 |
| rs35339313 | 2 | 145139727 | C | T | 0.61 | -0.06 | 0.008 | 73173.9 | 1.31e-12 | 0.6544 | 0.0248 | 0.02558 | 28104 | 0.3324 |
| rs353547 | 3 | 52268866 | T | C | 0.39 | -0.06 | 0.0079 | 73173.9 | 5.17e-14 | 0.3695 | -0.0028 | 0.02553 | 28104 | 0.9127 |
| rs35531336 | 7 | 133438132 | A | G | 0.89 | 0.07 | 0.013 | 73173.9 | 1.46e-08 | 0.8765 | -0.0251 | 0.0370 | 28104 | 0.4978 |
| rs35734242 | 4 | 706700 | T | C | 0.57 | -0.045 | 0.008 | 73173.9 | 3.927e-10 | 0.5683 | 9,00E-04 | 0.0251 | 28104 | 0.9714 |
| rs35844949 | 2 | 58380167 | G | A | 0.44 | -0.07 | 0.0078 | 73173.9 | 6.203e-21 | 0.4404 | -0.0058 | 0.0249 | 28104 | 0.8162 |
| rs36013966 | 8 | 89302448 | G | A | 0.78 | -0.06 | 0.0093 | 73173.9 | 7.548e-10 | 0.7662 | -0.0403 | 0.029 | 28104 | 0.1678 |
| rs36109883 | 6 | 26133070 | G | A | 0.93 | 0.18 | 0.0174 | 58248.36 | 6.789e-24 | 0.91004 | 0.0362 | 0.0407 | 28104 | 0.3741 |
| rs3732386 | 3 | 36871993 | C | T | 0.70 | -0.06 | 0.0087 | 73173.9 | 1.464e-10 | 0.6418 | 0.0433 | 0.025 | 28104 | 0.09107 |
| rs3739554 | 9 | 129940645 | A | G | 0.83 | -0.06 | 0.0103 | 73173.9 | 2.257e-08 | 0.8385 | -0.0034 | 0.033 | 28104 | 0.9194 |
| rs3765971 | 1 | 8445360 | C | T | 0.28 | 0.05 | 0.0088 | 73173.9 | 1.975e-09 | 0.3382 | -0.0131 | 0.025 | 28104 | 0.6132 |
| rs3779870 | 8 | 65519166 | A | G | 0.37 | -0.05 | 0.008 | 73173.9 | 7.63e-09 | 0.3731 | 0.0588 | 0.0255 | 28104 | 0.02144 |
| rs3808581 | 8 | 26250047 | G | A | 0.79 | -0.07 | 0.0097 | 73173.9 | 3.823e-12 | 0.8335 | 0.0564 | 0.033 | 28104 | 0.08788 |
| rs3929747 | 10 | 104214267 | C | T | 0.75 | 0.06 | 0.0091 | 73173.9 | 3.127e-12 | 0.7905 | 0.0041 | 0.03071 | 28104 | 0.8938 |
| rs39967 | 5 | 57744788 | T | C | 0.16 | -0.06 | 0.0107 | 73173.9 | 1.869e-08 | 0.1771 | -0.0376 | 0.03223 | 28104 | 0.2434 |
| rs4144848 | 2 | 200001233 | G | A | 0.54 | 0.07 | 0.0079 | 73173.9 | 2.328e-16 | 0.5789 | 0.0307 | 0.0248 | 28104 | 0.2163 |
| rs41533650 | 4 | 176869252 | G | A | 0.80 | -0.07 | 0.0097 | 73173.9 | 8.691e-14 | 0.7906 | -0.0196 | 0.0300 | 28104 | 0.5139 |
| rs4283241 | 16 | 29988349 | G | A | 0.57 | 0.06 | 0.0078 | 73173.9 | 2.531e-14 | 0.541 | -0.0886 | 0.0244 | 28104 | 2.91e-4 |
| rs431982 | 5 | 106732144 | T | C | 0.24 | 0.05 | 0.0093 | 73173.9 | 1.558e-08 | 0.1817 | 0.0593 | 0.0322 | 28104 | 0.06609 |
| rs4327046 | 16 | 9942377 | G | A | 0.73 | -0.06 | 0.0088 | 73173.9 | 4.1e-10 | 0.7061 | -0.0198 | 0.0274 | 28104 | 0.47 |
| rs4510068 | 17 | 44184828 | G | T | 0.60 | 0.05 | 0.0082 | 73173.9 | 1.757e-09 | 0.608 | -0.1634 | 0.0254 | 28104 | 1.28e-10 |
| rs459391 | 21 | 22120508 | T | C | 0.19 | 0.06 | 0.01 | 73173.9 | 1.543e-08 | 0.1932 | 0.0408 | 0.0308 | 28104 | 0.1863 |
| rs4712938 | 6 | 25449269 | T | C | 0.21 | -0.06 | 0.0097 | 73173.9 | 4.446e-09 | 0.237 | 0.0031 | 0.02914 | 28104 | 0.9153 |
| rs4793888 | 17 | 55737740 | G | A | 0.79 | -0.06 | 0.0097 | 73173.9 | 3.634e-10 | 0.8478 | -0.0307 | 0.03431 | 28104 | 0.371 |
| rs4808955 | 19 | 19552413 | T | C | 0.66 | -0.06 | 0.0081 | 73173.9 | 1.967e-13 | 0.646 | 0.0502 | 0.02578 | 28104 | 0.05159 |
| rs4936215 | 11 | 133852684 | A | G | 0.81 | 0.08 | 0.0104 | 72178.92 | 1.857e-15 | 0.7972 | 0.006 | 0.0303 | 28104 | 0.8431 |
| rs496108 | 5 | 49532628 | C | T | 0.45 | 0.05 | 0.0082 | 73173.9 | 8.617e-10 | 0.4659 | 0.0152 | 0.0247 | 28104 | 0.539 |
| rs4975957 | 5 | 46265302 | A | G | 0.46 | 0.05 | 0.0084 | 72359.83 | 3.581e-09 | 0.4686 | 0.0162 | 0.02479 | 28104 | 0.5135 |
| rs4990036 | 6 | 31323506 | C | T | 0.89 | 0.15 | 0.0135 | 64853.91 | 1.034e-27 | 0.8577 | 0.075 | 0.03397 | 28104 | 0.02726 |
| rs500102 | 9 | 77358745 | T | C | 0.43 | 0.04 | 0.0079 | 73173.9 | 4.153e-08 | 0.3944 | 0.0151 | 0.0250 | 28104 | 0.5467 |
| rs505061 | 9 | 22767164 | C | A | 0.51 | -0.05 | 0.0077 | 73173.9 | 1.025e-10 | 0.4962 | -0.0162 | 0.02430 | 28104 | 0.5051 |
| rs55704727 | 3 | 117616431 | C | T | 0.90 | 0.07 | 0.013 | 72630.42 | 3.296e-08 | 0.8757 | 0.0246 | 0.0365 | 28104 | 0.5014 |
| rs56086712 | 10 | 3800240 | T | C | 0.50 | 0.04 | 0.0081 | 73173.9 | 4.389e-08 | 0.4377 | 0.0199 | 0.0251 | 28104 | 0.4294 |
| rs5757730 | 22 | 39967430 | A | G | 0.39 | -0.05 | 0.0085 | 73173.9 | 1.185e-10 | 0.4986 | 0.0427 | 0.0245 | 28104 | 0.08162 |
| rs578470 | 12 | 50463325 | T | C | 0.62 | -0.05 | 0.0083 | 73173.9 | 2.327e-08 | 0.5828 | -0.0189 | 0.0248 | 28104 | 0.4467 |
| rs58946679 | 1 | 2372321 | C | T | 0.61 | -0.05 | 0.0084 | 73173.9 | 1.721e-10 | 0.6475 | 0.0067 | 0.0262 | 28104 | 0.7985 |
| rs6010045 | 22 | 51103091 | T | C | 0.32 | -0.05 | 0.0085 | 72992.88 | 1.815e-08 | 0.3176 | 0.0192 | 0.0264 | 28104 | 0.4681 |
| rs6065094 | 20 | 37453194 | A | G | 0.32 | -0.06 | 0.0082 | 73173.9 | 1.409e-14 | 0.3203 | 0.0074 | 0.0259 | 28104 | 0.7751 |
| rs61405217 | 4 | 170322312 | C | T | 0.42 | 0.05 | 0.0079 | 73173.9 | 5.387e-11 | 0.4588 | 0.0035 | 0.0247 | 28104 | 0.8876 |
| rs61789528 | 1 | 97157335 | G | A | 0.81 | -0.06 | 0.0098 | 73173.9 | 8.782e-09 | 0.8214 | -0.0191 | 0.0325 | 28104 | 0.5573 |
| rs61828917 | 1 | 173580303 | C | T | 0.85 | 0.07 | 0.011 | 73173.9 | 7.951e-10 | 0.8812 | 0.0594 | 0.0374 | 28104 | 0.1125 |
| rs61896127 | 11 | 47099867 | C | T | 0.91 | -0.10 | 0.0151 | 59339.13 | 1.238e-10 | 0.91695 | 0.0297 | 0.04353 | 28104 | 0.4951 |
| rs61937595 | 12 | 57682956 | C | T | 0.92 | 0.12 | 0.0158 | 69922.48 | 1.317e-14 | 0.9149 | 0.0096 | 0.0436 | 28104 | 0.8259 |
| rs62176172 | 2 | 185499482 | C | T | 0.64 | 0.05 | 0.0081 | 73173.9 | 5.03e-10 | 0.6814 | 0.0152 | 0.02636 | 28104 | 0.5642 |
| rs62184960 | 2 | 172974789 | C | T | 0.88 | 0.07 | 0.0122 | 73173.9 | 1.082e-08 | 0.8459 | 0.0027 | 0.0337 | 28104 | 0.9363 |
| rs634940 | 6 | 93077500 | G | T | 0.79 | -0.06 | 0.0098 | 73173.9 | 2.884e-11 | 0.7453 | -0.0659 | 0.0278 | 28104 | 0.018 |
| rs6482437 | 10 | 18726326 | A | C | 0.09 | -0.10 | 0.0135 | 73173.9 | 1.05e-14 | 0.103 | 0.0296 | 0.03876 | 28104 | 0.4451 |
| rs6500602 | 16 | 4497451 | T | C | 0.30 | 0.05 | 0.0084 | 73173.9 | 4.769e-08 | 0.2975 | 0.012 | 0.02699 | 28104 | 0.6567 |
| rs6504163 | 17 | 61545779 | C | T | 0.37 | -0.05 | 0.0082 | 73173.9 | 1.869e-09 | 0.3626 | 0.0028 | 0.02545 | 28104 | 0.9124 |
| rs6538539 | 12 | 95195293 | G | T | 0.47 | 0.05 | 0.0077 | 73173.9 | 5.634e-10 | 0.4501 | 0.001 | 0.0252 | 28104 | 0.9684 |
| rs6588168 | 1 | 66324118 | C | T | 0.48 | -0.059 | 0.0079 | 73173.9 | 5.941e-10 | 0.4433 | -0.0096 | 0.02488 | 28104 | 0.6997 |
| rs6670165 | 1 | 177280121 | C | T | 0.81 | -0.07 | 0.0098 | 73173.9 | 1.078e-11 | 0.81 | -0.0547 | 0.03119 | 28104 | 0.07949 |
| rs668437 | 3 | 135941739 | G | A | 0.42 | 0.05 | 0.0081 | 73173.9 | 9.253e-10 | 0.4735 | -0.0461 | 0.0247 | 28104 | 0.06267 |
| rs6713590 | 2 | 193937515 | C | T | 0.47 | 0.04 | 0.0077 | 73173.9 | 2.694e-08 | 0.481 | -0.0335 | 0.0245 | 28104 | 0.1721 |
| rs6715366 | 2 | 2327295 | G | A | 0.77 | -0.06 | 0.0094 | 73173.9 | 4.225e-09 | 0.7258 | 0.0401 | 0.02728 | 28104 | 0.1416 |
| rs67627854 | 2 | 76246645 | T | G | 0.82 | -0.06 | 0.0101 | 73173.9 | 2.202e-08 | 0.7936 | 0.0167 | 0.0303 | 28104 | 0.5821 |
| rs6839635 | 4 | 103872854 | C | A | 0.50 | -0.04 | 0.0079 | 73173.9 | 3.874e-08 | 0.5082 | 0.0606 | 0.0245 | 28104 | 0.01339 |
| rs6848123 | 4 | 80203425 | A | C | 0.56 | 0.05 | 0.0081 | 73173.9 | 2.054e-09 | 0.4971 | -0.0035 | 0.02454 | 28104 | 0.8866 |
| rs6919146 | 6 | 165059966 | T | G | 0.31 | -0.05 | 0.0085 | 73173.9 | 8.423e-09 | 0.3404 | -0.0242 | 0.02601 | 28104 | 0.3522 |
| rs6925079 | 6 | 64946311 | T | C | 0.64 | -0.04 | 0.0081 | 73173.9 | 3.581e-08 | 0.6802 | -0.0154 | 0.02621 | 28104 | 0.5569 |
| rs6926151 | 6 | 96476028 | A | G | 0.94 | 0.10 | 0.0176 | 59292.61 | 3.327e-08 | 0.92955 | 0.0647 | 0.04989 | 28104 | 0.1947 |
| rs6943762 | 7 | 86403263 | T | C | 0.87 | 0.10 | 0.0124 | 73173.9 | 6.296e-17 | 0.8791 | -0.0507 | 0.03707 | 28104 | 0.1715 |
| rs6969410 | 7 | 110069015 | T | G | 0.66 | 0.06 | 0.0083 | 73173.9 | 1.899e-11 | 0.6211 | -8.00e-04 | 0.02492 | 28104 | 0.9744 |
| rs6984242 | 8 | 60700469 | G | A | 0.40 | 0.05 | 0.0078 | 73173.9 | 1.5e-11 | 0.4009 | 0.0127 | 0.0249 | 28104 | 0.6103 |
| rs7002992 | 8 | 103676605 | T | C | 0.67 | 0.05 | 0.0083 | 73173.9 | 4.483e-09 | 0.6297 | 0.0116 | 0.02540 | 28104 | 0.6479 |
| rs7007361 | 8 | 111621265 | C | T | 0.84 | -0.07 | 0.0104 | 73173.9 | 4.429e-11 | 0.8274 | -0.0144 | 0.0323 | 28104 | 0.6564 |
| rs708228 | 11 | 57585662 | C | T | 0.70 | -0.05 | 0.0085 | 73173.9 | 2.062e-09 | 0.6864 | -0.0129 | 0.02617 | 28104 | 0.6221 |
| rs7112912 | 11 | 134246900 | C | T | 0.41 | 0.05 | 0.0084 | 73173.9 | 2.09e-10 | 0.4674 | -0.0502 | 0.02473 | 28104 | 0.04241 |
| rs7113068 | 11 | 124633529 | T | C | 0.86 | 0.08 | 0.0112 | 73173.9 | 4.605e-12 | 0.8367 | 0.0313 | 0.03369 | 28104 | 0.3529 |
| rs715170 | 18 | 53795514 | C | T | 0.75 | 0.06 | 0.009 | 73173.9 | 7.4e-13 | 0.743 | -0.0392 | 0.0277 | 28104 | 0.1584 |
| rs7177338 | 15 | 91428636 | G | A | 0.42 | -0.07 | 0.0082 | 73173.9 | 2.024e-19 | 0.4703 | -8.00e-04 | 0.0243 | 28104 | 0.9738 |
| rs7178152 | 15 | 89787055 | T | G | 0.53 | 0.05 | 0.0078 | 73173.9 | 2.628e-10 | 0.5362 | -0.0302 | 0.0244 | 28104 | 0.2177 |
| rs717947 | 4 | 33653757 | C | T | 0.87 | -0.07 | 0.0116 | 73173.9 | 3.096e-09 | 0.90095 | -0.0537 | 0.0430 | 28104 | 0.2121 |
| rs7207904 | 17 | 1267857 | G | A | 0.61 | 0.05 | 0.0079 | 73173.9 | 1.645e-09 | 0.6194 | 0.0155 | 0.0252 | 28104 | 0.5394 |
| rs7238071 | 18 | 77579812 | A | G | 0.69 | -0.0 | 0.0084 | 73173.9 | 9.292e-14 | 0.7263 | -0.0452 | 0.02797 | 28104 | 0.1062 |
| rs72728416 | 1 | 97834691 | A | G | 0.73 | -0.06 | 0.0087 | 73173.9 | 4.99e-12 | 0.7566 | -0.0042 | 0.0285 | 28104 | 0.8829 |
| rs72761691 | 9 | 134786548 | A | C | 0.86 | -0.07 | 0.0116 | 73173.9 | 1.07e-08 | 0.8598 | 0.0053 | 0.03540 | 28104 | 0.881 |
| rs72769124 | 1 | 239210058 | C | A | 0.91 | -0.10 | 0.0143 | 73173.9 | 4.36e-13 | 0.90654 | 0.0167 | 0.04214 | 28104 | 0.6919 |
| rs72974269 | 2 | 225454907 | C | T | 0.68 | 0.05 | 0.0083 | 73173.9 | 2.76e-10 | 0.695 | -0.0494 | 0.02648 | 28104 | 0.06219 |
| rs7312697 | 12 | 29933069 | T | C | 0.36 | -0.05 | 0.0081 | 73173.9 | 4.85e-11 | 0.3745 | -0.0139 | 0.0253 | 28104 | 0.5838 |
| rs73229090 | 8 | 27442127 | C | A | 0.89 | 0.10 | 0.0142 | 58749.13 | 4.34e-13 | 0.8933 | 0.0578 | 0.03875 | 28104 | 0.1359 |
| rs74695911 | 7 | 131541469 | T | C | 0.82 | 0.06 | 0.0103 | 73173.9 | 4.897e-08 | 0.784 | 0.0373 | 0.02998 | 28104 | 0.2135 |
| rs7563610 | 2 | 37566376 | A | G | 0.58 | 0.05 | 0.0079 | 73173.9 | 4.48e-10 | 0.6052 | -0.0307 | 0.02504 | 28104 | 0.2202 |
| rs758749 | 19 | 57189718 | C | T | 0.86 | -0.06 | 0.0112 | 73173.9 | 4.66e-8 | 0.8696 | 0.0116 | 0.03733 | 28104 | 0.756 |
| rs7600146 | 2 | 22543347 | G | T | 0.63 | 0.05 | 0.0083 | 73173.9 | 8.311e-09 | 0.569 | -0.013 | 0.02485 | 28104 | 0.601 |
| rs7609876 | 3 | 176790116 | T | C | 0.72 | -0.05 | 0.0089 | 73173.9 | 9.398e-9 | 0.7932 | 0.0313 | 0.03031 | 28104 | 0.3018 |
| rs7632834 | 3 | 17886678 | T | C | 0.42 | -0.05 | 0.0085 | 73173.9 | 2.091e-9 | 0.5062 | -0.0099 | 0.02461 | 28104 | 0.6875 |
| rs7647398 | 3 | 180733150 | C | T | 0.82 | 0.09 | 0.01 | 73173.9 | 2.207e-17 | 0.8063 | 0.0122 | 0.03067 | 28104 | 0.6908 |
| rs7703010 | 5 | 137711716 | C | A | 0.37 | 0.05 | 0.0082 | 73173.9 | 8.278e-09 | 0.4127 | -0.0028 | 0.0247 | 28104 | 0.9101 |
| rs7703618 | 5 | 44914579 | G | A | 0.44 | 0.05 | 0.008 | 73173.9 | 3.408e-09 | 0.3793 | -0.0051 | 0.02550 | 28104 | 0.8415 |
| rs7706116 | 5 | 45742624 | G | T | 0.56 | -0.06 | 0.0078 | 73173.9 | 6.786e-13 | 0.5612 | -0.0115 | 0.02502 | 28104 | 0.6458 |
| rs77154381 | 2 | 27982119 | A | G | 0.68 | 0.05 | 0.0086 | 73173.9 | 2.6e-08 | 0.7468 | 0.0184 | 0.02833 | 28104 | 0.5161 |
| rs778371 | 2 | 233743109 | A | G | 0.746 | -0.05 | 0.0089 | 73173.9 | 1.103e-16 | 0.7248 | 0.016 | 0.02743 | 28104 | 0.5597 |
| rs7811417 | 7 | 21534152 | T | C | 0.35 | 0.05 | 0.0081 | 73173.9 | 2.166e-09 | 0.33 | -8,00E-04 | 0.02681 | 28104 | 0.9762 |
| rs7816998 | 8 | 38257506 | G | A | 0.77 | 0.06 | 0.0092 | 73173.9 | 3.105e-10 | 0.7789 | 0.019 | 0.02893 | 28104 | 0.5114 |
| rs7915131 | 10 | 64418656 | C | T | 0.42 | 0.04 | 0.0078 | 73173.9 | 4.943e-08 | 0.4319 | 0.051 | 0.02469 | 28104 | 0.03894 |
| rs79210963 | 7 | 24717969 | T | C | 0.90 | -0.09 | 0.0129 | 73173.9 | 2.584e-11 | 0.8922 | -0.0214 | 0.03880 | 28104 | 0.5813 |
| rs7927176 | 11 | 123395864 | A | G | 0.66 | -0.05 | 0.0081 | 73173.9 | 6.604e-11 | 0.6617 | 0.0139 | 0.02606 | 28104 | 0.5938 |
| rs8055219 | 16 | 13753384 | G | A | 0.79 | -0.07 | 0.0095 | 73173.9 | 1.565e-12 | 0.7785 | -0.0525 | 0.0295 | 28104 | 0.07584 |
| rs8104557 | 19 | 31030189 | T | C | 0.82 | -0.06 | 0.011 | 61674.31 | 3.764e-08 | 0.8023 | -0.0594 | 0.03067 | 28104 | 0.05282 |
| rs8180995 | 8 | 143326237 | A | G | 0.53 | 0.06 | 0.0078 | 73173.9 | 2.19e-15 | 0.5216 | 0.0405 | 0.0246 | 28104 | 0.1005 |
| rs832195 | 3 | 63864612 | T | C | 0.34 | 0.05 | 0.0083 | 73173.9 | 2.969e-09 | 0.2972 | 0.0593 | 0.0271 | 28104 | 0.02872 |
| rs9257802 | 6 | 29343355 | C | T | 0.83 | 0.09 | 0.0107 | 70162.94 | 1.182e-15 | 0.8034 | 0.0493 | 0.0306 | 28104 | 0.108 |
| rs9287971 | 2 | 174931752 | G | A | 0.60 | -0.05 | 0.0083 | 73173.9 | 3.824e-08 | 0.671 | 0.0407 | 0.026 | 28104 | 0.131 |
| rs9304548 | 18 | 27500959 | C | A | 0.26 | 0.06 | 0.0089 | 73173.9 | 1.895e-11 | 0.2623 | 0.0041 | 0.0278 | 28104 | 0.8829 |
| rs9318638 | 13 | 79986153 | T | C | 0.43 | -0.05 | 0.0079 | 73173.9 | 2.044e-09 | 0.477 | 0.0169 | 0.0245 | 28104 | 0.4914 |
| rs9393741 | 6 | 26654189 | A | G | 0.82 | 0.09 | 0.0102 | 73173.9 | 5.897e-19 | 0.7878 | -0.0024 | 0.029 | 28104 | 0.9351 |
| rs9454727 | 6 | 70003389 | A | G | 0.77 | 0.05 | 0.0098 | 70865.66 | 1.93e-08 | 0.7358 | 0.0164 | 0.02774 | 28104 | 0.5544 |
| rs9569820 | 13 | 58702746 | G | T | 0.84 | -0.07 | 0.0109 | 73173.9 | 6.563e-10 | 0.8836 | 0.0254 | 0.03889 | 28104 | 0.5137 |
| rs9597388 | 13 | 56928696 | G | A | 0.82 | 0.07 | 0.0101 | 73173.9 | 3.239e-11 | 0.8067 | 0.0044 | 0.03113 | 28104 | 0.8876 |
| rs9623320 | 22 | 41478482 | G | A | 0.76 | -0.06 | 0.0092 | 73173.9 | 2.371e-10 | 0.7122 | -0.045 | 0.0270 | 28104 | 0.09665 |
| rs967005 | 6 | 28210688 | C | T | 0.83 | 0.07 | 0.0102 | 73173.9 | 1.3e-11 | 0.8046 | 0.0183 | 0.03081 | 28104 | 0.5526 |
| rs9803993 | 1 | 243672125 | G | A | 0.78 | 0.05 | 0.0093 | 73173.9 | 1.497e-08 | 0.7811 | 0.0242 | 0.02959 | 28104 | 0.4136 |
| rs9831201 | 3 | 80913228 | T | G | 0.72 | 0.054 | 0.0087 | 73173.9 | 6.7e-10 | 0.6993 | -0.0132 | 0.02657 | 28104 | 0.6194 |
| rs9882532 | 3 | 16865845 | T | C | 0.6 | -0.0 | 0.0087 | 73173.9 | 8.574e-10 | 0.6518 | -0.0494 | 0.02587 | 28104 | 0.05623 |
| rs9891739 | 17 | 19942177 | C | T | 0.54 | -0.04 | 0.008 | 73173.9 | 2.181e-08 | 0.4702 | -0.0815 | 0.02449 | 28104 | 0.0008751 |
| rs9926296 | 16 | 89818089 | A | G | 0.44 | 0.05 | 0.0086 | 71743.28 | 2.374e-08 | 0.49 | 0.0386 | 0.02457 | 28104 | 0.1162 |
| rs9975024 | 21 | 16439883 | A | G | 0.53 | -0.05 | 0.008 | 73173.9 | 1.777e-09 | 0.5254 | 0.012 | 0.02445 | 28104 | 0.6237 |
| rs999494 | 2 | 73157395 | C | T | 0.82 | 0.06 | 0.0101 | 73173.9 | 1.044e-08 | 0.7975 | -0.0032 | 0.03070 | 28104 | 0.917 |

Instrumental SNPs select for the SCZ to BAG classical Mendelian randomization analysis. CHR: chromosome; POS: hg19 genomic position; A1: effective allele; A2: the other allele; A1_exp_frq: A1 frequency in the exposure dataset; Beta_exp, effect of A1 for the exposure; SE_exp; standard error for Beta_exp; N_exp: sample size for exposure; P_exp, association p value for exposure; A1_out_frq: A1 frequency in the outcome dataset; Beta_out: effect size of A1 on outcome; SE_out: standard error of Beta_out; N_out: sample size for outcome; P_out: association p value for outcome.

### **Table S12. Harmonized instrumental SNPs (p<5x10^-8^) for the BIP to BAG MR.**

| SNP | CHR | POS | A1 | A2 | A1_exp_frq | Beta_exp | SE_exp | N_exp | P_exp | A1_out_frq | Beta_out | SE_out | N_out | P_out |
| --- | --- | --- | --- | --- | --- | --- | --- | --- | --- | --- | --- | --- | --- | --- |
| rs10043984 | 5 | 137712121 | C | T | 0.76 | -0.06 | 0.0108 | 50981.48 | 3.709e-08 | 0.7468 | -0.0202 | 0.0277 | 28104 | 0.467 |
| rs10994299 | 10 | 62076628 | C | A | 0.95 | -0.13 | 0.0201 | 50862.93 | 9.587e-11 | 0.94999 | 0.0062 | 0.0577 | 28104 | 0.9145 |
| rs113779084 | 7 | 11871787 | G | A | 0.70 | -0.08 | 0.0102 | 50981.48 | 1.423e-13 | 0.7099 | -0.0301 | 0.0273 | 28104 | 0.2716 |
| rs11647445 | 16 | 9926966 | T | G | 0.67 | -0.05 | 0.0098 | 50981.48 | 2.152e-08 | 0.6472 | -0.0169 | 0.0257 | 28104 | 0.5109 |
| rs11764361 | 7 | 105043229 | A | G | 0.67 | 0.06 | 0.0104 | 50981.48 | 3.471e-09 | 0.6565 | -0.0357 | 0.0258 | 28104 | 0.1674 |
| rs11856299 | 15 | 83534421 | C | T | 0.75 | 0.06 | 0.011 | 50981.48 | 1.528e-08 | 0.7649 | -0.0026 | 0.0295 | 28104 | 0.9298 |
| rs12289486 | 11 | 79092527 | C | T | 0.89 | -0.08 | 0.0149 | 50981.48 | 3.297e-08 | 0.892 | 0.0176 | 0.0389 | 28104 | 0.6517 |
| rs1235162 | 6 | 29537224 | A | G | 0.91 | 0.11 | 0.0164 | 49000.67 | 6.899e-12 | 0.8804 | 0.0822 | 0.03623 | 28104 | 0.02328 |
| rs12575685 | 11 | 70517927 | G | A | 0.67 | -0.07 | 0.0101 | 50981.48 | 1.243e-10 | 0.7012 | -0.0301 | 0.02643 | 28104 | 0.2549 |
| rs12668848 | 7 | 2020995 | G | A | 0.58 | 0.06 | 0.0095 | 50981.48 | 1.902e-09 | 0.5709 | 0.0043 | 0.0247 | 28104 | 0.8618 |
| rs12672003 | 7 | 24647222 | A | G | 0.89 | -0.09 | 0.0155 | 46163.71 | 2.718e-09 | 0.8924 | -0.0139 | 0.03892 | 28104 | 0.721 |
| rs13044225 | 20 | 60865815 | A | G | 0.56 | -0.05 | 0.0095 | 50186.29 | 8.498e-09 | 0.5319 | 0.0124 | 0.0244 | 28104 | 0.6127 |
| rs13195401 | 6 | 26463574 | G | T | 0.92 | 0.14 | 0.0175 | 50615.04 | 6.979e-15 | 0.90102 | 0.039 | 0.0389 | 28104 | 0.3163 |
| rs1487445 | 6 | 98565211 | C | T | 0.51 | -0.07 | 0.0093 | 50981.48 | 1.484e-15 | 0.5324 | 0.0098 | 0.02467 | 28104 | 0.6913 |
| rs17183814 | 2 | 166152389 | G | A | 0.92 | 0.10 | 0.0185 | 50772.41 | 2.678e-08 | 0.92605 | 0.0694 | 0.0464 | 28104 | 0.1354 |
| rs17202645 | 2 | 169399768 | C | T | 0.81 | 0.07 | 0.0121 | 50981.48 | 2.494e-08 | 0.8177 | -0.0273 | 0.0319 | 28104 | 0.3923 |
| rs174581 | 11 | 61606683 | G | A | 0.63 | -0.07 | 0.0097 | 50981.48 | 4.944e-13 | 0.6495 | -0.0509 | 0.0256 | 28104 | 0.04681 |
| rs17526722 | 6 | 25918855 | G | A | 0.93 | 0.10 | 0.0181 | 50615.04 | 1.759e-08 | 0.91019 | 0.0409 | 0.04075 | 28104 | 0.3156 |
| rs17693963 | 6 | 27710165 | A | C | 0.91 | 0.12 | 0.0161 | 50772.41 | 5.836e-14 | 0.8858 | 0.0283 | 0.036 | 28104 | 0.4431 |
| rs1894401 | 15 | 91429042 | G | A | 0.46 | -0.05 | 0.0096 | 50035.72 | 2.799e-08 | 0.4709 | 1,00E-04 | 0.01899 | 28104 | 0.9958 |
| rs1998821 | 10 | 18751242 | G | A | 0.89 | 0.08 | 0.0152 | 50981.48 | 4.25e-08 | 0.8948 | -0.0299 | 0.038 | 28104 | 0.4395 |
| rs2019611 | 15 | 85122620 | C | A | 0.75 | 0.07 | 0.0108 | 50981.48 | 1.437e-10 | 0.7509 | -0.0423 | 0.0282 | 28104 | 0.1338 |
| rs2069772 | 4 | 123373133 | T | C | 0.73 | -0.06 | 0.0103 | 50981.48 | 1.567e-08 | 0.7318 | -0.0386 | 0.0275 | 28104 | 0.1616 |
| rs2126180 | 1 | 61105668 | G | A | 0.54 | -0.06 | 0.0094 | 50981.48 | 1.621e-09 | 0.5298 | -3,00E-04 | 0.0232 | 28104 | 0.9897 |
| rs2159100 | 12 | 2346393 | C | T | 0.67 | -0.08 | 0.0098 | 50981.48 | 1.872e-15 | 0.6682 | -0.04 | 0.02614 | 28104 | 0.126 |
| rs237475 | 20 | 48049506 | T | C | 0.52 | -0.05 | 0.0093 | 50981.48 | 5.171e-09 | 0.5201 | 0.0134 | 0.02457 | 28104 | 0.5856 |
| rs2577831 | 3 | 52628056 | C | A | 0.51 | 0.07 | 0.0093 | 50981.48 | 3.834e-13 | 0.5291 | -0.003 | 0.02414 | 28104 | 0.9011 |
| rs2719164 | 2 | 194437889 | A | G | 0.57 | 0.05 | 0.0095 | 50981.48 | 4.847e-08 | 0.5763 | -0.011 | 0.02488 | 28104 | 0.6585 |
| rs28565152 | 5 | 7542911 | G | A | 0.767 | -0.07 | 0.0112 | 48268.74 | 1.963e-09 | 0.7557 | -0.0354 | 0.02874 | 28104 | 0.2181 |
| rs2899059 | 15 | 42844361 | C | T | 0.927 | -0.1 | 0.017 | 50981.48 | 1.243e-08 | 0.91467 | 0.0753 | 0.04554 | 28104 | 0.09826 |
| rs2953928 | 8 | 34152492 | G | A | 0.937 | -0.12 | 0.02 | 50615.04 | 6.249e-09 | 0.94541 | -0.0462 | 0.05104 | 28104 | 0.3654 |
| rs3094035 | 6 | 30363136 | G | T | 0.907 | 0 1 | 0.0153 | 49000.67 | 4.168e-10 | 0.8645 | 0.0588 | 0.03447 | 28104 | 0.0881 |
| rs3115674 | 6 | 31799076 | T | G | 0.897 | 0.08 | 0.0154 | 48295.99 | 3.871e-08 | 0.8713 | 0.0882 | 0.03535 | 28104 | 0.01261 |
| rs3130557 | 6 | 31094703 | C | T | 0.89 | 0.09 | 0.0147 | 50862.93 | 9.734e-11 | 0.8564 | 0.074 | 0.033624 | 28104 | 0.02775 |
| rs35741362 | 6 | 27007687 | T | C | 0.89 | 0.10 | 0.0148 | 50862.93 | 1.366e-12 | 0.8638 | 0.0135 | 0.0347 | 28104 | 0.6978 |
| rs35958438 | 15 | 38973793 | G | A | 0.77 | 0.0 | 0.0117 | 47077.45 | 3.83e-08 | 0.7742 | 0.0192 | 0.0293 | 28104 | 0.5131 |
| rs3856207 | 1 | 163747095 | A | G | 0.45 | 0.05 | 0.0095 | 50186.29 | 1.156e-08 | 0.4603 | -0.0053 | 0.0244 | 28104 | 0.8284 |
| rs39824 | 5 | 169255522 | G | A | 0.33 | -0.06 | 0.0099 | 50981.48 | 9.568e-09 | 0.343 | 0.0217 | 0.0258 | 28104 | 0.4018 |
| rs4072458 | 3 | 36862680 | T | C | 0.46 | -0.08 | 0.0094 | 50981.48 | 5.233e-17 | 0.4529 | 0.0167 | 0.0246 | 28104 | 0.4973 |
| rs41315395 | 6 | 32201469 | C | A | 0.86 | -0.07 | 0.0127 | 49209.73 | 1.476e-08 | 0.8502 | 0.0314 | 0.03501 | 28104 | 0.3698 |
| rs4619651 | 2 | 97416153 | G | A | 0.67 | 0.07 | 0.0101 | 50981.48 | 4.776e-11 | 0.6917 | -0.0239 | 0.02633 | 28104 | 0.3642 |
| rs4672 | 11 | 64009879 | G | A | 0.92 | -0.10 | 0.0172 | 50035.72 | 3.42e-09 | 0.92068 | -0.1075 | 0.04518 | 28104 | 0.01735 |
| rs4790841 | 17 | 1835482 | C | T | 0.85 | -0.07 | 0.0132 | 50035.72 | 3.141e-08 | 0.8462 | 0.0367 | 0.03379 | 28104 | 0.2775 |
| rs531490 | 11 | 66324360 | T | G | 0.46 | 0.05 | 0.0093 | 50981.48 | 6.016e-09 | 0.456 | -0.0145 | 0.02441 | 28104 | 0.5526 |
| rs5758064 | 22 | 41153879 | T | C | 0.52 | 0.05 | 0.0093 | 50981.48 | 2.011e-08 | 0.5221 | 0.029 | 0.02474 | 28104 | 0.2413 |
| rs61554907 | 17 | 38220432 | G | T | 0.88 | -0.09 | 0.0154 | 50035.72 | 1.636e-08 | 0.8947 | -0.0842 | 0.0395 | 28104 | 0.03331 |
| rs67712855 | 20 | 43682551 | T | G | 0.69 | 0.07 | 0.0103 | 50186.29 | 4.221e-11 | 0.7129 | 0.0039 | 0.02710 | 28104 | 0.8856 |
| rs6806239 | 3 | 70488207 | T | G | 0.80 | 0.07 | 0.0119 | 50981.48 | 2.641e-08 | 0.8163 | -0.0315 | 0.03145 | 28104 | 0.3166 |
| rs6865469 | 5 | 78849505 | G | T | 0.72 | -0.06 | 0.0103 | 50981.48 | 1.65e-08 | 0.7309 | -0.0366 | 0.02798 | 28104 | 0.1909 |
| rs6887473 | 5 | 80961069 | G | A | 0.74 | 0.06 | 0.0105 | 50981.48 | 8.814e-09 | 0.7178 | -0.0204 | 0.0272 | 28104 | 0.4546 |
| rs6954854 | 7 | 21492589 | G | A | 0.43 | 0.06 | 0.0094 | 50981.48 | 5.936e-10 | 0.426 | -0.0075 | 0.0249 | 28104 | 0.7634 |
| rs7461753 | 8 | 144981498 | A | C | 0.6 | -0.06 | 0.0098 | 50981.48 | 5.24e-09 | 0.5842 | -0.0182 | 0.02501 | 28104 | 0.4668 |
| rs763193 | 6 | 166996127 | A | G | 0.51 | -0.0 | 0.0094 | 50981.48 | 1.192e-08 | 0.5185 | -0.0142 | 0.0248 | 28104 | 0.5676 |
| rs7813581 | 8 | 9882652 | G | T | 0.83 | -0.08 | 0.0119 | 50981.48 | 9.911e-11 | 0.8286 | 0.0275 | 0.0325 | 28104 | 0.3988 |
| rs9393926 | 6 | 28682725 | G | A | 0.09 | -0.11 | 0.0165 | 49000.67 | 7.501e-12 | 0.1176 | -0.031 | 0.0367 | 28104 | 0.3987 |

Instrumental SNPs select for the BIP to BAG classical Mendelian randomization analysis. CHR: chromosome; POS: hg19 genomic position; A1: effective allele; A2: the other allele; A1_exp_frq: A1 frequency in the exposure dataset; Beta_exp, effect of A1 for the exposure; SE_exp; standard error for Beta_exp; N_exp: sample size for exposure; P_exp, association p value for exposure; A1_out_frq: A1 frequency in the outcome dataset; Beta_out: effect size of A1 on outcome; SE_out: standard error of Beta_out; N_out: sample size for outcome; P_out: association p value for outcome.

### **Table S13. Harmonized instrumental SNPs (p<5x10^-8^) for the MDD to BAG MR.**

| SNP | CHR | POS | A1 | A2 | A1_exp_frq | Beta_exp | SE_exp | N_exp | P_exp | A1_out_frq | Beta_out | SE_out | N_out | P_out |
| --- | --- | --- | --- | --- | --- | --- | --- | --- | --- | --- | --- | --- | --- | --- |
| rs12658032 | 5 | 103904226 | A | G | 0.36 | 0.05 | 0.0083 | 69115.85 | 1.181e-10 | 0.3497 | 0.0225 | 0.0257 | 28104 | 0.3804 |
| rs1460943 | 1 | 72813129 | T | C | 0.61 | 0.04 | 0.0081 | 69115.85 | 4.27e-8 | 0.609 | -0.0145 | 0.0249 | 28104 | 0.5611 |
| rs148696809 | 6 | 28934352 | T | C | 0.90 | 0.08 | 0.0132 | 69115.85 | 4.355e-9 | 0.8902 | 0.0412 | 0.037 | 28104 | 0.2712 |
| rs2799077 | 6 | 28234597 | T | C | 0.15 | -0.07 | 0.0112 | 69115.85 | 2.864e-9 | 0.1637 | -0.0213 | 0.0327 | 28104 | 0.5153 |
| rs4593766 | 1 | 73773043 | T | C | 0.38 | 0.05 | 0.0081 | 69115.85 | 1.705e-8 | 0.3694 | 0.0147 | 0.0254 | 28104 | 0.563 |
| rs58825580 | 6 | 26365679 | T | G | 0.88 | 0.07 | 0.012 | 69115.85 | 1.258e-8 | 0.8664 | 0.0422 | 0.0349 | 28104 | 0.2272 |
| rs6934329 | 6 | 27158033 | A | G | 0.82 | 0.06 | 0.0104 | 69115.85 | 1.416e-9 | 0.8128 | -0.008 | 0.0308 | 28104 | 0.7952 |
| rs6940116 | 6 | 27708732 | A | G | 0.84 | 0.06 | 0.0107 | 69115.85 | 4.912e-8 | 0.8228 | -0.0048 | 0.0316 | 28104 | 0.8795 |
| rs8013655 | 14 | 42089908 | T | C | 0.47 | 0.04 | 0.0079 | 69115.85 | 2.672e-8 | 0.4774 | 0.0518 | 0.0247 | 28104 | 0.03578 |

Instrumental SNPs select for the MDD to BAG classical Mendelian randomization analysis. CHR: chromosome; POS: hg19 genomic position; A1: effective allele; A2: the other allele; A1_exp_frq: A1 frequency in the exposure dataset; Beta_exp, effect of A1 for the exposure; SE_exp; standard error for Beta_exp; N_exp: sample size for exposure; P_exp, association p value for exposure; A1_out_frq: A1 frequency in the outcome dataset; Beta_out: effect size of A1 on outcome; SE_out: standard error of Beta_out; N_out: sample size for outcome; P_out: association p value for outcome.

### **Table S14. Harmonized instrumental SNPs (p<5x10^-8^) for the AD to BAG MR.**

| SNP | CHR | POS | A1 | A2 | A1_exp_frq | Beta_exp | SE_exp | N_exp | P_exp | A1_out_frq | Beta_out | SE_out | N_out | P_out |
| --- | --- | --- | --- | --- | --- | --- | --- | --- | --- | --- | --- | --- | --- | --- |
| rs10412413 | 19 | 45327309 | T | C | 0.3 | 0.0526 | 0.0023 | 429332.77 | 6.97e-115 | 0.32 | 0.0203 | 0.0264 | 28104 | 0.44 |
| rs10808026 | 7 | 143099133 | A | C | 0.202 | -0.0161 | 0.0027 | 425571.98 | 2.82e-9 | 0.22 | -0.0219 | 0.03 | 28104 | 0.47 |
| rs11257240 | 10 | 11719074 | G | T | 0.3 | 0.0129 | 0.0023 | 423921.06 | 1.28e-8 | 0.36 | -0.0021 | 0.0252 | 28104 | 0.93 |
| rs113260531 | 17 | 5138980 | A | G | 0.13 | 0.02 | 0.0033 | 429194.03 | 7.91e-10 | 0.12 | 0.0018 | 0.0376 | 28104 | 0.96 |
| rs11672748 | 19 | 45490192 | G | A | 0.34= | 0.023 | 0.0023 | 427777.91 | 7.25e-24 | 0.32 | -0.026 | 0.0263 | 28104 | 0.32 |
| rs117618017 | 15 | 63569902 | T | C | 0.12= | 0.018 | 0.0033 | 428753.23 | 3.44e-8 | 0.14 | 0.0015 | 0.0345 | 28104 | 0.97 |
| rs12539172 | 7 | 100091795 | T | C | 0.31= | -0.0167 | 0.0023 | 429961.08 | 5.85e-13 | 0.32 | -0.0605 | 0.0261 | 28104 | 2.06e-2 |
| rs12590654 | 14 | 92938855 | A | G | 0.34 | -0.0148 | 0.0023 | 420451.85 | 1.32e-10 | 0.34 | -0.0367 | 0.0263 | 28104 | 0.16 |
| rs1532278 | 8 | 27466315 | T | C | 0.38 | -0.0201 | 0.0022 | 428741.91 | 2.07e-19 | 0.39 | 0.0092 | 0.0251 | 28104 | 0.714 |
| rs2081545 | 11 | 59958380 | A | C | 0.3 | -0.0179 | 0.0022 | 425881.39 | 1.11e-15 | 0.40 | -0.0624 | 0.025 | 28104 | 1.23e-2 |
| rs2647066 | 6 | 32571122 | T | C | 0.13 | -0.0208 | 0.0032 | 428954.21 | 8.55e-11 | 0.18 | 0.0101 | 0.0314 | 28104 | 0.75 |
| rs28394864 | 17 | 47450775 | A | G | 0.45 | 0.0123 | 0.0022 | 424326.36 | 1.68e-8 | 0.45 | 0.0749 | 0.0248 | 28104 | 2.50e-3 |
| rs28469095 | 19 | 45655333 | C | T | 0.1 | 0.0466 | 0.0036 | 426787.79 | 1.07e-38 | 0.09 | 0.0769 | 0.0427 | 28104 | 7.17e-2 |
| rs28834970 | 8 | 27195121 | C | T | 0.37 | 0.0153 | 0.0022 | 428248.62 | 8.49e-12 | 0.36 | 0.0151 | 0.0254 | 28104 | 0.55 |
| rs3752231 | 19 | 1043638 | T | C | 0.25 | 0.0159 | 0.0025 | 424712.83 | 2.38e-10 | 0.25 | 0.0665 | 0.0284 | 28104 | 1.93e-2 |
| rs3810140 | 19 | 45316807 | T | C | 0.07 | -0.0286 | 0.0043 | 424090.94 | 4.54e-11 | 0.06 | 0.0105 | 0.0562 | 28104 | 0.85 |
| rs412776 | 19 | 45379516 | A | G | 0.11 | 0.02834 | 0.0035 | 428640.06 | 8.10e-16 | 0.12 | -0.023 | 0.0379 | 28104 | 0.54 |
| rs4575098 | 1 | 161155392 | A | G | 0.23 | 0.01641 | 0.0026 | 427808.62 | 1.90e-10 | 0.23 | 0.0763 | 0.0291 | 28104 | 8.69e-3 |
| rs4663105 | 2 | 127891427 | C | A | 0.43 | 0.0311 | 0.0022 | 418699.97 | 1.45e-44 | 0.42 | -0.0275 | 0.0252 | 28104 | 0.28 |
| rs4803750 | 19 | 45247627 | G | A | 0.06 | -0.0491 | 0.0044 | 427688.71 | 1.61e-28 | 0.07 | 0.0258 | 0.0481 | 28104 | 0.59 |
| rs59007384 | 19 | 45396665 | T | G | 0.2 | 0.1389 | 0.0027 | 427829.43 | 1,00e-200 | 0.21 | 0.103 | 0.0306 | 28104 | 7.57e-4 |
| rs593742 | 15 | 59045774 | G | A | 0.31 | -0.0138 | 0.0023 | 426926.64 | 3.73e-9 | 0.31 | 0.0281 | 0.0267 | 28104 | 0.29 |
| rs6014724 | 20 | 54998544 | G | A | 0.1 | -0.0229 | 0.0037 | 427344.83 | 5.38e-10 | 0.09 | 0.0096 | 0.0433 | 28104 | 0.82 |
| rs6448453 | 4 | 11026028 | A | G | 0.26 | 0.0147 | 0.0025 | 429961.08 | 1.98e-9 | 0.26 | 0.032 | 0.0276 | 28104 | 0.25 |
| rs679515 | 1 | 207750568 | T | C | 0.17 | 0.0254 | 0.0027 | 429226.74 | 6.83e-19 | 0.17 | 0.0781 | 0.032 | 28104 | 1.47e-2 |
| rs74862042 | 19 | 45217784 | C | A | 0.161 | 0.0281 | 0.0029 | 429642.96 | 9.11e-22 | 0.15 | 0.0142 | 0.0342 | 28104 | 0.68 |
| rs846881 | 19 | 45078553 | C | A | 0.21 | -0.0174 | 0.0027 | 422334.58 | 9.89e-11 | 0.21 | -0.0278 | 0.0301 | 28104 | 0.36 |
| rs867611 | 11 | 85776544 | G | A | 0.32 | -0.0204 | 0.0023 | 427263.57 | 1.48e-18 | 0.31 | 0.0186 | 0.0263 | 28104 | 0.48 |
| rs9381563 | 6 | 47432637 | C | T | 0.36 | 0.0145 | 0.0023 | 422732.93 | 1.99e-10 | 0.37 | 0.0211 | 0.0257 | 28104 | 0.41 |
| rs9653111 | 19 | 45592475 | T | C | 0.11 | -0.0194 | 0.0034 | 427366.64 | 1.22e-8 | 0.12 | 0.0228 | 0.0364 | 28104 | 0.53 |

Instrumental SNPs select for the AD to BAG classical Mendelian randomization analysis. CHR: chromosome; POS: hg19 genomic position; A1: effective allele; A2: the other allele; A1_exp_frq: A1 frequency in the exposure dataset; Beta_exp, effect of A1 for the exposure; SE_exp; standard error for Beta_exp; N_exp: sample size for exposure; P_exp, association p value for exposure; A1_out_frq: A1 frequency in the outcome dataset; Beta_out: effect size of A1 on outcome; SE_out: standard error of Beta_out; N_out: sample size for outcome; P_out: association p value for outcome.

### **Table S15. Harmonized instrumental SNPs (p<5x10^-8^) for the PD to BAG MR.**

| SNP | CHR | POS | A1 | A2 | A1_exp_frq | Beta_exp | SE_exp | N_exp | P_exp | A1_out_frq | Beta_out | SE_out | N_out | P_out |
| --- | --- | --- | --- | --- | --- | --- | --- | --- | --- | --- | --- | --- | --- | --- |
| rs10513789 | 3 | 182760073 | T | G | 0.82 | 0.1596 | 0.0219 | 62649 | 3.185e-13 | 0.80 | 0.0217 | 0.0312 | 28104 | 0.4874 |
| rs11942911 | 4 | 90601310 | A | G | 0.92 | -0.1872 | 0.0331 | 62649 | 1.566e-08 | 0.92 | 0.0745 | 0.0474 | 28104 | 0.1162 |
| rs12817488 | 12 | 123296294 | A | G | 0.46 | 0.1032 | 0.0178 | 62649 | 7.111e-09 | 0.46 | 0.0452 | 0.0248 | 28104 | 0.06877 |
| rs1372518 | 4 | 90757294 | A | C | 0.21 | -0.2334 | 0.0209 | 62649 | 5.354e-29 | 0.22 | 0.0511 | 0.0299 | 28104 | 0.08718 |
| rs1474055 | 2 | 169110394 | T | C | 0.13 | 0.1763 | 0.0248 | 62649 | 1.144e-12 | 0.12 | -0.0547 | 0.0385 | 28104 | 0.1559 |
| rs329652 | 11 | 133769699 | A | G | 0.36 | 0.1082 | 0.0174 | 62649. | 5.027e-10 | 0.35 | -0.0014 | 0.0256 | 28104 | 0.9565 |
| rs34311866 | 4 | 951947 | T | C | 0.80 | -0.2272 | 0.0231 | 62649 | 7.974e-23 | 0.83 | -0.0409 | 0.0324 | 28104 | 0.207 |
| rs35118762 | 6 | 32560631 | T | C | 0.16 | -0.2042 | 0.0312 | 60613 | 6.026e-11 | 0.18 | 0.0123 | 0.0312 | 28104 | 0.6943 |
| rs356175 | 4 | 90630814 | T | C | 0.66 | -0.1538 | 0.0181 | 62649 | 1.607e-17 | 0.68 | -0.0033 | 0.0261 | 28104 | 0.8997 |
| rs4566208 | 17 | 16010920 | A | G | 0.43 | 0.0957 | 0.0174 | 62649 | 3.884e-08 | 0.44 | 0.0318 | 0.0246 | 28104 | 0.1967 |
| rs4588066 | 18 | 40672964 | A | G | 0.33 | 0.1046 | 0.0178 | 62649 | 4.453e-09 | 0.32 | 0.0147 | 0.0264 | 28104 | 0.5781 |
| rs4631042 | 4 | 15712550 | A | C | 0.53 | 0.1213 | 0.0169 | 62649 | 6.64e-13 | 0.52 | 0.0055 | 0.0249 | 28104 | 0.8254 |
| rs4774417 | 15 | 61993702 | A | G | 0.74 | 0.1052 | 0.0192 | 62649 | 4.626e-08 | 0.73 | -0.0355 | 0.0282 | 28104 | 0.2081 |
| rs56236914 | 17 | 43483551 | T | C | 0.19 | -0.2147 | 0.0258 | 62649 | 8.739e-17 | 0.17 | 0.2877 | 0.0316 | 28104 | 9.633e-20 |
| rs620490 | 8 | 16697579 | T | G | 0.72 | 0.1174 | 0.019 | 62649 | 6.456e-10 | 0.752 | 0.0072 | 0.0270 | 28104 | 0.79 |
| rs7210219 | 17 | 44018519 | T | C | 0.78 | 0.2607 | 0.0275 | 60613 | 2.657e-21 | 0.79 | -0.2681 | 0.0301 | 28104 | 5.582e-19 |
| rs76339698 | 5 | 60372588 | A | G | 0.89 | -0.1818 | 0.0263 | 62649 | 5.00e-12 | 0.90 | 0.0423 | 0.0422 | 28104 | 0.3171 |
| rs76904798 | 12 | 40614434 | T | C | 0.15 | 0.1352 | 0.0235 | 62649 | 9.224e-09 | 0.15 | -0.0563 | 0.0348 | 28104 | 0.1059 |
| rs7695720 | 4 | 77183300 | A | C | 0.79 | 0.1255 | 0.0208 | 62649 | 1.528e-09 | 0.78 | 0.0567 | 0.0294 | 28104 | 0.05426 |
| rs858290 | 7 | 23248036 | A | G | 0.61 | 0.1036 | 0.0177 | 62649 | 4.443e-09 | 0.60 | 0.0166 | 0.0251 | 28104 | 0.5095 |

Instrumental SNPs select for the PD to BAG classical Mendelian randomization analysis. CHR: chromosome; POS: hg19 genomic position; A1: effective allele; A2: the other allele; A1_exp_frq: A1 frequency in the exposure dataset; Beta_exp, effect of A1 for the exposure; SE_exp; standard error for Beta_exp; N_exp: sample size for exposure; P_exp, association p value for exposure; A1_out_frq: A1 frequency in the outcome dataset; Beta_out: effect size of A1 on outcome; SE_out: standard error of Beta_out; N_out: sample size for outcome; P_out: association p value for outcome.

### **Table S16. Statistical finemapping for associated loci.**

| Locus | Lead SNP | PP_1 | PP_2 | PPI_1 | SNP2 | PPI_2 |
| --- | --- | --- | --- | --- | --- | --- |
| Chr3:183892867-183975709 | rs73185796 | 0.51 | 0.46 | 0.7 | rs11402585 | 0.07 |
| Chr4:38591172-38779512 | rs13132853 | 0.74 | 0.26 | 0.16 | rs11727467 | 0.11 |
| Chr5:78388694-78451813 | rs79107704 | 0.75 | 0.25 | 0.48 | rs185486757 | 0.03 |
| Chr6:45407654-45511945 | rs2790102 | 0.84 | 0.16 | 0.05 | rs1934327 | 0.04 |
| Chr8:124661974-124682971 | rs7461069 | 0.82 | 0.18 | 0.1 | rs13340533 | 0.09 |
| Chr10:134544247-134597265 | rs4880424 | 0.85 | 0.15 | 0.08 | rs12767391 | 0.04 |
| Chr14: 88391116-88556525 | rs17203398 | 0.5 | 0.5 | 0.04 | rs10137195 | 0.04 |
| Chr17: 43101281-44863413 | rs2106786 | 0.85 | 0.15 | 0.01 | rs62062797 | 0.01 |

PP_1: posterior probability for the causal structural assuming only one causal variant in the locus. PP_2: posterior probability for the causal structural assuming only two causal variants.

PPI_1: posterior probability of inclusion to the one-causal variant structure; PPI_2: posterior probability of inclusion for the ranked second SNP to the one-causal variant structure;

### **Table S17. MR results using BAG as exposure.**

| outcome | effect | se | pval | padj | Model |
| --- | --- | --- | --- | --- | --- |
| AD | -0.0034 | 0.0062 | 0.59 | 0.74 | IVW |
| BIP | 0.0467 | 0.0276 | 9.04x10-2 | 0.15 | IVW |
| MDD | -0.0063 | 0.0212 | 0.77 | 0.77 | IVW |
| PD | -0.4012 | 0.1413 | **4.54x10-3** | **2.27x10-2** | IVW |
| SCZ | -0.0854 | 0.0372 | **2.18x10-2** | 5.46x10-2 | IVW |
| AD | -0.0542 | 0.0172 | **1.96x10-2** | **4.9x10-2** | Egger |
| BIP | 0.0765 | 0.1162 | 0.53 | 0.53 | Egger |
| MDD | 0.1092 | 0.0874 | 0.26 | 0.32 | Egger |
| PD | -1.4666 | 0.4074 | **1.14x10-2** | **4.9x10-2** | Egger |
| SCZ | -0.3272 | 0.1254 | **4.02x10-2** | 6.7x10-2 | Egger |
| AD | -0.0098 | 0.0061 | 0.11 | 0.14 | wMed |
| BIP | 0.0661 | 0.0272 | **1.51x10-2** | **3.77x10-2** | wMed |
| MDD | -0.0207 | 0.02485 | 0.41 | 0.41 | wMed |
| PD | -0.2741 | 0.0865 | **1.53x10-3** | **7.65x10-3** | wMed |
| SCZ | -0.0537 | 0.0308 | 8.11x10-2 | 0.14 | wMed |
| AD | -0.0034 | 0.0062 | 0.61 | 0.76 | PRESSO |
| BIP | 0.0467 | 0.0276 | 0.13 | 0.22 | PRESSO |
| MDD | -0.0063 | 0.0212 | 0.77 | 0.77 | PRESSO |
| PD | -0.2371 | 0.0861 | 7.06x10-2 | 0.22 | PRESSO |
| SCZ | -0.073 | 0.0344 | 8.72x10-2 | 0.22 | PRESSO |
| AD | 0.0018 | 0.0011 | 0.11 | 0.197 | RAPS |
| BIP | 0.0127 | 0.0051 | **1.27x10-2** | **4.87x10-2** | RAPS |
| MDD | 0.004 | 0.0042 | 0.34 | 0.43 | RAPS |
| PD | -0.0237 | 0.0101 | **1.95x10-2** | **4.88x1-0-2** | RAPS |
| SCZ | 0.0017 | 0.0051 | 0.74 | 0.738 | RAPS |
| AD | 2.14729 | 1 | 0.98 | 0.98 | CAUSE |
| BIP | 0.8014 | 1 | 0.79 | 0.98 | CAUSE |
| MDD | 1.9085 | 1 | 0.97 | 0.98 | CAUSE |
| PD | -0.1551 | 1 | 0.44 | 0.98 | CAUSE |
| SCZ | 0.4667 | 1 | 0.68 | 0.98 | CAUSE |

The six MR models are: IVW, Inverse variance weighted model; Egger, Egger regression; wMed, weighted median; PRESSO, MR-PRESSO; RAPS, MR-RAPS and CAUSE. causal effect size padj, FDR corrected p value; AD, Alzheimer’s disease; BIP, bipolar disorder; MDD, major depression disorder; PD, Parkinson’s disease; SCZ, schizophrenia. Note, negative CAUSE estimates indicate the existence of causal relations.

### **Table S18. MR results using disorders as exposure.**

| exposure | effect | se | pval | padj | Model |
| --- | --- | --- | --- | --- | --- |
| AD | 0.69 | 0.21 | **7.9x10-4** | **3.95x10-3** | IVW |
| BIP | 0.15 | 0.06 | **1.35x10-2** | **3.37x10-2** | IVW |
| MDD | 0.32 | 0.17 | 6.48x10-2 | 8.1x10-2 | IVW |
| PD | -0.25 | 0.12 | 3.35x10-2 | 5.59x10-2 | IVW |
| SCZ | 0.06 | 0.04 | 0.14 | 0.14 | IVW |
| AD | 0.48 | 0.30 | 0.13 | 0.16 | Egger |
| BIP | 0.68 | 0.22 | **3.67x10-3** | **9.18x10-3** | Egger |
| MDD | 0.29 | 0.91 | 0.76 | 0.76 | Egger |
| PD | -1.11 | 0.31 | **1.96x10-3** | **9.18x10-3** | Egger |
| SCZ | 0.39 | 0.14 | **5.66x10-3** | **9.44x10-3** | Egger |
| AD | 0.67 | 0.22 | **2.40x10-3** | **1.2x10-2** | wMed |
| BIP | 0.14 | 0.08 | 9.71x10-2 | 0.18 | wMed |
| MDD | 0.33 | 0.23 | 0.16 | 0.19 | wMed |
| PD | -0.02 | 0.07 | 0.75 | 0.75 | wMed |
| SCZ | 0.08 | 0.05 | 0.11 | 0.18 | wMed |
| AD | 0.69 | 0.21 | **2.22x10-3** | **1.11x10-2** | PRESSO |
| BIP | 0.15 | 0.06 | **1.67x10-2** | **4.11x10-2** | PRESSO |
| MDD | 0.32 | 0.14 | 5.87x10-2 | 0.07.34x10-2 | PRESSO |
| PD | -0.05 | 0.06 | 0.36 | 0.36 | PRESSO |
| SCZ | 0.08 | 0.04 | **2.46x10-2** | **04.11x10-2** | PRESSO |
| AD | 0.50 | 0.11 | **6.97x10-6** | **3.485x10-5** | RAPS |
| BIP | 0.05 | 0.02 | **9.05x10-3** | **0.01.8x10-2** | RAPS |
| MDD | 0.02 | 0.03 | 0.56 | 0.56 | RAPS |
| PD | -0.01 | 0.01 | 0.451 | 0.556 | RAPS |
| SCZ | 0.04 | 0.02 | **1.08x10-2** | **1.8x10-2** | RAPS |
| AD | -0.59 | 1 | 0.28 | 0.99 | CAUSE |
| BIP | 1.01 | 1 | 0.84 | 0.99 | CAUSE |
| MDD | 3.18 | 1 | 0.99 | 0.99 | CAUSE |
| PD | 1.44 | 1 | 0.93 | 0.99 | CAUSE |
| SCZ | -0.09 | 1 | 0.47 | 0.99 | CAUSE |

The six MR models are: IVW, Inverse variance weighted model; Egger, Egger regression; wMed, weighted median; PRESSO, MR-PRESSO; RAPS, MR-RAPS and CAUSE. causal effect size padj, FDR corrected p value; AD, Alzheimer’s disease; BIP, bipolar disorder; MDD, major depression disorder; PD, Parkinson’s disease; SCZ, schizophrenia. Note, negative CAUSE estimates indicate the existence of causal relations.
